# Survey of residential indoor Particulate Matter measurements 1990-2019

**DOI:** 10.1101/2021.11.10.21266177

**Authors:** Vito Ilacqua, Nicole Scharko, Jordan Zambrana, Daniel Malashock

## Abstract

We surveyed literature on measurements of indoor particulate matter in all size fractions, in residential environments free of solid fuel combustion. Data from worldwide studies from 1990-2019 were assembled into the most comprehensive collection to date. Out of 2,752 publications retrieved, 538 articles from 433 research projects met inclusion criteria and reported unique data, from which more than 2,000 unique sets of indoor PM measurements were collected. Distributions of mean concentrations were compiled, weighted by study size. Long-term trends, the impact of non-smoking, air cleaners, and the influence of outdoor PM were also evaluated. Similar patterns of indoor PM distributions for North America and Europe could reflect similarities in the indoor environments of these regions. Greater observed variability for all regions of Asia may reflect greater heterogeneity in indoor conditions, but also low numbers of studies for some regions. Indoor PM concentrations of all size fractions were mostly stable over the survey period, with the exception of observed declines in PM_2.5_ in European and North American studies, and in PM_10_ in North America. While outdoor concentrations were correlated with indoor concentrations across studies, indoor concentrations had higher variability, illustrating a limitation of using outdoor measurements to approximate indoor PM exposures.

**Practical implications:**

- Residential indoor PM concentration ranges for several size fractions measured in different worldwide regions are provided and may inform future public health research and practice, including PM exposure and risk assessment, and evaluation of IAQ-related interventions and consumer products, such as portable air cleaners.
- This long-term indoor PM concentration record provides insights regarding the degree of change in observed indoor PM concentrations by world region and some of the factors contributing to increasing or decreasing temporal trends.
- Outdoor air pollution remains a major influence on indoor concentrations of PM of all sizes.
- Greater variability of indoor concentrations of PM relative to outdoor concentrations demonstrate the potential for exposure misclassification when using outdoor concentrations to estimate indoor exposures and risk.
- IAQ interventions, including removing environmental tobacco smoke or using filtration-based portable air cleaners, can produce major improvements in IAQ through reduced indoor particle concentrations.

## 1. Introduction

The public health burden of airborne particulate matter (PM) is well established and supported by lines of evidence drawn from environmental epidemiology, toxicology, and controlled exposure studies ^1^. This abundant and consistent scientific evidence has prompted national and international standards to reduce the public health burden of PM in ambient air. This includes regulatory standards in the United States under the Clean Air Act and its amendments, and similar regulations internationally. These regulations have generally mandated official monitoring of PM and other air pollutants to ensure statutory limits are achieved. Consequently, an extensive record of reliable, comparable, and population-representative measurements has been generated, which has contributed to increased understanding of the population-level exposure to, and burden associated with ambient PM. The majority of a population exposure to PM, however, typically happens indoors ^2–4^ and in residential settings, where people spend the vast majority of their time ^5, 6^, and where levels of air pollutants may differ from those in ambient air ^2, 7, 8^. No comprehensive record of PM levels indoors exists, however, that is comparable in terms of temporal coverage, methodological consistency, and population representativeness to that available for PM in ambient air. This comparatively limited record can be explained by several factors including legal, economic, and ethical barriers, as well as practical challenges relating to residential access ^9–11^. Underlying all these factors, the greatest methodological challenge is the heterogeneity of indoor environments and indoor air quality, both in terms of temporal variability of indoor pollutant concentrations, influenced by individual human behaviors, building materials and design, and local climate, and the fragmentation of indoor air into a multitude of micro-environments within and between buildings.

Despite the absence of a systematic and extensive indoor PM record, scientific interest in indoor air quality has sustained the measurement of PM indoors for just as long as ambient air. Numerous *ad-hoc* research studies have measured indoor PM concentrations and have contributed to the understanding of the levels of indoor PM as well as the building-related, behavioral, geographic, and temporal factors that affect their variability. These research studies have necessarily varied in terms of goals, scope, resources, target populations, sampling and measurement methodology, study duration, observation conditions, and other important characteristics. With few exceptions, e.g., ^12–14^, these studies did not attempt to be truly population representative. Nevertheless, they contribute objective observations and understanding of an otherwise uncharted landscape of possible indoor air quality conditions. The information collected through measurements and modeling in existing studies allows us to understand how indoor PM characteristics and concentrations vary based on: PM in ambient air; building tightness; heating and cooling systems and operation; size, geometry and available surface area of a space; the nature, frequency, and intensity of human behaviors and activities such as smoking, cooking, burning candles, or vacuuming; the operation of windows, mechanical ventilation, and air cleaning devices; the generation of secondary organic particles through indoor chemical reactions; and more ^2, 8, 15–20^. Yet, the generalizability of existing studies is limited by their focus on specific geographic areas, episodic duration of measurement, and other constraints. The high cost and participant burden of large observational studies has limited more comprehensive population-level assessment. Consequently, most individual studies do not offer the panoramic view that is necessary to answer larger questions concerning PM in residential environments, such as:

- What are the levels of PM normally found inside homes?
- How do these levels vary geographically?
- How have they changed over the years?
- How much does outdoor PM contribute to indoor PM?

### 1.1. Purpose

To contribute to addressing these questions, we surveyed available literature on indoor PM measurements in residential environments worldwide and assembled a comprehensive collection of measurement data. This work aims to document the range of observed indoor PM concentrations and advance the understanding of long-term trends and their determinants, to improve exposure estimates for air pollution epidemiology and risk assessment, allow comparative analyses, inform the development of indoor air quality products, and provide context for policies targeting the role of the built environment in public health. Study findings may also increase awareness of the fundamental continuity in relationship between indoor and outdoor air quality, and of the need to address both in order to reduce the health burden of PM exposures, in the face of the widespread separation of both research and policy between indoor and outdoor domains.

It is acknowledged upfront that no such undertaking can be truly comprehensive, with limitations ranging from selection in study publication and database indexing, to language-based compartmentalization, to imperfect search criteria and retrieval. But if high diversity characterizes the air quality of indoor environments, then the larger the set of data, the better this diversity can be described, provided it was sampled randomly. Most of the research studies on residential indoor PM were not designed to be representative of entire countries, cities, building types, cooking behaviors, or health status, for example, nor was sampling truly random, although some degree of randomness was attempted in the larger studies. The same variety of study characteristics noted above, however, suggests that each project may miss or oversample subpopulations in its own specific way, so that when enough of them are considered together they may approach something similar to a stratified sample (albeit non-proportional). This agglomeration is still short of a population-representative sample in important ways but lessens the chances that the range of possible conditions (e.g., extreme concentrations, unusual sources, or behaviors) will be missed. In the near absence of data that is truly representative of national populations, a literature survey that reflects the current knowledge on indoor PM can still serve to formulate additional research questions and to guide measures aiming to reduce high exposures and the associated public health risks.

### 1.2. Scope

We focused this review on studies reporting indoor PM measurements, worldwide but limited to residential environments comparable to those in the United States and other high-income countries. Indoor environments across the world share several characteristics including the limited rate of air exchange between indoor and ambient air, a low level of solar radiation, effective barriers against precipitation, high surface to volume ratio, and complex systems to control temperature, supply water and energy, and remove water- and air-borne waste. Despite all their differences in architecture, climate, and culture, these elements of indoor environments are easily identifiable in apartments, detached homes, elderly care facilities, and dormitories of higher-income areas across the world, and we therefore included all residential arrangements. These similarities among residences around the world, however, are no longer consistent in buildings below a certain economic status, and specifically below the point on the energy ladder where solid fuels or biomass are used. The public health burden of indoor air pollution where solid fuels are used overshadows that of other air pollution exposures ^21^, offering little insight applicable to managing indoor air pollution in high-income countries. Rather than exclude regions of the world where biomass fuels are prevalent, which could miss data from many suitable buildings from regions with great economic disparity, we opted to exclude measurements where solid fuels use was reported. As an exception to this, we still included data from homes reporting wood burning for recreation or space heating, which are common particularly in colder climates, regardless of economic status ^22^.

This survey collected studies from the broadest set of metrics that are used to represent PM concentrations (indoor or outdoor) - by mass, number, or surface area - and categorized them into different size classes of particle aerodynamic diameter. The elemental composition of PM has been investigated by several studies. While the relevant literature was collected as a part of this project, this information was considered beyond the scope of this survey, at this stage.

### 1.3. Existing literature reviews

Review articles on the topic of airborne particles in residential indoors environments do exist. To the best of our knowledge, however, no single existing literature review covers the wide range of published studies that are included in this survey. Some of the published reviews are narrative in nature and provide an inventory of airborne particle concentrations from the individual studies and refrain from pooling data across studies in a quantitative manner ^23–25^. Table S1a summarizes existing literature reviews that do not provide pooled summary statistics. Other reviews carried out analyses of the data across studies and reported corresponding summary statistics. Those reviews are summarized in Table S1b, which also includes the researchers’ quantitative approach for obtaining pooled statistics. Most of the summary statistics were obtained by weighing the number of measurements within each study ^26, 27^.

## 2. Methods

### 2.1. Literature search strategy

Peer-reviewed publications were collected from *PubMed* and *ISI Web of Science* databases. Search strategies were optimized by trial and error to privilege completeness, even at the cost of a higher rate of irrelevant results. Publications in any language were searched, if at least an English-language abstract was available in the databases. The two primary search criteria for publications included keywords identified anywhere in the publication (title, abstract, and text) referring to PM (including synonyms and subtypes) and to indoor environment (and synonyms). A requirement for mentions of measurement (and synonyms) was included to decrease results only reporting modeling, policy perspectives, or commentary on the topic. Results were then limited to residential environments through a broad range of descriptors. Finally, articles primarily concerned with household use of solid fuel combustions (limited to mentions in abstract or title) were excluded. The search criteria are listed in Table 2-1, without consideration for the specific syntax required of these databases. The ability of the search to retrieve a predetermined set of relevant papers was also tracked throughout the search development process to ensure the search sensitivity was in agreement with study goals.

Collected abstracts were screened by human readers to exclude irrelevant results. Publications whose relevance to the study could not be determined from their abstract alone were included, along with those articles deemed relevant, for full-text review. As a quality control measure, abstracts not selected for full-paper review were evaluated by a second reviewer, and in cases of discordant opinion regarding study eligibility, were included for full-text review. The results of this process of literature search and publication review are shown in Figure 1.

**Figure 1.**
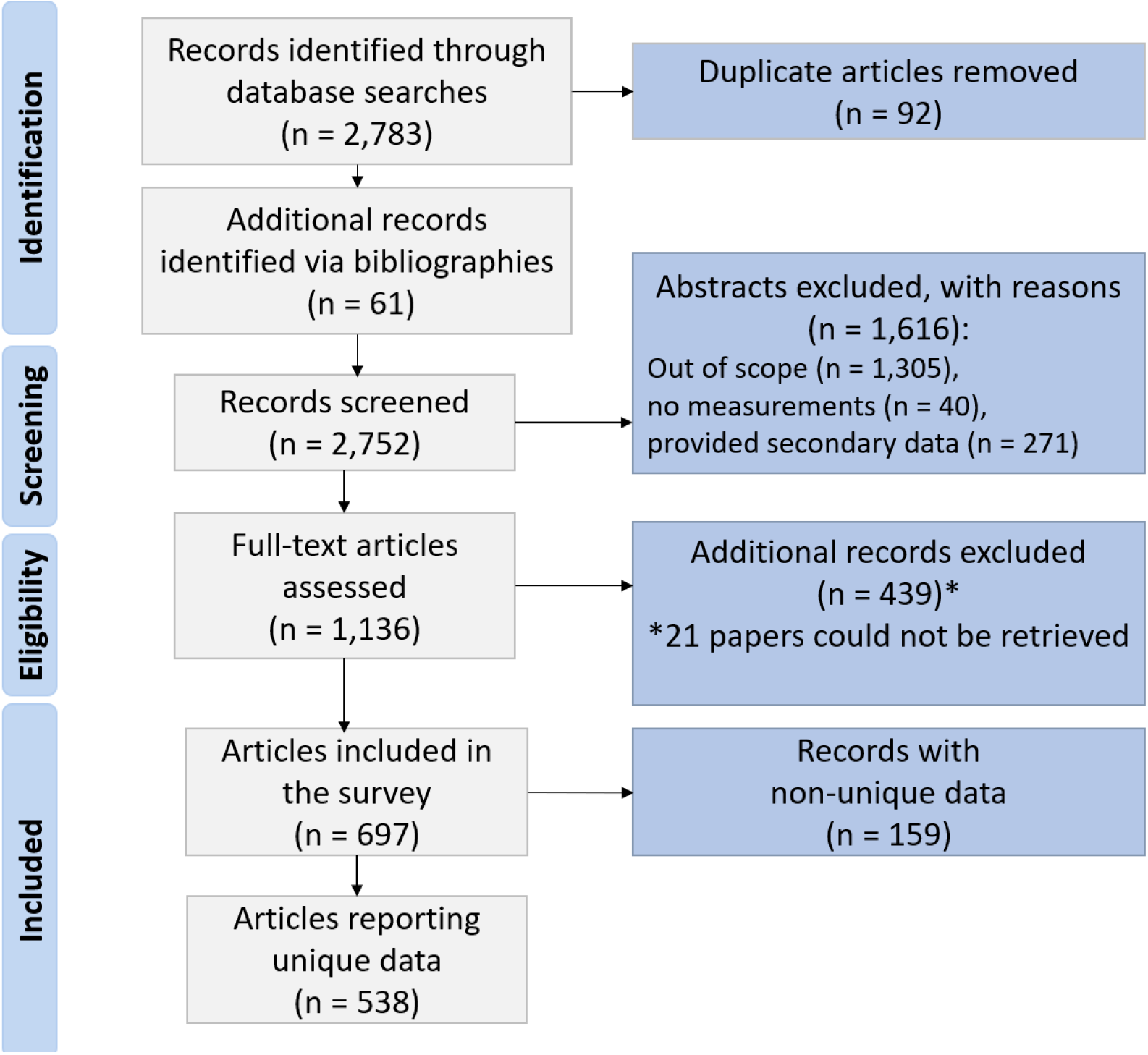
Results of literature search and papers review

### 2.2. Data extraction

Publications selected for full review were evaluated by a human reader against inclusion criteria. Data from publications deemed relevant were extracted using a standard operating procedure and entered into a database using standard forms designed to minimize data entry errors. Reasons for exclusion were noted. Additional possibly relevant papers not captured by the original searches were obtained from the literature cited. Papers resulting from a single research project (e.g., RIOPA, EXPOLIS, French Building Survey) were characterized in the database. Relationship to an existing research project was discerned by the reviewers based on mention of a project name in text or dates, author names, sampling locations, and grants acknowledged. Where an existing project name was not available an *ad-hoc* name was created using the study location and starting year (Figure 7). Primary data extracted included statistics on PM concentrations for sets of measurement samples reported. Some papers reported results for multiple sets of samples, distinguished by type (e.g., TSP and PM_2.5_), geography, location (i.e., indoor, outdoor), season, time of day, or other criteria used to establish contrasts (e.g., cooking vs. not, proximity to road, and weekday vs. weekend).

Journal articles were examined for use of established sampling methodology, but the size of this survey (number of included publications) prevented more rigorous evaluation of individual study methodology, including data quality evaluation and classification that may be used as a weighting factor. Authors’ and peer-reviewers’ assessment of the adequacy of the methodology to measure PM concentrations was not questioned during the data-extraction phase. However, outlier concentrations were flagged and checked for special circumstances during sampling and possible methodological flaws. It is also worth noting that the data extraction process in this survey is limited to the measurement results as reported and ignores the interpretations of results offered by authors to address individual study goals, where much of the variability in quality between individual publications resides.

Indoor PM concentrations measured by fixed or portable samplers or monitors, in any number of indoor residential environments, for any length of time, and with any analytical method were extracted. In addition, indoor concentrations resulting from personal exposure studies were extracted, if the authors reported the calculated concentrations for the residential indoor environments and the average time spent in that microenvironment. Outdoor concentrations were extracted from both on-site paired outdoor air sampling or paired ambient air monitoring, but coded separately. Measures of central tendency of these concentrations (mean, median, and geometric mean) were extracted as available, as well as second moments (e.g., standard deviations) and extreme values (min, max).

### 2.3. Statistical Analyses

The analysis of data collected from a variety of studies with different methodologies, time scales, and data distributions requires establishing a framework with explicit assumptions to avoid introducing the risk of improper inference. We defined the statistical universe (or population) of interest as the concentrations in all residential environments (within limitations stated), at all points in time. This statistical space was sampled by individual measurements of PM concentrations (almost always) in a single spot within selected indoor environments, under the assumption that these environments are sufficiently homogeneous over the time scale of the measurement. This assumption may introduce a bias, but the potentially much bigger source of bias is the general lack of randomness in the overall sampling, as noted, leaving open the possibility that certain regions of the statistical universe are either ignored (e.g., locations less accessible by researchers; holiday periods) or oversampled (e.g., homes of researchers themselves; evenings after-work hours). This is an intrinsic limitation that should be kept in mind in interpreting all results. All statistical analyses were performed in R 3.6.1 ^28^ (through R Studio 1.2).

#### 2.3.1. Calculated means and excluded measurements

Publications reported central tendency estimates for Indoor PM measurements in several different ways, depending on their data and purpose. The arithmetic mean was reported for more than 80% of the indoor measurements, but only the median for three quarters of the remaining measurements, and the geometric mean for the rest. For consistency of presentation, and to meet the requirements of some analyses, we calculated mean values, where possible, for the measurements that did not report them. Exact formulas exist to calculate the arithmetic mean from the geometric mean and geometric standard deviation^29^. In other cases, the mean was estimated from the median, sample size and extreme values using the methods by McGrath et al.^30^ with the R package *estmeansd*^31^. The performance of this latter method was evaluated by comparing estimated means to reported means in cases where both were available. The results were satisfactory (Table S2): the median relative error was +2.0% (so a slight bias towards overestimation), the median absolute relative error was 6.4%, and its (outlier) maximum was 376% (for a particle number). About 8% of means used in further analyses were calculated. Almost 13% of the measurements (indoor or outdoor) did not provide enough information to estimate the mean and were excluded from the work presented here.

#### 2.3.2. Weighting

The total duration of measurements in the included studies varied by over 6 orders of magnitude in scale, from measurements lasting a few minutes in a single home, to large field campaigns monitoring concentrations over weeks or months in hundreds of homes. The ability of a study to capture the variability of indoor PM (between homes and over time) necessarily reflects this range, though even small studies do contribute to our understanding. Throughout the statistical analyses performed, from descriptive statistics to regressions, we therefore used weights proportional to the total amount of time sampling was performed, unless otherwise noted. These weights can be most easily understood as the product of the number of homes in a sample and the average duration of sampling in each home over the study, expressed in units of buildings × hours. Figure S1 shows the distribution of this weighting factor. To avoid possible confusion, the expression *weighted regression* in this work uses the weights above and does not refer to the commonly used approach of weighting by the inverse of the standard deviation, which in our case can reflect actual variability more than measurement error.

#### 2.3.3. Regressions

Long-term temporal trends and the role of outdoor concentrations were explored through multiple regression, controlling for other factors, such as different locations, absence of environmental tobacco smoke (ETS), or use of air cleaners when possible. Multiple regressions are general and flexible tools, but ordinary least-squares linear regression was generally not appropriate for our data since the assumption of normality of the residuals was usually violated. Indoor concentrations (the dependent variable) were approximately log-normally distributed, as is common with PM concentrations^29, 32^, as were the residuals. Different approaches can be used^33^, and often the assumptions of normality and homoscedasticity can be ignored by relying on the central limit theorem, but in many cases our sample sizes were fairly small. Another common approach, the log transformation of the data, has been shown to provide biased estimates for lognormally distributed data^34^. This also has the drawback of less intuitive multiplicative interpretation of results, rather than absolute change. We therefore addressed these issues with a non-linear regression with lognormal error distribution^35^ and identity link, with the R package *logNormReg*^36^. All regressions were weighted as described above. The implications for the results of using this approach rather than a linear model or a log-transformation can be seen as an example in Figure 8, where the same data are fitted using different approaches. Although all methods perform similarly near the mean, both slope and intercept are affected by the choice.

There is no universally accepted way to assess the fit of a model, or the amount of variation explained in a non-linear model. With a non-linear regression, the coefficient of determination R^2^ can no longer generally be used^37^, because the residuals sum of squares (RSS) and the regression model sum of squares (MSS) do not add up to the total sum of squares (TSS). However, when TSS ≈ MSS +RSS, a pseudo-coefficient of determination that preserves its definition as R^2^_d_ = MSS/TSS, could still be informative, and more readily interpreted than other goodness of fit measures. Since this was the case in almost all our regressions, we reported a R^2^_d_, along with its distortion (RSS + MSS -TSS)/TSS. A positive distortion value indicates R^2^_d_ exceeds the true fraction of variance explained and a negative value indicates it falls short of it.

Regressions using the larger geographic groupings (e.g., regions) assume a homogeneity of the residuals variance among groups, which may not be the case. On the other hand, pooling together more observations can help estimate additional parameters. Our approach, therefore, was first to perform regressions using smaller geographic units (e.g., country), when numerous observations were available. Then, for regional estimates, we pooled together observations from the largest groups we could build that failed to reject the hypothesis of homogeneous residuals variances with the Fligner-Killeen non-parametric test. When more limited numbers of observations were available, we simply performed pooled regressions first, and then separate regressions on subgroups when possible.

Regressions of indoor concentrations to outdoor (or ambient monitor) concentrations can have a physical interpretation. The time-varying PM concentration indoors can be approximated^38^ by

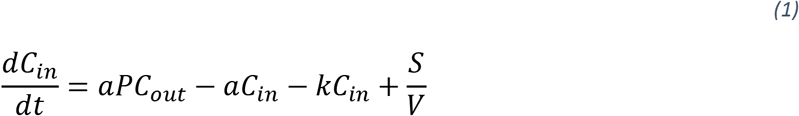

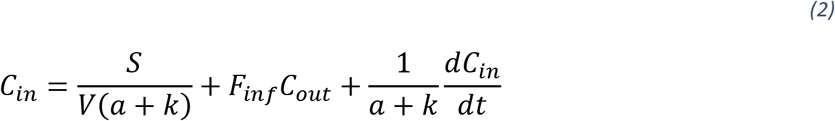

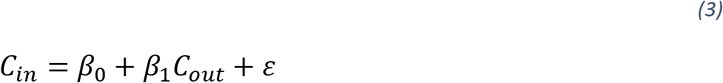

where *C_in_* and *C_out_* are the indoor and outdoor concentrations, respectively, *a* is the air exchange rate, *P* is the fraction of particles penetrating the building envelope, *k* captures the sum of all processes removing particles indoors (deposition or filtration) and *S* is the particle emission rate from all indoor sources in an indoor space of volume *V*. The fraction F_inf_ *= aP/(a+k*) is often referred to as infiltration factor, representing the ratio of competing processes adding and removing particles from the outdoors. Rearranging (1) into (2), yields an equation that can be written as a regression model (3)^39^, where *β_0_* and *β_1_* are the coefficients to be estimated by the regression. To the extent that measurements are intended to yield representative mean concentrations, the time derivative term is small and random, and can be considered part of the error (*ε*) in the regression models.

## 3. Results and Discussion

### 3.1. Literature search results

A total of 2844 publications were retrieved from literature searches (n=2,783) and identified while reviewing references of collected literature (n=61), including 92 duplicate publications. More than 60% of these results (n=1,616) were excluded based on abstract screening alone. The remaining 1,136 papers required review of the full journal article text (Figure 1). The review resulted in the classification of 697 papers as relevant to the goals of this survey and its boundaries, of which 23% reported only indoor PM concentration data that had been reported in other publications. A total of 538 articles reported unique data that was included in reviewer extractions.

Relevant papers could be traced to 433 different research study projects, conservatively attributed based on the criteria outlined in 2.2. In a few cases, where the same researchers published results on sampling cohorts over longer time spans and with multiple sources of funding, distinctions between projects were unclear and results from different papers were attributed to separate projects. An average of 1.6 papers were published for a study project. The number of published papers per decade reporting data on indoor PM increased by a factor of 9 over the timespan of this survey (Figure 2), from 45 in the first decade (1990-1999) to 413 in the latest (2010-2019). This increase is faster than the general growth rate in scientific journal article publications (by a factor of 3 for PubMed index)^40, 41^, suggesting a growing interest in indoor air quality, especially in East Asia.

**Figure 2.**
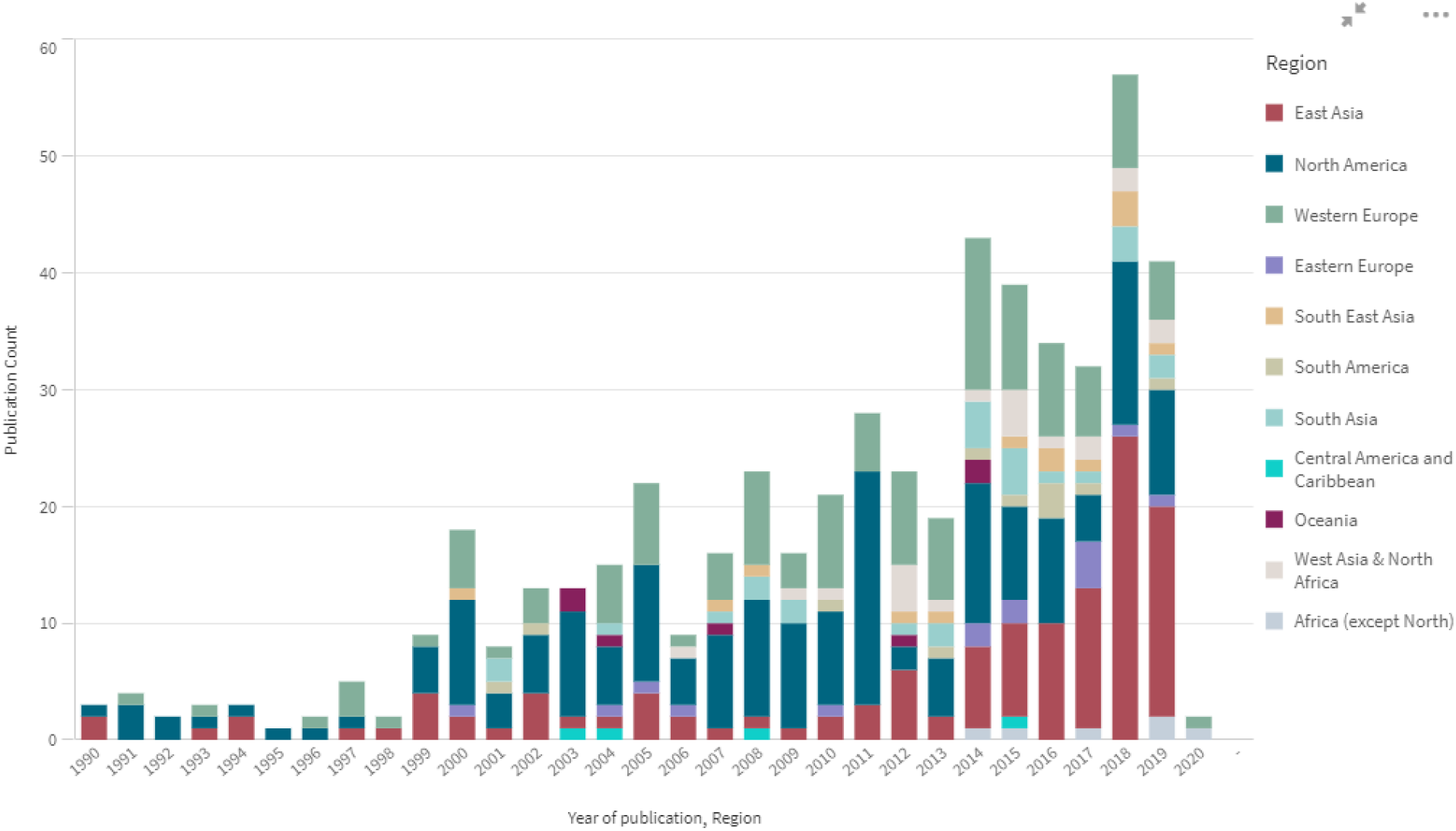
Publications with data included in the survey. Some journal articles with a 2020 date of publication were available in 2019.

The degree of interest in different PM sizes is reflected in the number of research projects and publications investigating them (Figure 3), with PM_2.5_ claiming the greatest attention of researchers and twice as much as PM_10_, the next most researched fraction. In a few cases, investigators opted to measure unusual size fractions (e.g., PM_0.5_, PM_3.5_, PM_>4_), or reported actual cut-off sizes they verified for their samplers. Where possible, these sporadic results were combined with similar size fractions (e.g., PM_0.25-2.5_ mass with PM_2.5_ mass), but otherwise proved to be too infrequent to lend themselves to further analysis here. Ultrafine particles (UFP) were variously defined, but most commonly as those with an aerodynamic diameter less than 0.3 μm, so we used that cutoff size in this study.

**Figure 3.**
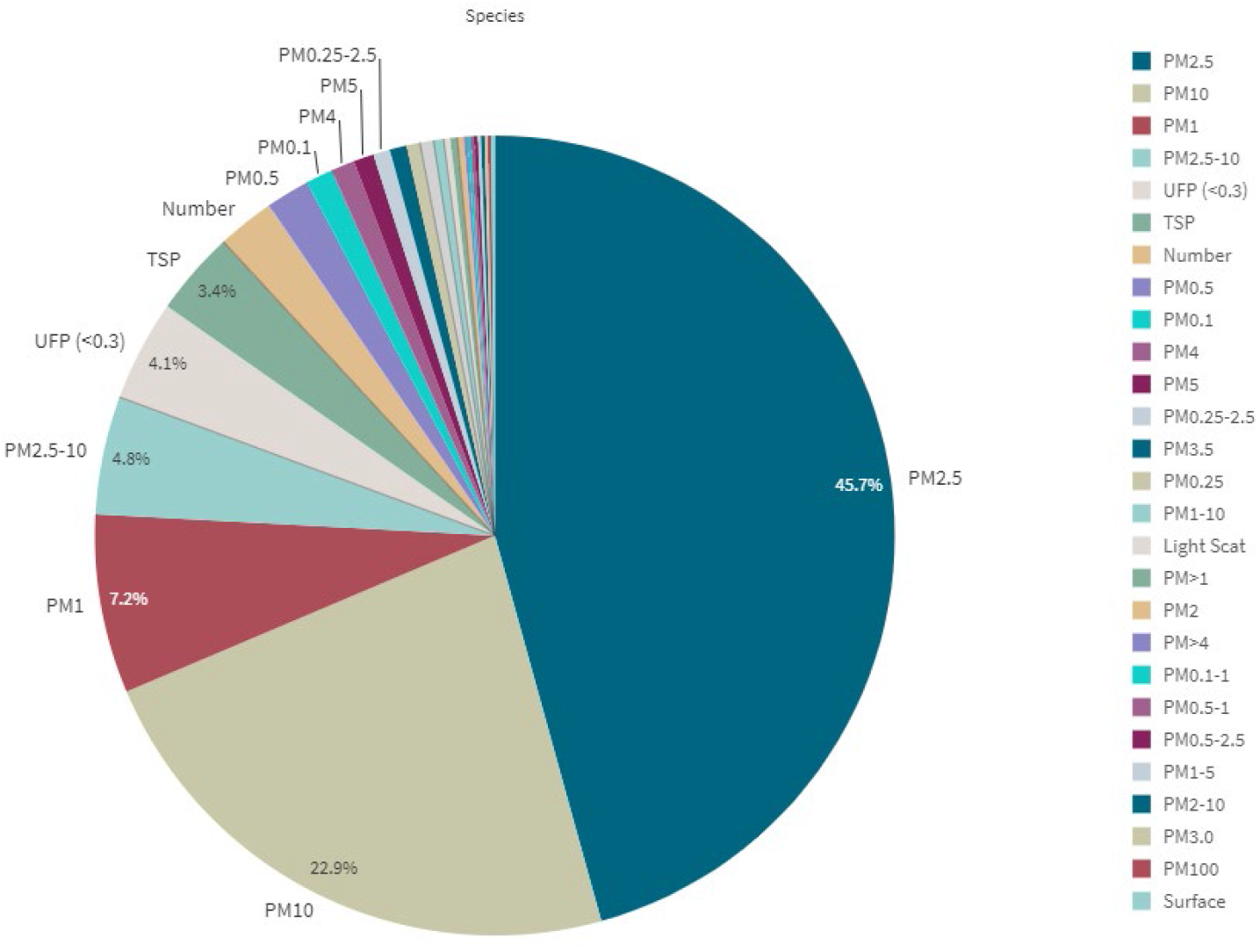
Fraction of studies investigating the different PM size fractions and other PM metrics.

### 3.2. Sampling approaches

Collectively, the research projects in this survey sampled more than 21,200 homes, adding up to 4.91 million hours of sampling time across those homes. Individual studies varied in breadth of sampling from a single home (in about 14% of the studies) to 755^42, 43^, with a median number of 16 homes. The median duration of a study sampling campaign was 13 weeks. Samples were often collected repeatedly, ranging from repeated measures within a single day to 3 years ^44^; the median time between first and last home visit was 5 days. The sampling time for reported measurement statistics also varied substantially, from 5 seconds (basically real-time monitoring) to 3 months of continuous monitoring^45^, but the median was 24 hours. Even the largest studies apportioned their resources to either sampling numerous homes or collecting samples of long duration, but rarely both. Logistical and funding limitations likely prevented the collection of long-term samples in a large number of homes.

A variety of building types and residential arrangements were sampled by the different studies, with multi-unit buildings (of unspecified height) most reported (14% of the studies), followed by single-family homes (11%), although, in more than 60% of studies, the type of home remained unspecified, or results were only reported for mixed types. Similarly, within a home, 36% of studies did not specify where air samples were collected. Among studies that reported sampling location(s), living rooms (or equivalent room where most time was spent), bedrooms, and kitchens accounted for 78%, 23%, and 17% of the sampling locations, respectively. Less than 10% of studies reported sampling multiple rooms.

### 3.3. Indoor PM Concentrations

#### 3.3.1. Descriptive statistics

Frequency distributions of weighted mean PM concentrations for the three regions with most data (Figures 5 and 6; Figures S2 to S5 show all other regions) vary by up to 3 orders of magnitude and are right-skewed. These general patterns similarly apply to unweighted means, which are even more widely distributed and skewed towards higher values (distributions not shown). Modal sections of the distributions across regions cluster around similar values for North America, Western Europe, and, based on more limited data, also Eastern Europe, at 5-10 μg/m^3^ for PM_2.5-10_ (coarse particles); 5-30 μg/m^3^ for PM_2.5_; 5,000-20,000 cm^-3^ for UFP; and, less sharply, 0-25 μg/m^3^ for PM_1_. Distributions of PM_10_ are bimodal in North America and Western Europe, clustering around 15-25 and 50-60 μg/m^3^. This signifies that most homes are relatively similar across these regions in terms of indoor PM, and regions mostly differ on the frequency of extreme values. Size distributions for East Asia are generally more spread out than for other regions, with a less clearly identifiable modal section, and generally higher values (except for PM_2.5 -10_). This pattern can reflect a great heterogeneity between sampling locations, drastic changes in concentrations over the time span considered, or both. To a degree, but with much less available data, a similarly spread-out distribution pattern can be seen for the larger size fractions (PM_10_, PM_2.5_, and PM_2.5-10_) in South Asia, and less clearly for West Asia & North Africa. The highest weighted mean concentrations are also occurring in these two regions. Much less data are available about other regions to identify consistent patterns, demonstrating again the scarcity of measurements for some regions in this survey.

**Figure 4.**
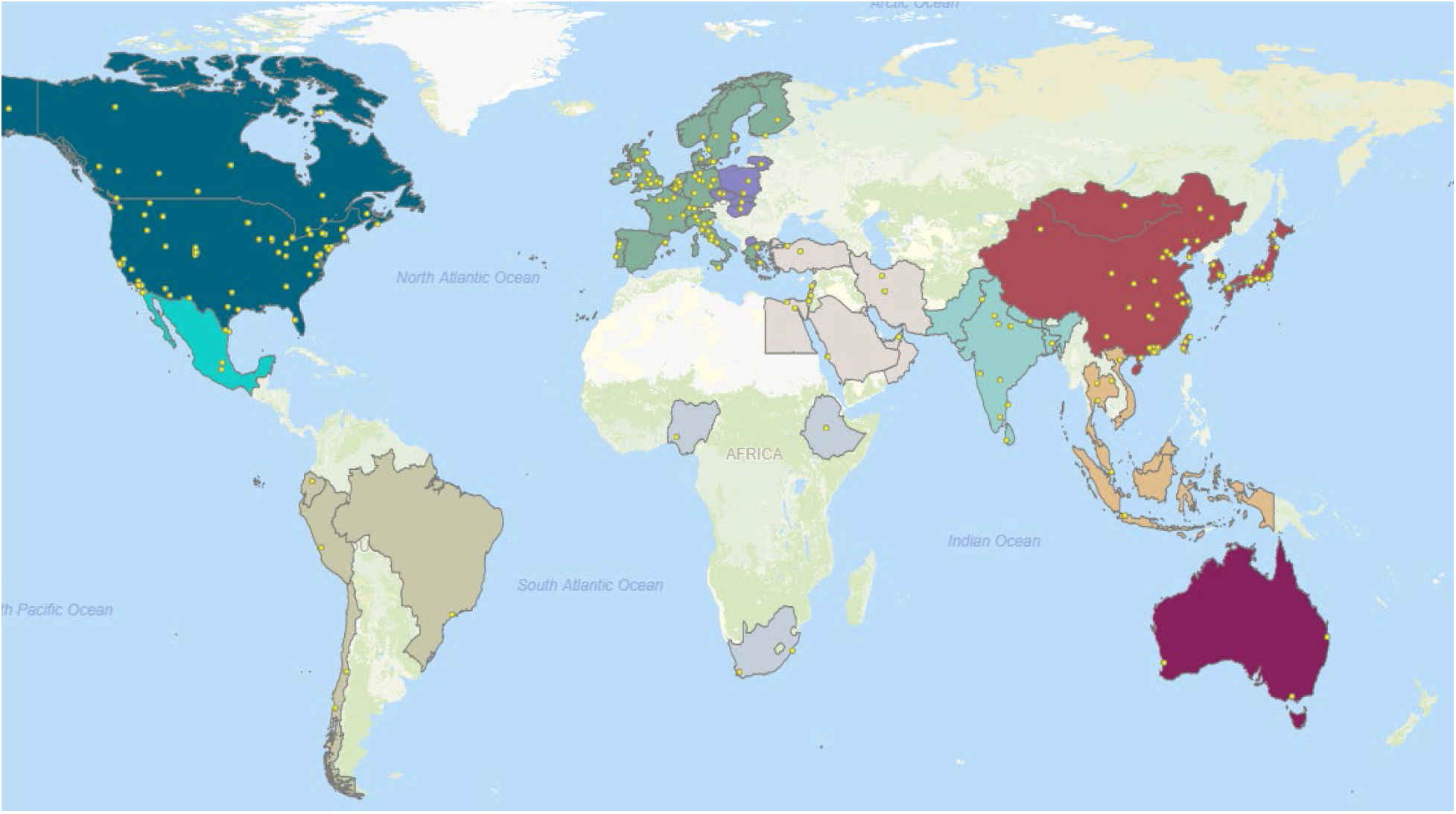
Location of sampling sites for studies included in the survey. Country colors identify the world region groupings used in the analyses. A few sampling locations could not be accurately mapped, and some locations markers completely overlap at this scale. Some nationwide studies are marked in the country capital

**Figure 5.**
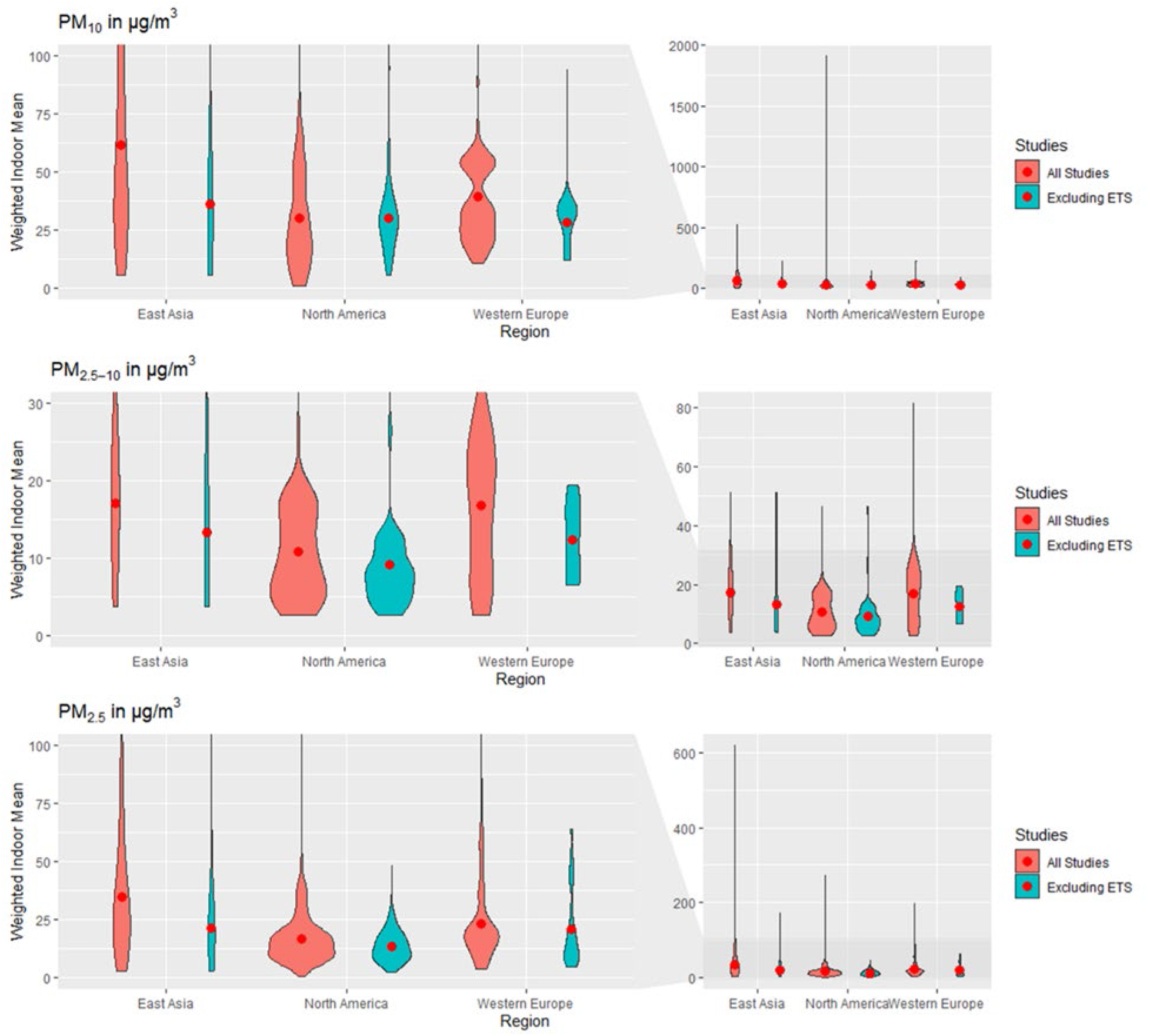
Weighted frequency distributions of means for PM10, PM2.5-10 and PM2.5 for East Asia, North America and Western Europe. Width is proportional to the number of means, within a size fraction. The red points are displaying the weighted indoor mean of means. Studies labeled ‘Excluded ETS’ are restricted to mean measurements in homes without environmental tobacco smoke. Left panels show magnified region of right panels.

**Figure 6.**
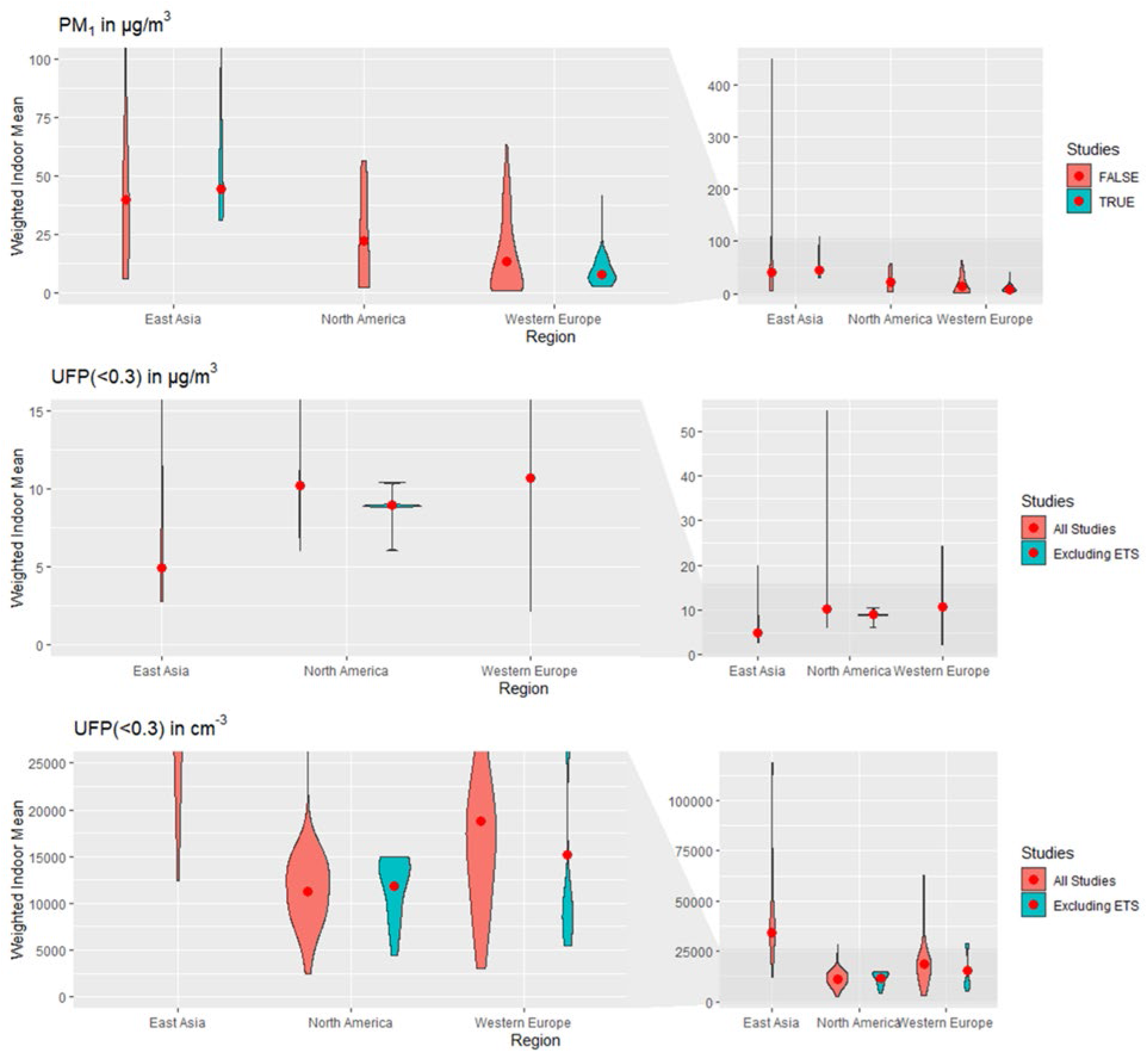
Weighted frequency distributions of means for PM1, UFP (in µg/m^3^ and cm^−3^) for East Asia, North America and Western Europe. The red points are displaying the weighted indoor mean of means, within a size fraction. Studies labeled ‘Excluded ETS’ are restricted to mean measurements in homes without environmental tobacco smoke. Left panels show magnified region of right panels

To meet the needs of exposure and risk assessments in particular, quantiles of the weighted mean distributions were tabulated along with the number of studies synthesized by each group, and the total number of homes and observation times they collectively gathered (Tables 2 – 7). Although some of these sampling numbers may seemingly add up to impressive statistical power (up to thousands of homes and hundreds of thousands of hours of observation), two key limitations should be considered when using these concentration data. First, these observations were not generally drawn from truly random samples of indoor residences or times (as discussed above). Second, they can represent multiple decades of observations of indoor concentrations, which may have been changing over time. Tables 8 through 11 provide the first and last year of observations, which are described in greater detail in Section 3.3.2. To fully characterize the range of PM exposure and risk, such as worst-case scenario analysis, two statistics on maximum concentrations were synthesized as well (Tables 2 – 7). Generally, the maximum of the distribution of weighted means is comparable to the median of maximum values. Means that far exceed the median of maxima could be reflecting special circumstances during sampling, such as wildfires, dust storms, or short-term measurements during cooking. Statistics on study minima are generally reduced to the trivial values of 0 (or the limits of detection) and are not presented.

**Table 1.** Search criteria. A * indicates a wild character, capturing every ending of the word.

| Item | Criterion |
| --- | --- |
| Limits | 1990 ≤ Publication Date ≤ 2019; Any language |
| 1 | Particulate(s) OR particulate matter OR ultrafine particle(s) OR PM10 OR PM2.5 OR fine particle(s) |
| 2 | Indoor* OR indoor air OR indoor environment OR IAQ OR IEQ |
| 3 | measure* OR concentration* OR characteriz* |
| 4 | residence* OR residential OR home* OR house* OR apartment* OR housing OR lodging* OR dwelling* OR condo* |
| 5 | biomass OR cookstove* ( searched only in Abstract OR Title) |
| Combined search | (1 AND 2 AND 3 AND 4) NOT 5 |

**Table 2.** Weighted statistics for means of PM10 and PM1-10 (μg/m^3^) measurements (including calculated means) and statistics for maximum of PM10 (μg/m^3^) measurements. Statistics labeled ‘No ETS’ are restricted to measurements in homes without Environmental Tobacco Smoke (ETS). Studies may report means for multiple locations and times (n is the total number of means). Not all studies provided a mean or a maximum.

| Region | Studies | Homes | Total Obs. Time (hr) | Statistics for Mean Measurements |  |  |  |  |  |  |  |  | Maximum |  |
| --- | --- | --- | --- | --- | --- | --- | --- | --- | --- | --- | --- | --- | --- | --- |
|  |  |  |  | n | Mean | SD | Wt. Mean | Min | Wt. 10% | Wt. 50% | Wt. 90% | Max | Median | Highest Value |
| All | 160 | 8228 | 8.95E+05 | 301 | 86.7 | 173 | 43.7 | 1.03 | 13.5 | 31.4 | 66.3 | 1910 | 115 | 3910 |
| No ETS | 62 | 1886 | 2.65E+05 | 98 | 59.4 | 72.4 | 43.1 | 5.6 | 17.9 | 34 | 56.2 | 518 | 102.8 | 985.8 |
| Africa (except North) | 3 | 791 | 1.83E+04 | 3 | 119 | 60.4 | 74.4 | 55 | 55 | 55 | 175 | 175 | 68.2 | 68.2 |
| Central America and Caribbean | 1 | 30 | 1.75E+03 | 3 | 44.3 | 13 | 51.5 | 31 | 31 | 57 | 57 | 57 | 97 | 154 |
| No ETS | 1 | 30 | 1.75E+03 | 3 | 44.3 | 13 | 51.5 | 31 | 31 | 57 | 57 | 57 | 97 | 154 |
| East Asia | 43 | 1676 | 1.23E+05 | 79 | 91.3 | 92.7 | 61.7 | 5.5 | 20.4 | 41 | 112 | 525 | 143.6 | 974.6 |
| No ETS | 17 | 264 | 7.39E+04 | 22 | 75.7 | 61.2 | 36.1 | 5.6 | 17.9 | 29.5 | 41 | 226 | 188 | 641 |
| Eastern Europe | 7 | 265 | 1.28E+04 | 14 | 39.5 | 21.4 | 32.9 | 15 | 20.8 | 28.1 | 66 | 79 | 75.15 | 202 |
| No ETS | 4 | 87 | 4.55E+03 | 7 | 50.6 | 23 | 49.2 | 22 | 22 | 38.8 | 79 | 79 | 52.42 | 90.3 |
| North America | 39 | 2258 | 2.90E+05 | 68 | 106 | 324 | 30.4 | 1.03 | 5 | 26.5 | 56.2 | 1910 | 105.6 | 3910 |
| No ETS | 18 | 573 | 6.30E+04 | 31 | 34.8 | 26.1 | 30.3 | 5.63 | 12.3 | 27.7 | 45 | 140 | 100.75 | 985.8 |
| Oceania | 3 | 142 | 1.53E+04 | 7 | 16.3 | 2.21 | 19.6 | 14 | 15.6 | 20.4 | 20.4 | 20.4 | 24.5 | 65 |
| No ETS | 2 | 118 | 1.51E+04 | 3 | 17 | 2.99 | 19.6 | 14.9 | 15.6 | 20.4 | 20.4 | 20.4 |  |  |
| South America | 4 | 225 | 4.77E+04 | 4 | 87.6 | 65.1 | 42.4 | 35.2 | 38.3 | 38.3 | 38.3 | 173 | 194.85 | 208.2 |
| No ETS | 2 | 146 | 4.58E+04 | 2 | 71 | 46.3 | 41.3 | 38.3 | 38.3 | 38.3 | 38.3 | 104 | 194.85 | 208.2 |
| South Asia | 14 | 56 | 5.57E+03 | 25 | 219 | 100 | 235 | 61 | 143 | 231 | 311 | 503 | 329 | 1381.81 |
| No ETS | 2 | 5 | 7.08E+02 | 5 | 216 | 47.4 | 249 | 166 | 184 | 265 | 265 | 268.2 | 375 | 734 |
| South East Asia | 5 | 613 | 9.23E+03 | 7 | 40.6 | 25.6 | 37.5 | 14 | 14 | 31.4 | 64.1 | 71.6 | 105.3 | 105.3 |
| West Asia & North Africa | 9 | 901 | 1.56E+05 | 13 | 114 | 127 | 107 | 37.9 | 51.7 | 51.7 | 156 | 518 | 240 | 492 |
| No ETS | 3 | 210 | 1.26E+04 | 5 | 157 | 203 | 188 | 38.1 | 38.1 | 104 | 518 | 518 | 192.55 | 240.1 |
| Western Europe | 35 | 1271 | 2.15E+05 | 78 | 37.4 | 30.7 | 39.3 | 10.7 | 18.3 | 34 | 54 | 227 | 92.3 | 3598.4 |
| No ETS | 13 | 453 | 4.78E+04 | 20 | 26.7 | 18.4 | 28.4 | 12.4 | 12.4 | 34 | 34 | 94 | 42.25 | 374 |

**Table 3.** Weighted statistics for means of PM2.5-10 (μg/m^3^) measurements (including calculated means) and statistics for maximum of PM2.5-10 (μg/m3) measurements. Statistics labeled ‘No ETS’ are restricted to measurements in homes without Environmental Tobacco Smoke. Studies may report means for multiple locations and times (n is the total number of means). Not all studies provided a mean or a maximum.

| Region | Studies | Homes | Total Obs. Time (hr) | Statistics for Mean Measurements |  |  |  |  |  |  |  |  | Maximum |  |
| --- | --- | --- | --- | --- | --- | --- | --- | --- | --- | --- | --- | --- | --- | --- |
|  |  |  |  | n | Mean | SD | Wt. Mean | Min | Wt. 10% | Wt. 50% | Wt. 90% | Max | Median | Highest Value |
| All | 53 | 2773 | 4.54E+05 | 97 | 24.2 | 37.7 | 14.5 | 2.6 | 4.65 | 13.6 | 21.9 | 221 | 37.63 | 769.56 |
| No ETS | 28 | 1042 | 1.89E+05 | 51 | 24.4 | 37.8 | 13.2 | 2.62 | 4.64 | 12.8 | 19.4 | 202 | 45.65 | 335.4 |
| East Asia | 7 | 242 | 7.95E+04 | 10 | 20.5 | 15.4 | 17.1 | 3.8 | 5 | 20 | 20 | 51.2 | 36.25 | 39.2 |
| No ETS | 4 | 154 | 5.54E+04 | 7 | 21.2 | 18.8 | 13.4 | 3.8 | 5 | 13.6 | 34.1 | 51.2 | 33.3 | 33.3 |
| Eastern Europe | 3 | 54 | 5.78E+03 | 5 | 7.32 | 2.42 | 8.25 | 4.4 | 4.4 | 7.8 | 11 | 11 | 11.6 | 11.6 |
| No ETS | 1 | 1 | 2380 | 3 | 7.07 | 0.751 | 7.1 | 6.3 | 6.3 | 7.1 | 7.8 | 7.8 |  |  |
| North America | 22 | 1307 | 1.73E+05 | 35 | 9.57 | 8.29 | 10.8 | 2.62 | 4.41 | 8.6 | 17.4 | 46.5 | 45.65 | 335.4 |
| No ETS | 12 | 631 | 7.32E+04 | 21 | 9.36 | 9.99 | 9.24 | 2.62 | 3.5 | 8.6 | 12.8 | 46.5 | 46.3 | 335.4 |
| South America | 3 | 207 | 4.75E+04 | 3 | 19.5 | 14.3 | 15.9 | 7.83 | 15.2 | 15.2 | 15.2 | 35.4 | 102.1 | 117.5 |
| No ETS | 2 | 148 | 4.61E+04 | 2 | 25.3 | 14.3 | 16.1 | 15.2 | 15.2 | 15.2 | 15.2 | 35.4 | 102.1 | 117.5 |
| South Asia | 5 | 18 | 2.48E+03 | 9 | 120 | 59.8 | 123 | 23.9 | 80.9 | 119 | 221 | 221 | 197 | 769.56 |
| No ETS | 2 | 5 | 7.08E+02 | 5 | 126 | 42.8 | 122 | 97.7 | 105 | 120 | 120 | 202 | 197 | 197 |
| South East Asia | 1 | 6 | 4.83E+02 | 1 | 27 | NA | 27 | 27 | 27 | 27 | 27 | 27 |  |  |
| West Asia & North Africa | 3 | 646 | 1.20E+05 | 3 | 27.8 | 8.52 | 29.2 | 22.1 | 22.1 | 23.7 | 37.6 | 37.6 |  |  |
| No ETS | 2 | 10 | 3.67E+03 | 3 | 27.5 | 8.73 | 29.3 | 22.1 | 22.1 | 22.1 | 37.6 | 37.6 |  |  |
| Western Europe | 9 | 293 | 2.53E+04 | 31 | 16.9 | 18.4 | 16.9 | 2.6 | 5.5 | 17 | 26.7 | 81.4 | 19 | 88 |
| No ETS | 5 | 92.5 | 7.72E+03 | 10 | 11.4 | 5.37 | 12.4 | 6.58 | 7 | 13.1 | 17 | 19.5 | 9 | 31.2 |

**Table 4.** Weighted statistics for means of PM2.5 and PM0.25-2.5 (μg/m^3^) measurements (including calculated means) and statistics for maximum of PM2.5 (μg/m^3^) measurements. Statistics labeled ‘No ETS’ are restricted to measurements in homes without Environmental Tobacco Smoke. Studies may report means for multiple locations and times (n is the total number of means). Not all studies provided a mean or a maximum.

| Region | Studies | Homes | Total Obs. Time (hr) | Statistics for Mean Measurements |  |  |  |  |  |  |  |  | Maximum |  |
| --- | --- | --- | --- | --- | --- | --- | --- | --- | --- | --- | --- | --- | --- | --- |
|  |  |  |  | n | Mean | SD | Wt. Mean | Min | Wt. 10% | Wt. 50% | Wt. 90% | Max | Median | Highest Value |
| All | 326 | 16269 | 3.16E+06 | 596 | 46.4 | 66.9 | 24.7 | 0.5 | 8.4 | 17.8 | 43 | 733 | 90.95 | 4641 |
| No ETS | 149 | 5810 | 9.71E+05 | 230 | 30.8 | 41.8 | 19.2 | 2.21 | 7.7 | 15 | 28.6 | 302 | 72 | 4378 |
| Africa (except North) | 3 | 59 | 7.62E+02 | 4 | 42.8 | 22.9 | 63.0 | 25.8 | 25.8 | 76.5 | 76.5 | 76.5 | 126.05 | 218 |
| Central America and Caribbean | 4 | 76 | 9.20E+03 | 15 | 41.6 | 12.1 | 32.7 | 26 | 26 | 31 | 35.1 | 68.5 | 67 | 141.8 |
| No ETS | 1 | 34 | 6.82E+03 | 4 | 29.5 | 4.36 | 30.8 | 26 | 26 | 31 | 35 | 35 | 91.5 | 115 |
| East Asia | 81 | 2930 | 1.07E+06 | 154 | 75.6 | 81 | 35.0 | 2.7 | 12.8 | 24.8 | 73 | 618 | 153 | 888.8 |
| No ETS | 33 | 821 | 1.99E+05 | 46 | 57.1 | 42.5 | 21.5 | 2.8 | 11.8 | 18.4 | 25.5 | 174 | 98.7 | 623 |
| Eastern Europe | 10 | 304 | 1.76E+04 | 18 | 23.5 | 15.7 | 21.4 | 4.9 | 9.3 | 15.7 | 53 | 55 | 74.35 | 203.2 |
| No ETS | 5 | 134 | 6.81E+03 | 8 | 28.7 | 16.8 | 32.6 | 11.4 | 15.7 | 25 | 55 | 55 | 60.62 | 96 |
| North America | 109 | 6518 | 1.06E+06 | 204 | 20.2 | 24.7 | 16.8 | 0.5 | 7.71 | 16.2 | 29.5 | 272 | 78 | 2100 |
| No ETS | 61 | 2712 | 4.73E+05 | 98 | 14.5 | 8.78 | 13.6 | 2.21 | 7 | 12.8 | 20.9 | 48.2 | 65.23 | 508.2 |
| Oceania | 3 | 132 | 1.60E+04 | 6 | 9.46 | 1.58 | 8.44 | 7.99 | 8.4 | 8.4 | 8.67 | 12 | 15.3 | 18 |
| No ETS | 2 | 118 | 1.53E+04 | 3 | 8.35 | 0.34 | 8.42 | 7.99 | 8.4 | 8.4 | 8.67 | 8.67 |  |  |
| South America | 10 | 606 | 1.20E+05 | 12 | 68.9 | 42.4 | 32.4 | 20 | 23.2 | 27.1 | 64.8 | 135 | 177.55 | 373.9 |
| No ETS | 3 | 150 | 4.65E+04 | 3 | 44.6 | 22.8 | 25.5 | 23.2 | 23.2 | 23.2 | 23.2 | 68.5 | 177.05 | 204.3 |
| South Asia | 19 | 452 | 4.47E+04 | 35 | 151 | 127 | 162 | 37.1 | 117 | 165 | 165 | 733 | 258.5 | 4378 |
| No ETS | 4 | 34 | 1.84E+03 | 8 | 134 | 103 | 194 | 40 | 40 | 285 | 302 | 302 | 196 | 4378 |
| South East Asia | 8 | 532 | 1.49E+03 | 14 | 48.2 | 76.6 | 41.5 | 5.8 | 19.8 | 39.2 | 70 | 308 | 1140 | 1240 |
| No ETS | 1 | 5 | 3.25E+02 | 2 | 8.65 | 4.03 | 9.22 | 5.8 | 5.8 | 11.5 | 11.5 | 11.5 |  |  |
| West Asia & North Africa | 13 | 938 | 1.56E+05 | 15 | 64.4 | 67.6 | 58.6 | 13.2 | 18.6 | 26.9 | 108 | 283 | 74.15 | 4641 |
| No ETS | 5 | 224 | 1.32E+04 | 7 | 76.1 | 96.2 | 99.9 | 7.76 | 16 | 68.6 | 283 | 283 | 62.5 | 72.3 |
| Western Europe | 68 | 3722 | 6.55E+05 | 119 | 24 | 24.8 | 23.1 | 4 | 7.96 | 19 | 49 | 197 | 63.7 | 2120 |
| No ETS | 35 | 1578 | 2.09E+05 | 51 | 17.9 | 12.5 | 21.1 | 4.61 | 7 | 15.6 | 51 | 64 | 39.5 | 403 |

**Table 5.** Weighted statistics for means of PM1 and PM0.1-1(μg/m^3^) measurements (including calculated means) and statistics for maximum of PM1, and PM0.1-1(μg/m^3^) measurements. Statistics labeled ‘No ETS’ are restricted to measurements in homes without Environmental Tobacco Smoke. Studies may report means for multiple locations and times (n is the total number of means). Not all studies provided a mean or a maximum

| Region | Studies | Homes | Total Obs. Time (hr) | Statistics for Mean Measurements |  |  |  |  |  |  |  |  | Maximum |  |
| --- | --- | --- | --- | --- | --- | --- | --- | --- | --- | --- | --- | --- | --- | --- |
|  |  |  |  | n | Mean | SD | Wt. Mean | Min | Wt. 10% | Wt. 50% | Wt. 90% | Max | Median | Highest Value |
| All | 43 | 1280 | 6.63E+04 | 77 | 52.3 | 74.5 | 23.5 | 0.9 | 1 | 10.4 | 51 | 449 | 73.5 | 862 |
| No ETS | 12 | 171 | 1.08E+04 | 20 | 31.6 | 30.6 | 19 | 3 | 6.9 | 10.4 | 48.2 | 110 | 38.165 | 181 |
| Africa (except North) | 1 | 15 | 4.18E+01 | 2 | 11.5 | 3.99 | 11.3 | 8.65 | 8.65 | 8.65 | 14.3 | 14.3 |  |  |
| East Asia | 10 | 600 | 2.36E+03 | 16 | 101 | 135 | 40.1 | 6.1 | 25.9 | 33.4 | 48.2 | 449 | 141 | 153 |
| No ETS | 2 | 21 | 1.10E+03 | 3 | 63.1 | 41.5 | 44.6 | 31.2 | 31.2 | 48.2 | 48.2 | 110 |  |  |
| Eastern Europe | 3 | 52 | 5.31E+03 | 5 | 11.8 | 6.64 | 10.8 | 5.7 | 7.3 | 7.3 | 22.7 | 22.7 | 34.33 | 51.9 |
| No ETS | 1 | 1 | 2.38E+03 | 3 | 15.3 | 6.38 | 15.6 | 11.6 | 11.6 | 11.7 | 22.7 | 22.7 | 34.33 | 51.9 |
| North America | 5 | 115 | 1.94E+04 | 7 | 21.9 | 21.9 | 22.2 | 2.55 | 5.2 | 8.3 | 47 | 56.5 | 110 | 862 |
| No ETS | 1 | 59 | 1.44E+03 | 1 | 23.6 | NA | 23.6 | 23.6 | 23.6 | 23.6 | 23.6 | 23.6 |  |  |
| South Asia | 6 | 15 | 2.93E+03 | 16 | 90 | 42.9 | 106 | 31 | 99 | 103 | 135 | 157 | 76 | 260 |
| No ETS | 2 | 5 | 7.08E+02 | 5 | 56.8 | 27.4 | 87 | 34.9 | 35 | 101 | 101 | 101 | 76 | 181 |
| West Asia & North Africa | 2 | 5 | 4.90E+03 | 4 | 43.7 | 39.1 | 30.5 | 10.1 | 10.1 | 10.4 | 84.4 | 84.4 | 42 | 42 |
| No ETS | 1 | 1 | 3.46E+03 | 2 | 10.2 | 0.212 | 10.2 | 10.1 | 10.1 | 10.1 | 10.4 | 10.4 | 42 | 42 |
| Western Europe | 15 | 419 | 3.00E+04 | 26 | 20.7 | 17.3 | 13.4 | 0.9 | 0.9 | 8.1 | 37.9 | 63.4 | 75.7 | 601 |
| No ETS | 6 | 143 | 3.14E+03 | 7 | 13.3 | 13.1 | 7.78 | 3 | 3 | 6.9 | 14.7 | 41.6 | 16 | 22.4 |

**Table 6.** Weighted statistics for means of particle number - cutoffs unspecified (cm^−3^) measurements (including calculated means) and statistics for maximum of particle number - cutoffs unspecified (cm^−3^) measurements. Statistics labeled ‘No ETS’ are restricted to measurements in homes without Environmental Tobacco Smoke. Studies may report means for multiple locations and times (n is the total number of means). Not all studies provided a mean or a maximum.

| Region | Studies | Homes | Total Obs. Time (hr) | Statistics for Mean Measurements |  |  |  |  |  |  |  |  | Maximum |  |
| --- | --- | --- | --- | --- | --- | --- | --- | --- | --- | --- | --- | --- | --- | --- |
|  |  |  |  | n | Mean | SD | Wt. Mean | Min | Wt. 10% | Wt 50% | Wt 90% | Max | Median | Highest Value |
| **All** | 17 | 365 | 2.40E+04 | 26 | 15983 | 35708 | 14298 | 4.58 | 75.2 | 12083 | 20352 | 185211 | 103582.5 | 389076 |
| No ETS | 9 | 146 | 2.11E+04 | 14 | 5882 | 6218 | 12923 | 4.58 | 29.1 | 12083 | 20352 | 20352 | 17451 | 246000 |
| East Asia | 3 | 10 | 4.20E+02 | 4 | 50433 | 90182 | 62377 | 5.19 | 16400 | 16400 | 185211 | 185211 | 243.399 | 303247 |
| No ETS | 1 | 1 | 1.20E+01 | 2 | 60.3 | 77.9 | 60.3 | 5.19 | 5.19 | 60.3 | 115.34 | 115.34 | 128.03 | 243.40 |
| North America | 8 | 174 | 1.62E+04 | 10 | 5158 | 7202 | 13780 | 4.58 | 75.2 | 12083 | 20352 | 20352 | 203068.5 | 389076 |
| No ETS | 4 | 101 | 1.86E+04 | 7 | 7300 | 7743 | 13191 | 4.58 | 29.1 | 12083 | 20352 | 20352 | 10.7 | 10.7 |
| Oceania | 1 | 14 | 6.72E+02 | 1 | 10900 | NA | 10900 | 10900 | 10900 | 10900 | 10900 | 10900 | 22100 | 22100 |
| South East Asia | 1 | 6 | 9.68E+02 | 1 | 18366.1 | NA | 18366 | 18366 | 18366 | 18366 | 18366 | 18366 | 103165 | 103165 |
| Western Europe | 4 | 161 | 5.79E+03 | 10 | 13298 | 10483 | 15247 | 2250 | 3000 | 12715 | 26653 | 31816 | 104000 | 246000 |
| No ETS | 4 | 44 | 2.55E+03 | 5 | 6225 | 3887 | 7019 | 2250 | 2250 | 7948 | 11815 | 11815 | 89200 | 246000 |

**Table 7.** Weighted statistics for means of Ultra Fine Particles (UFP) (including calculated means) and statistics for maximum of UFP measurements. Statistics labeled ‘No ETS’ are restricted to measurements in homes without Environmental Tobacco Smoke. Studies may report means for multiple locations and times (n is the total number of means). Not all studies provided a mean or a maximum.

| PM species and unit | Studies | Homes | Total Obs. Time (hr) | Statistics for Mean Measurements |  |  |  |  |  |  |  |  | Maximum |  |
| --- | --- | --- | --- | --- | --- | --- | --- | --- | --- | --- | --- | --- | --- | --- |
|  |  |  |  | n | Mean | SD | Wt. Mean | Min | Wt. 10% | Wt 50% | Wt 90% | Max | Median | Highest Value |
| UFP, PM<sub>0.1</sub>, PM<sub>0.25</sub> and PM<sub>0.5</sub> (µg/m<sup>3</sup>) | 12 | 228 | 3.08E+04 | 31 | 18.2 | 17 | 14.1 | 2.2 | 2.8 | 8.99 | 37.6 | 68.2 | 136.5 | 143.9 |
| UFP, PM<sub>0.1</sub> and PM<sub>0.25</sub> (µg/m<sup>3</sup>) | 10 | 208 | 2.94E+04 | 19 | 15.1 | 15.5 | 10.4 | 2.2 | 2.2 | 8.93 | 19.8 | 54.5 | 103.18 | 136.5 |
| UFP, PM<sub>0.1</sub> and PM<sub>0.25</sub> (cm<sup>-3</sup>) | 31 | 1 074 | 6.92E+04 | 54 | 17154 | 18648 | 15608 | 2.44E+03 | 7990 | 14000 | 26000 | 1.19E+05 | 69640 | 3.80E+06 |

**Table 8.** Coefficients mean, standard error, and significance for log-normal weighted regression of mean concentrations of indoor PM2.5 (and PM0.25-2.5) over time: Indoor ∼ Intercept + b1 * Weight *(Sampling midpoint – 1980). Additional parameters added to pooled models. Concentrations in mg/m^3^. R^2^ is the pseudo coefficient of determination, along with relative distortion (dist.); FK = Fligner-Killeen non-parametric test on homogeneity of residuals variances. Only results from locations with at least 4 different studies are presented. Significance codes: *** <1E-3; **<1E-2; *<5E-2; .<1E-1

| Location | Intercept | Year since 1980 ($\beta_1$) | Earliest estimate | Latest estimate | n of means | n of studies | $R^2_d$ dist. | Pooled models |
| --- | --- | --- | --- | --- | --- | --- | --- | --- |
| Canada | 25.6 ± 8.7<br>7.5E-3 ** | -0.55 ± 0.28<br>6.3E-2 . | 16.7<br>(1996) | 5.2<br>(2016) | 24 | 15 | 0.16<br>-0.5% | FK p-value = 0.1083<br>$R^2_d$ = 0.23, dist. = 5.0%<br>Intercept: 21.6 ± 2.8; p = 7E-13 ***<br>Year: -0.35 ± 0.09; p = 3.0E-4 ***<br>No ETS: - 3.4 ± 0.9; p = 3.9E-4 ***<br>Air cleaners: - 4.5 ± 1.2; p = 4.0E-4 ***<br>USA: +4.3 ± 1.1; p = 6.6E-5 *** |
| USA | 27.1 ± 3.1<br>3.2E-15<br>*** | -0.43 ± 0.11<br>1.0E-4 *** | 24.1<br>(1987) | 10.2<br>(2019) | 180 | 87 | 0.08<br>0.9% | |
| Baltimore | 124 ± 39<br>9.6E-3 *** | -3.7 ± 1.3<br>1.9E-2 ** | 55.1<br>(1998) | 12.9<br>(2010) | 13 | 6 | 0.62<br>14% | |
| Boston | 32.8 ± 6.0<br>8.4E-5 *** | -0.71 ± 0.19<br>2.4E-3 ** | 21.3<br>(1996) | 8.9<br>(2013) | 17 | 11 | 0.54<br>2.8% | |
| Detroit | 53.4 ± 13.3<br>1.0E-2 * | -1.2 ± 0.43<br>3.9E-2 * | 28.9<br>(2000) | 10.8<br>(2015) | 8 | 5 | 0.53<br>0.9% | |
| Los Angeles | 54.0 ± 12.8<br>1.8E-3 ** | -1.7 ± 0.50<br>5.5E-3 ** | 20.8<br>(1998) | 6.6<br>(2007) | 13 | 4 | 0.64<br>3.4% | |
| New York | 33.4 ± 7.0<br>3.1E-4 *** | -0.53 ± 0.24<br>4.3E-2 * | 23.3<br>(1999) | 14.8<br>(2015) | 17 | 8 | 0.23<br>-3.0% | |
| China | 112 ± 32<br>6.6E-4 *** | -1.9 ± 0.88<br>3.1E-2 * | 98.1<br>(1987) | 37.8<br>(2018) | 118 | 55 | 0.06<br>0.8% | Pooled model also includes data from Japan<br>n of means = 142; n of studies = 67<br>FK p-value = 0.1016 |
| | | | | | | | | $R^2_d = 0.53$, dist. = -0.8% |
| **Taiwan** | 64.2 ± 19.4<br>3.8E-3 ** | -1.4 ± 0.24<br>2.7E-2 * | 45.9<br>(1993) | 14.6<br>(2015) | 21 | 9 | 0.27<br>-1.2% | Intercept: 119 ± 31; p = 2.1E-4 ***<br>Year: -2.09 ± 0.86; p = 1.7E-2 *<br>No ETS: - 12.8 ± 4.0; p 1.9E-3 **<br>Air cleaners: - 6.2 ± 2.2; p 7.0E-3 **<br>Japan : -70.7 ± 35.7; p 4.9E-2*<br>Taiwan: -72.1 ± 36.9; p 5.2E-2.<br>Japan x Year: 1.4 ± 1.0 p = 0.18<br>Taiwan x Year: 1.6 ± 1.1 p = 0.13 |
| **South Korea** | -149 ± 39.3<br>5.3E-3 ** | +5.2 ± 1.2<br>2.0E-3 ** | 8.2<br>(2010) | 40.0<br>(2016) | 11 | 7 | 0.51<br>4.3% |  |
| *Beijing* | 212 ± 47<br>2.6E-4 *** | -4.8 ± 1.3<br>1.9E-3 ** | 178<br>(1987) | 33.2<br>(2017) | 22 | 15 | 0.64<br>1.1% |  |
| *Guangzhou* | -276 ± 244<br>0.30 | +14.2 ± 10<br>0.21 | 57.5<br>(2003) | 257<br>(2017) | 9 | 4 | 0.40<br>1.9% |  |
| *Hong Kong* | 34.2 ± 17.3<br>6.4E-2 . | -0.07 ±<br>0.53<br>0.89 | 32.7<br>(1998) | 31.2<br>(2018) | 21 | 9 | 0.00<br>3E-5 |  |
| *Seoul* | -122 ± 72<br>0.15 | +4.5 ± 2.10<br>0.89 | 13.8<br>(2010) | 38.9<br>(2016) | 8 | 6 | 0.33<br>-0.2% |  |
| *Shanghai* | 187 ± 187<br>0.35 | -4.4 ± 5.1<br>0.41 | 36.7<br>(2014) | 21.9<br>(2017) | 10 | 7 | 0.10<br>0.4% |  |
| *Taipei* | 60.4 ± 22.0<br>1.9E-2 * | -1.3 ± 0.66<br>7.7E-2 . | 43.6<br>(1993) | 15.7<br>(2014) | 14 | 7 | 0.27<br>-1.5% |  |
| *Tianjin* | 1755 ±<br>1087<br>0.21 | -45.4 ±<br>28.8<br>0.21 | 209<br>(2014) | 43.0<br>(2017) | 6 | 4 | 0.51<br>0.7% |  |
| *Xi'an* | 109 ± 96<br>0.37 | -1.1 ± 2.6<br>0.73 | 74.7<br>(2011) | 68.5<br>(2017) | 5 | 4 | 0.05<br>3E-4 |  |
| **France** | 76.7 ± 26.9<br>2.2E-2 * | -2.0 ± 0.84<br>4.1E-2 * | 61.9<br>(1992) | 13.0<br>(2014) | 11 | 5 | 0.39<br>-1.0% | Pooled model also includes data from Belgium, Denmark, Germany, Ireland, Switzerland<br>n of means = 109; n of studies = 63<br>FK p-value = 0.05062<br>R<sup>2</sup><sub>d</sub> = 0.47, dist. = 5.8%<br>Intercept: 43.8 ± 3.8; p < 2E-16 ***<br>Year: -0.94 ± 0.14; p = 3.4E-10 ***<br>No ETS: - 3.0 ± 2.0; p 0.12<br>Air cleaners: - 6.8 ± 2.0; p 7.0E-4 *** |
| **Greece** | 41.2.6 ± 6.7<br>1.7E-4 *** | -0.78 ± 0.20<br>4.5E-7 *** | 26.7<br>(1998) | 12.5<br>(2016) | 12 | 7 | 0.65<br>-1.8% |  |
| **Italy** | 87.6 ± 5.9<br>1.2E-8 *** | -2.1 ± 0.21<br>4.5E-7 *** | 61.9<br>(1992) | 13.0<br>(2014) | 14 | 7 | 0.87<br>-0.7% |  |
| **Netherlands** | 37.3 ± 7.9<br>4.2E-2 * | -0.71 ± 0.26<br>0.11 | 26.4<br>(1995) | 11.7<br>(2015) | 5 | 4 | 0.67<br>-2.2% |  |
| **Norway** | 25.1 ± 11.4<br>0.16 | -0.35 ± 0.37<br>0.44 | 19.9<br>(1994) | 13.4<br>(2013) | 5 | 4 | 0.27<br>-2.2% |  |
| **Portugal** | 21.0 ± 151<br>0.89 | +0.73 ± 4.8<br>0.88 | 41.4<br>(2008) | 45.9<br>(2014) | 11 | 6 | 0.00<br>0.0% |  |
| **Sweden** | -12.2 ± 51.2<br>0.83 | +0.94 ± 2.1<br>0.70 | 9.6<br>(2003) | 13.4<br>(2006) | 5 | 4 | 0.06<br>-3.0% |  |
| **UK** | 27.8 ± 5.8<br>8.3E-5 *** | -0.35 ± 0.37<br>0.14 | 22.3<br>(1995) | 14.3<br>(2017) | 25 | 13 | 0.08<br>1.6% |  |
| **Finland** | 19.0 ± 2.9<br>1.2E-3 ** | -0.42 ± 0.11<br>1.3E-2 * | 11.3<br>(1998) | 5.4<br>(2012) | 8 | 6 | 0.61<br>-6.4% |  |
| *Athens* | 46.7 ± 5.0<br>1.3E-5 *** | -1.0 ± 0.15<br>1.5E-4 *** | 28.1<br>(1998) | 10.7<br>(2015) | 11 | 7 | 0.85<br>0.4% |  |
| *Oslo* | 25.1 ± 11.4<br>0.16 | -0.35 ± 0.37<br>0.44 | 19.9<br>(1994) | 13.4<br>(2013) | 5 | 4 | 0.26<br>-2.2% |  |
| *Oporto* | -53 ± 207<br>0.81 | +3.53 ± 6.7<br>0.64 | 45.6<br>(2008) | 63.2<br>(2013) | 6 | 4 | 0.06<br>-0.6% |  |
| **Czech Republic** | 47.0 ± 23.6<br>0.12 | -1.16 ± 0.87<br>0.25 | 25.5<br>(1998) | 8.8<br>(2012) | 7 | 4 | 0.23<br>-11% |  |
| *Prague* | 46.8 ± 26.8<br>0.18 | -1.11 ± 1.00<br>0.35 | 26.3<br>(1998) | 10.4<br>(2012) | 6 | 4 | 0.20<br>-9.3% |  |
| **Chile** | 239 ± 56<br>5.2E-3 ** | -5.6 ± 1.49<br>9.4E-3 ** | 129<br>(1999) | 27.7<br>(2017) | 9 | 6 | 0.89<br>7.4% |  |
| **Singapore** | 36.4 ± 37.3<br>0.36 | -0.60 ± 1.14<br>0.61 | 21.7<br>(2004) | 14.8<br>(2015) | 10 | 4 | 0.04<br>-1.5% |  |
| **India** | 224 ± 90<br>2.4E-2 * | -3.5 ± 2.9<br>0.23 | 156<br>(1999) | 92.3<br>(2017) | 19 | 11 | 0.09<br>0.1% |  |
| **Pakistan** | -655 ± 415<br>0.15 | +29 ± 14<br>7.5E-2 . | 154<br>(2007) | 337<br>(2014) | 10 | 4 | 0.34<br>0.9% |  |
| *Lahore* | -480 ± 710<br>0.52 | +24 ± 23<br>0.33 | 195<br>(2008) | 327<br>(2014) | 9 | 4 | 0.11<br>0.4% |  |
| **Israel** | 927 ± 458<br>0.13 | -24.1 ± 12.4<br>0.15 | 139<br>(2012) | 22.8<br>(2017) | 6 | 4 | 0.40<br>7.7% |  |

**Table 9.** Coefficients mean, standard error, and significance for log-normal weighted regression of mean concentrations of indoor PM10 (and PM1-10) over time: Indoor ∼ Intercept + β1 * Weight *(Sampling midpoint – 1980). Additional parameters added to pooled models. Concentrations in μg/m^3^. R^2^ is the pseudo coefficient of determination, along with relative distortion (dist.); FK = Fligner-Killeen non-parametric test on homogeneity of residuals variances. Only results from locations with at least 4 different studies are presented. Significance codes: *** <1E-3; **<1E-2; *<5E-2; .<1E-1

| Location | Intercept | Year since 1980 | Earliest estimate | Latest estimate | n of means | n of studies | $R^2_d$ dist. | Pooled models |
| --- | --- | --- | --- | --- | --- | --- | --- | --- |
| USA | 45.7±11.2<br>1.4E-4*** | -1.05±0.43<br>1.7E-2* | 37.4<br>(1987) | 9.7<br>(2014) | 58 | 31 | 0.10<br>-2.5% |  |
| Baltimore | -136±49.4<br>0.2E-2 | 8.18±2.3<br>1.7E-1 | 15.9<br>(1998) | 68.7<br>(2005) | 4 | 4 | 0.92<br>23.7% |  |
| China | 101±33.4<br>4.5E-3*** | -0.13±1.05<br>0.90 | 99.9<br>(1987) | 95.7<br>(2018) | 40 | 20 | 0.00<br>-0.0% | n of means = 64; n of studies = 32<br>FK p-value =0.1111<br>$R^2_d = 0.71$, dist. = -0.4%<br>Intercept: $98.9 \pm 31.3$; $p = 2.6\text{E-}3$ **<br>Year: $0.01 \pm 1.0$; $p = 0.98$<br>No ETS: $-6.3 \pm 9.4$; $p = 0.50$<br>Air cleaners: $-56.5 \pm 24.5$; $p = 2.5\text{E-}2$ *<br>Japan: $-122 \pm 89.5$; $p = 0.17$<br>Taiwan: $30.3 \pm 61.0$; $p = 0.62$<br>Japan x Year: $3.2 \pm 5.1$ $p = 0.53$<br>Taiwan x Year: $-3.1 \pm 2.0$ $p = 0.14$ |
| Japan | -50.0±67.2<br>.51 | 4.46±4.21<br>.37 | 16.4<br>(1994) | 104.0<br>(2014) | 6 | 4 | 0.26<br>-7.0% |  |
| Taiwan | 137±49.9<br>0.1E-1* | -3.5±1.5<br>3.7E-2* | 91.4<br>(1993) | 13.2<br>(2015) | 18 | 8 | 0.41<br>-1.1% |  |
| South Korea | -25.7±40.2<br>5.3E-1 | 2.5±1.25<br>6.9E-2. | 34.5<br>(2004) | 63.0<br>(2015) | 15 | 9 | 0.20<br>-1.2% |  |
| Hong Kong | 86±37<br>4.4E-2* | -1.7<br>2.8E-1 | 57.7<br>(1996) | 21.7<br>(2018) | 12 | 6 | 0.06<br>-0.6% |  |
| Seoul | -22±69<br>7.7E-1 | 2.4±2.1<br>0.3 | 46.5<br>(2008) | 63.0<br>(2015) | 9 | 6 | 0.14<br>-1.3% |  |
| Taipei | 128±69<br>1.1E-1 | -3.2±2.3<br>1.9E-1 | 86.3<br>(1993) | 24.8<br>(2012) | 10 | 6 | 0.35<br>-0.8% |  |
| Portugal | -129±102 | 5.7±3.3 | 34.1 | 66.0 | 17 | 6 | 0.15 |  |
|  | 0.23 | 0.10 | (2008) | (2014) |  |  | -0.6% |  |
| **Greece** | 56.8±17.6<br>1.8E-2* | -0.88±0.62<br>0.21 | 37.0<br>(2002) | 25.3<br>(2015) | 9 | 5 | 0.20<br>2.6% | Pooled model also includes data from Finland, France, Germany, Netherlands, Norway, and Sweden<br>n of means = 56; n of studies = 31<br>FK p-value = 0.05062<br>R<sup>2</sup><sub>d</sub> = 0.81, dist. = -0.1%<br>Intercept: -11.7 ± 45.7; p = 0.80<br>Year: 0.88 ± 1.48; p = 0.55<br>No ETS: -3.3 ± 11.2; p = 0.77<br>France: 153 ± 50; p = 3.7E-3 **<br>Sweden: -859 ± 269; p = 2.9E-3 **<br>France x Year: -4.4 ± 1.6; p = 9.5E-3 **<br>Sweden x Year: 29.5 ± 9.1; p = 2.6E-3 **<br>All other country and interaction terms not significant at 0.05 level. |
| **Italy** | 95.6 ±<br>23.7<br>5.6E-2 · | -2.1 ± 0.6<br>8.5E-2 · | 38.9<br>(2007) | 18.3<br>(2017) | 5 | 4 | 0.85<br>0.3% |  |
| **United Kingdom** | 42.2 ±<br>10.3<br>7.9E-4*** | -0.50 ±<br>0.51<br>0.34 | 34.5<br>(1995) | 29.1<br>(2006) | 20 | 6 | 0.05<br>-0.2% |  |
| *Athens* | 56.8 ±<br>17.6<br>1.8E-2* | -0.9 ± 0.6<br>0.21 | 37.0<br>(2002) | 25.3<br>(2015) | 9 | 5 | 0.20<br>2.6% |  |
| **India** | 389 ± 141<br>1.9E-2 | -6.3 ± 4.9<br>0.23 | 267<br>(1999) | 154<br>(2017) | 14 | 10 | 0.13<br>0.5% |  |
| *Delhi* | 552 ±<br>1018<br>0.68 | -14.8 ±<br>49.3<br>0.81 | 267<br>(1999) | 186<br>(2004) | 4 | 4 | 0.03<br>-1.7% |  |
| **Thailand** | 126 ± 85<br>0.24 | -2.8 ± 2.3<br>0.31 | 80.6<br>(1996) | 19.9<br>(2017) | 6 | 4 | 0.54<br>6.2% |  |

**Table 10.** Coefficients mean, standard error, and significance for log-normal weighted regression of mean concentrations of indoor PM1 (and PM0.1-1) over time: Indoor ∼ Intercept + β1 * Weight *(Sampling midpoint – 1980). Concentrations in μg/m^3^. R^2^ is the pseudo coefficient of determination, along with relative distortion (dist.). Only results from locations with at least 4 different studies are presented. Significance codes: *** <1E-3 **<1E-2 *<5E-2 .<1E-1

| Location | Intercept | Year since 1980 | Earliest estimate | Latest estimate | n of means | n of studies | $R^2_d$ dist. |
| --- | --- | --- | --- | --- | --- | --- | --- |
| **Western Europe** | 24.4 ± 12.6<br>6.5E-2 . | -0.63 ± 0.38<br>0.11 | 12.7<br>(1998) | 1.6<br>(2015) | 26 | 15 | 0.11<br>0.1% |
| **East Asia** | 13.5 ± 21.0<br>0.53 | 0.68 ± 0.66<br>0.32 | 27.1<br>(1999) | 38.0<br>(2015) | 16 | 9 | 0.06<br>0.3% |
| *China* | 213 ± 327<br>0.54 | -4.9 ± 9.2<br>0.61 | 55.6<br>(2011) | 36.2<br>(2015) | 10 | 5 | 0.05<br>-1.6% |
| *Taiwan* | 8.5 ± 16.9<br>0.65 | 0.88 ± 0.69<br>0.29 | 25.9<br>(1999) | 36.6<br>(2012) | 6 | 4 | 0.30<br>0.1% |
| **North America** | -150 ± 105<br>0.22 | 5.4 ± 3.5<br>0.20 | 2.0<br>(2008) | 46.1<br>(2016) | 7 | 5 | 0.09<br>-13% |
| **South Asia** | 302 ± 48<br>2.7E-5 *** | - 6.9 ± 1.6<br>7.1E-4 *** | 144<br>(2003) | 42.9<br>(2017) | 16 | 6 | 0.41<br>0.1% |

**Table 11.**
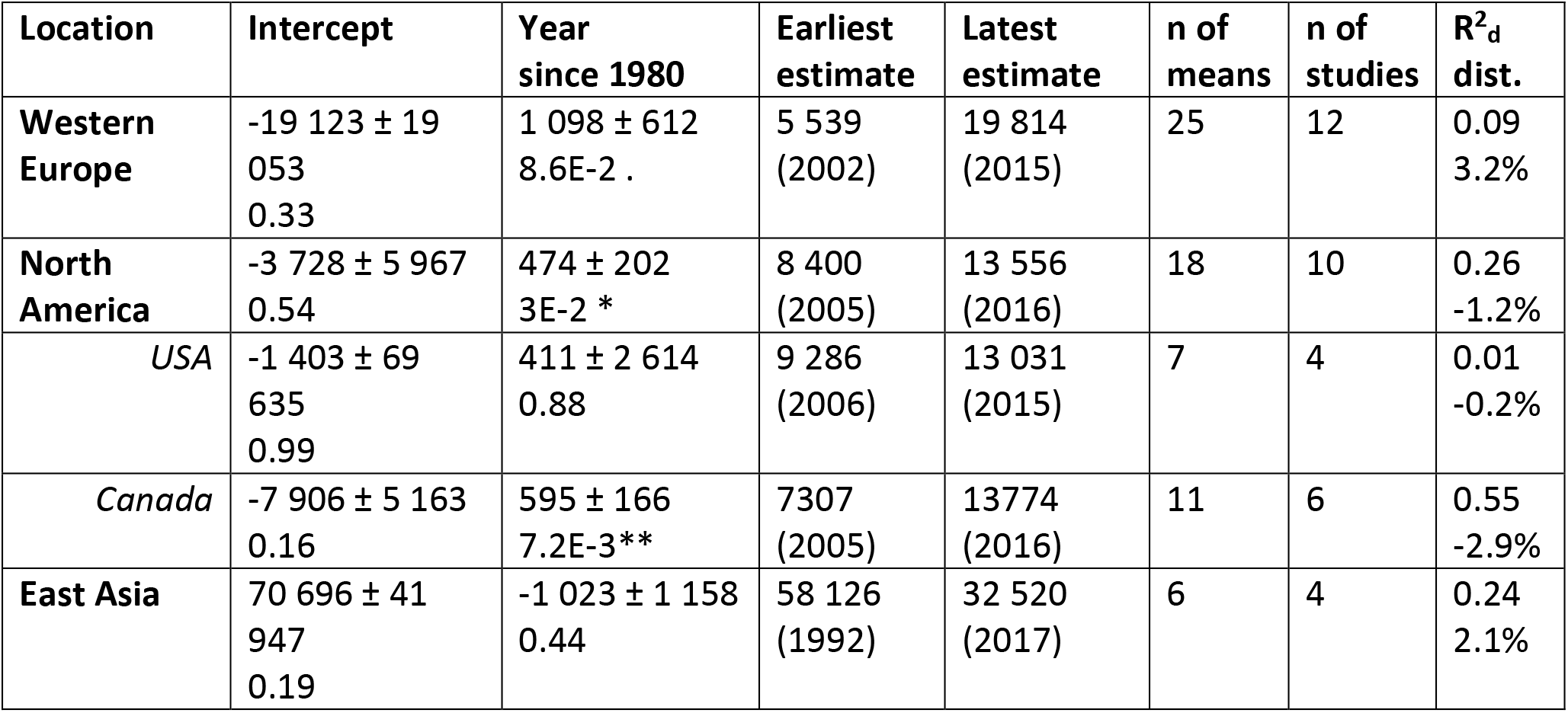
Coefficients mean, standard error, and significance for log-normal weighted regression of mean concentrations of indoor UFP (and PM0.25 and PM0.1) over time: Indoor ∼ Intercept + β1 * Weight *(Sampling midpoint – 1980). Concentrations in cm^-3^. R^2^ is the pseudo coefficient of determination, along with relative distortion (dist.); s = standard deviation of model. Only results from locations with at least 4 different studies are presented. Significance codes: *** <1E-3; **<1E-2; *<5E-2; .<1E-1

| Location | Intercept | Year since 1980 | Earliest estimate | Latest estimate | n of means | n of studies | $R^2_d$ dist. |
| --- | --- | --- | --- | --- | --- | --- | --- |
| **Western Europe** | -19 123 ± 19 053<br>0.33 | 1 098 ± 612<br>8.6E-2 . | 5 539<br>(2002) | 19 814<br>(2015) | 25 | 12 | 0.09<br>3.2% |
| **North America** | -3 728 ± 5 967<br>0.54 | 474 ± 202<br>3E-2 * | 8 400<br>(2005) | 13 556<br>(2016) | 18 | 10 | 0.26<br>-1.2% |
| USA | -1 403 ± 69 635<br>0.99 | 411 ± 2 614<br>0.88 | 9 286<br>(2006) | 13 031<br>(2015) | 7 | 4 | 0.01<br>-0.2% |
| Canada | -7 906 ± 5 163<br>0.16 | 595 ± 166<br>7.2E-3** | 7307<br>(2005) | 13774<br>(2016) | 11 | 6 | 0.55<br>-2.9% |
| **East Asia** | 70 696 ± 41 947<br>0.19 | -1 023 ± 1 158<br>0.44 | 58 126<br>(1992) | 32 520<br>(2017) | 6 | 4 | 0.24<br>2.1% |

It is tempting to compare weighted mean values of indoor PM concentrations to regulatory limits set for PM in ambient air, which are more often available than for indoor air. However such comparisons are generally inappropriate and can be misleading. This is primarily because different national or international standards may be set weighing economic or practical considerations, in addition to evidence of health effects. Even when entirely health-based, the evidence on which they are built does not necessarily account for the reduction in exposure to ambient PM provided by buildings, nor for the variability of exposure to indoor-generated particles. Consequently, indoor exposures at levels below outdoor limits should not automatically be regarded as safe. One international set of health-based guidelines specifically developed to be applicable to indoor environments are the World Health Organization’s AQG levels (2021 update), recommending PM_10_ levels below 15 μg/m^3^ (annual) and 45 μg/m^3^ (daily); and PM_2.5_ levels below 5 μg/m^3^ (annual) and 15 μg/m^3^ (daily). In this analysis, all regions (Table 2) had weighted PM_10_ means well above 15 μg/m^3^, and several also above 45 μg/m^3^, ranging from 30 μg/m^3^ for North America to more than 200 μg/m^3^ for South Asia. Similarly, all regions (Table 4) registered weighted PM_2.5_ means well in excess of 5 μg/m^3^, and all regions except Oceania also above the daily 15 μg/m^3^ level, from 16.8 μg/m^3^ for North America to 162 μg/m^3^ for South Asia. As already remarked, the studies in this survey are not population representative, but if mean (and median) values generally exceed these guidelines, it would be reasonable to expect that, over the past decades, an important fraction of the population has been exposed indoors to levels of PM that are of concern for health, even in the absence of solid fuels or ETS.

#### 3.3.2. Long-term trends

Indoor PM concentrations have been measured over the years through *ad-hoc* studies (Figure 7), so that real time series for specific locations are not available and time trends estimates inevitably confound some spatial variability within a region, country, or city. Yet, where results are consistent at different scales, they may still reveal real time trends. To understand if and how indoor PM concentrations have changed over the decades of the survey, we can consider the regression results in Tables 8-11. Where few studies were available, regression parameters are of limited inferential value and the most informative elements reported in the table are the earliest and latest estimates, offering a relative comparison of measurements and their time span. No generally applicable conclusion can be drawn about the time course of indoor concentrations, but a few regional and local trends stand out. Concentrations of indoor PM_2.5_ in North America, and most clearly in the United States, have been decreasing at a rate of about - 0.3 to -0.5 μg/m^3^ per year. Concentrations of PM_10_ have been trending down as well, at a rate of −1.0 ± 0.4 μg/m^3^ per year. This pattern (for PM_2.5_) is also clearly seen more locally, in several US cities with sufficient available data. For the smaller size fractions, UFP number concentrations have shown a small increase from the earliest measurements in 2005, though it is significant only for Canada.

For East Asian locations, evidence of trends is mixed and more limited. While PM_2.5_ concentrations show statistically significant decreasing patterns for China, Taiwan, Beijing, and marginally significant for Taipei, no trends are significant for other individual East Asian cities, or for Japan, which can reflect a real lack of trend, lack of data, or both. Concentrations for Hong Kong, a city with relatively abundant data, are decidedly flat over time. South Korean measurements shows a significant increase in PM_2.5_ and a marginally significant increase in PM_10_ (from a larger set of studies). The decreases for China and Beijing, however, strongly reflect the high concentrations in the early (1987) measurements of a single smaller study^46^. No trends are significant for other size fractions in the region, except for a PM_10_ decrease in Taiwan, at a rate of −3.5 ± 1.5 μg/m^3^ per year.

Countries and cities in Western and Eastern Europe also display generally decreasing trends of indoor PM_2.5_ concentrations, but significantly so only for Finland, France, Greece, Italy, and the city of Athens. For the UK, the relatively numerous studies suggest no significant trend over time. The lack of significant trends elsewhere must be considered in light of the administrative fragmentation, leading to few studies per jurisdiction. The pooled regression for Western Europe (excluding Spain and Finland, whose residuals variances were too different from the rest) shows a highly significant decreasing trend of −0.94 ± 0.14 μg/m^3^ per year (p-value of coefficient: 3E-10). Trends for indoor PM_10_ and PM_1_ were mixed and not significant for any location, including for the pooled regressions. A marginally significant increase may be noted for UFP, like for North America.

Information on indoor concentrations in other regions was generally too sparse to produce informative results on their long-term changes, with few exceptions. Statistically significant decreases in PM_2.5_ concentrations can be observed for Chile (−5.6 ± 1.5 μg/m^3^ per year) and Bangladesh (−11 ± 1 μg/m^3^ per year), and a significant increase for Mexico (2.1 ± 0.9 μg/m^3^ per year), though the latter two estimates rely on only 2 and 3 studies respectively (data not shown in the table). Comparatively more numerous data were available for Singapore, India, and Pakistan, where no significant trends are observed. South Asia is the only region showing a significant decrease in PM_1_ concentrations, though this may simply reflect a tendency towards the mean from the very high levels reported in the earliest measurements.

Overall, results are consistent with generally stable indoor concentrations for the three decades of this survey, with specific regional exceptions for PM_2.5_ and PM_10_. Understanding all the factors contributing to these observed differences is beyond the scope of this survey, but these localized decreases may reflect well-documented declines in prevalence of smoking^47^ and ambient air concentrations of regulated pollutants^48, 49^. Even in a survey of this size, the limited availability of indoor PM measurements repeated in the same areas limits further exploration of these temporal trends.

### 3.4. Factors affecting indoor concentrations

The importance of a long-term trend in explaining the observed variability in indoor PM concentrations varies for different locations, but is generally low for locations with richer data sets (e.g., USA, China, UK, Canada), as can be seen from the R^2^_d_ values in Tables 8-11, ranging as low as 0.00 to 0.16. The apparent influence of time trend on observed variability increases when few observations are available at a given location, but this is really only an indication of limited number of indoor PM measurements. Even when time trends are significant, however, the evolution of indoor PM concentrations must be traced to direct causes that also change over time. Here we explored outdoor PM concentrations and indoor environmental tobacco smoke (ETS) in some detail. Any effects of climate and seasonality, also related to time, will be explored in future work.

#### 3.4.1. Outdoor air concentrations

About half (198/428) of the studies in the survey reported paired indoor-outdoor (on site) measurements, and another 45 reported paired concentrations measured at off-site ambient monitors, usually regulatory monitors. These pairs of indoor-outdoor concentrations were remarkably well correlated across studies, as can be seen in Figure 8 for PM_2.5_, demonstrating the important role of outdoor air in residential indoor air quality. The results of lognormal regressions on these paired concentrations, based on equation (3), are reported in Tables 12 through 17.

**Table 12.** Coefficients mean, standard error, and significance for log-normal weighted regression of mean concentrations of indoor PM2.5 (and PM0.25-2.5) with respect to concentrations measured outdoor (on site): Indoor ∼ Intercept + β1 * Weight *Outdoor + β2 * ETS excluded (True or False). Concentrations in μg/m^3^. Weight is total sampling time. Only results from locations with at least 4 different studies are presented. Significance codes: *** <1E-3; **<1E-2; *<5E-2; .<1E-1

| Location | Intercept | Outdoor ($\beta_1$) | ETS excluded ($\beta_2$) | $R^2_d$ dist. | n of means pairs | n of studies |
| --- | --- | --- | --- | --- | --- | --- |
| Global model | 4.1 ± 0.6<br>2.3E-10 *** | 0.86 ± 0.05<br><2.0E-16 *** | -1.6 ± 0.5<br>1.9E-3 ** | 0.72<br>8.2% | 389 | 149 |
| Global model, only outdoor concentrations $\leq 50$ | 3.2 ± 0.8<br>8.4E-5 *** | 0.95 ± 0.06<br><2.0E-16 *** | -1.7 ± 0.6<br>4.1E-3 ** | 0.51<br>7.8% | 290 | 97 |
| Global model, only outdoor concentrations $\leq 25$ | 2.0 ± 0.9<br>3.2E-2 * | 1.07 ± 0.08<br><2.0E-16 *** | -1.5 ± 0.6<br>1.9E-3 ** | 0.47<br>6.6% | 219 | 81 |
| Global model, only pairs excluding ETS | 3.8 ± 1.1<br>4.2E-4 *** | 0.75 ± 0.08<br>2.4E-16 *** | | 0.55<br>2.4% | 150 | 59 |
| North America | 2.6 ± 1.0<br>1.4E-2 * | 1.04 ± 0.09<br><2.0E-16 *** | -2.7 ± 0.6<br>4.5E-3 ** | 0.43<br>5.1% | 149 | 48 |
| *Canada* | 1.8 ± 1.4<br>0.19 | 0.87 ± 0.16<br>2.0E-5 *** | -3.2 ± 1.1<br>7.7E-3 ** | 0.55<br>-2.9% | 27 | 9 |
| *USA* | 7.0 ± 1.6<br>3.2E-5 *** | 0.79 ± 0.13<br>1.1E-8 *** | -2.8 ± 0.9<br>2.7E-3 ** | 0.30<br>2.5% | 122 | 39 |
| East Asia | 14.3 ± 2.8<br>2.2E-6 *** | 0.48 ± 0.04<br><2.0E-16 *** | -8.0 ± 2.5<br>2.0E-3 ** | 0.86<br>9.1% | 86 | 31 |
| *China* | 19.8 ± 3.8<br>2.1E-6 *** | 0.37 ± 0.06<br>6.5E-9 *** | 6.9 ± 6.2<br>0.26 | 0.58<br>2.7% | 68 | 23 |
| *Taiwan* | 26.2 ± 3.8<br>3.E-5 *** | 0.31 ± 0.03<br>6.5E-5 *** | 3.9 ± 2.2<br>0.14 | 0.96<br>0.1% | 9 | 4 |
| Western Europe | 5.8 ± 1.2<br>1.9E-5 *** | 0.64 ± 0.08<br>8.1E-12 *** | -2.9 ± 0.8<br>1.2E-3 ** | 0.70<br>1.1% | 60 | 26 |
| *Finland* | 9.9 ± 5.3<br>0.20 | 0.07 ± 0.45<br>0.89 | -2.3 ± 2.4<br>0.45 | 0.25<br>0.2% | 6 | 4 |
| *Greece* | 9.4 ± 6.0<br>0.18 | 0.52 ± 0.23<br>0.07 . | -4.6 ± 3.8<br>0.27 | 0.51<br>-0.4% | 9 | 5 |
| *Italy* | 5.4 ± 6.9<br>0.46 | 0.67 ± 0.39<br>0.13 | -0.3 ± 10.6<br>0.98 | 0.29<br>-2.6% | 12 | 5 |
| *UK* | 11.2 ± 4.2<br>3.1E-2 * | 0.39 ± 0.23<br>0.13 | -1.3 ± 4.6<br>0.79 | 0.32<br>-1.8% | 11 | 5 |
| Eastern Europe | 0.4 ± 2.3<br>0.87 | 0.58 ± 0.14<br>4.0E-4 *** | 17.7 ± 6.0<br>8.3E-3 ** | 0.61<br>-1.0% | 24 | 7 |
| West Asia & North Africa | 2.3 ± 3.6<br>0.53 | 1.09 ± 0.14<br>5.9E-7 *** | -16.8 ± 2.3<br>9.8E-7 *** | 0.98<br>5.1% | 22 | 7 |
| South America | 6.1 ± 5.0<br>0.25 | 0.93 ± 0.13<br>5.3E-6 *** | 4.4 ± 3.9<br>0.27 | 0.90<br>0.3% | 18 | 6 |
| *Chile* | 48.8 ± 17.0<br>2.1E-2 * | 0.27 ± 0.28<br>0.35 | -6.4 ± 8.5<br>0.46 | 0.14<br>0.5% | 12 | 4 |
| Central America & Caribbean | 27.1 ± 5.4<br>4.9E-4 *** | 0.50 ± 0.11<br>1.4E-3 ** | -12.1 ± 3.6<br>6.8E-3 ** | 0.80<br>0.5% | 14 | 2 |

**Table 13.** Coefficients mean, standard error, and significance for log-normal weighted regression of mean concentrations of indoor PM2.5 (and PM0.25-2.5) with respect to concentrations measured at nearest ambient air monitor (not on site): Indoor ∼ Intercept + β1 * Weight *Ambient + β2 * ETS excluded (True or False). Concentrations in μg/m^3^. Weight is total sampling time. Only results from locations with at least 4 different studies are presented. Significance codes: *** <1E-3; **<1E-2; *<5E-2; .<1E-1

| Location | Intercept | Ambient (not on site) ($\beta_1$) | ETS excluded ($\beta_2$) | R<sup>2</sup><sub>d</sub> dist. | n of means pairs | n of studies |
| --- | --- | --- | --- | --- | --- | --- |
| Global model | 6.1 ± 1.3<br>2.1E-5 *** | 0.49 ± 0.07<br>3.0E-9 *** | -0.01 ± 1.3<br>0.995 | 0.56<br>7.3% | 78 | 36 |
| Global model, only pairs excluding ETS | 7.7 ± 1.5<br>2.0E-5 *** | 0.41 ± 0.11<br>5.6E-4 *** |  | 0.37<br>12.0% | 46 | 22 |
| North America | 0.3 ± 3.1<br>0.92 | 1.13 ± 0.31<br>1.2E-3 ** | 0.2 ± 1.7<br>0.93 | 0.46<br>1.9% | 30 | 16 |
| *USA* | 0.6 ± 3.3<br>0.84 | 1.09 ± 0.33<br>3.5E-3 ** | 0.6 ± 1.8<br>0.75 | 0.44<br>2.3% | 26 | 15 |
| East Asia | 24.5 ± 8.1<br>8.5E-3 ** | 0.16 ± 0.14<br>0.28 | -17.8 ± 5.1<br>3.3E-3 ** | 0.45<br>-4.5% | 19 | 11 |
| *China* | 7.7 ± 9.2<br>0.42 | 0.51 ± 0.19<br>1.9E-2 * | -28.7 ± 5.3<br>1.1E-4 *** | 0.46<br>-5.2% | 17 | 10 |
| Western Europe | 0.6 ± 1.6<br>0.71 | 0.92 ± 0.15<br>3.1E-6 *** | 1.7 ± 1.8<br>0.35 | 0.61<br>-6.7% | 26 | 6 |

**Table 14.**
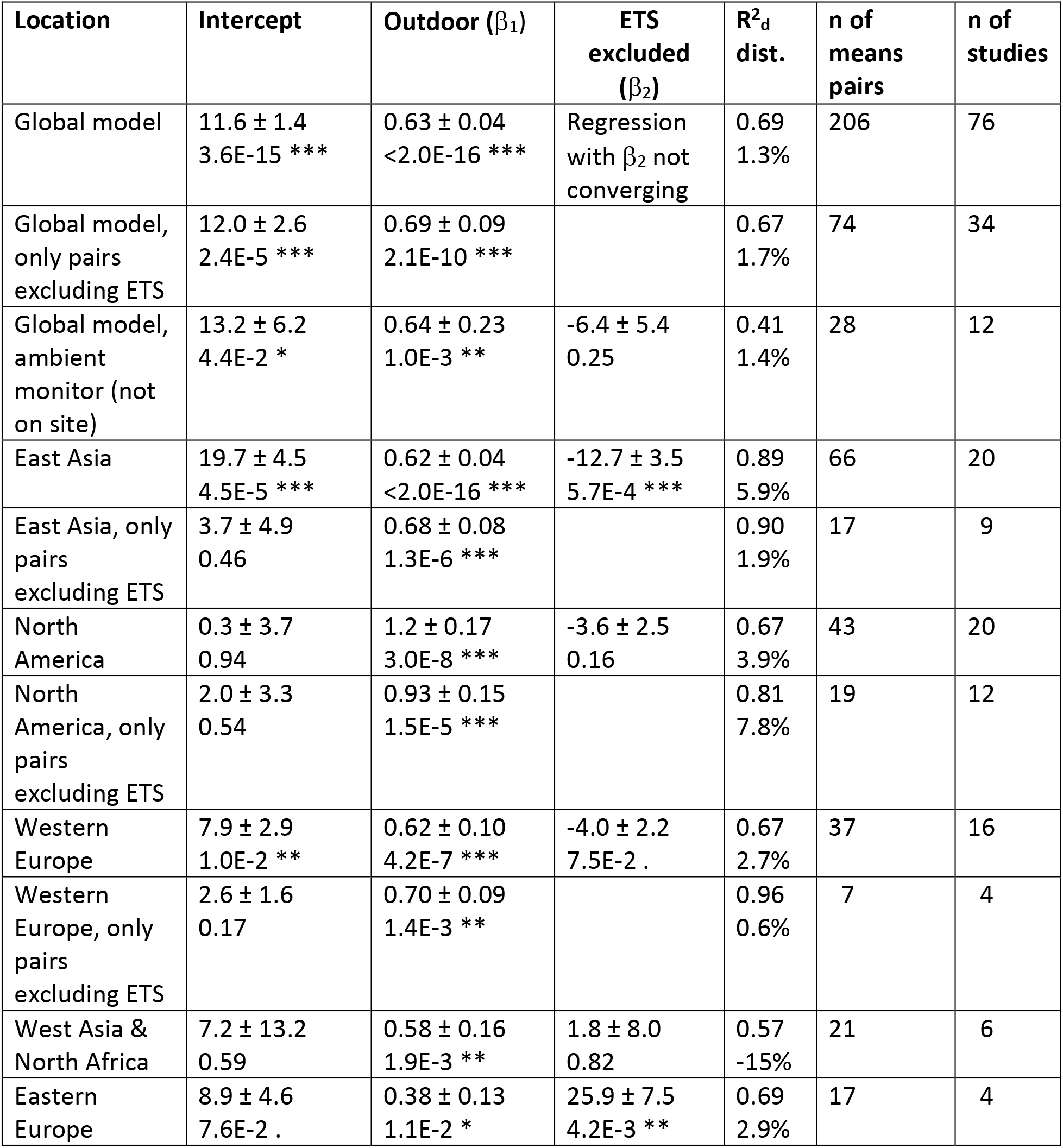
Coefficients mean, standard error, and significance for log-normal weighted regression of mean concentrations of indoor PM10 (and PM1-10) with respect to concentrations measured outdoor (on site) or, when specified, at an ambient monitor (not on site): Indoor ∼ Intercept + β1 * Weight *Outdoor + β2 * ETS.excluded (True or False). Concentrations in μg/m^3^. Weight is total sampling time. Only results from locations with at least 4 different studies are presented. Significance codes: *** <1E-3 **<1E-2 *<5E-2 .<1E-1

**Table 15.** Coefficients mean, standard error, and significance for log-normal weighted regression of mean concentrations of indoor PM2.5-10 (and PM2-10) with respect to concentrations measured outdoor (on site): Indoor ∼ Intercept + β1 * Weight *Outdoor + β2 * ETS excluded (True or False). Concentrations in μg/m^3^. Weight is total sampling time. Only results from locations with at least 4 different studies are presented. Significance codes: *** <1E-3; **<1E-2; *<5E-2; .<1E-1

| Location | Intercept | Outdoor ($\beta_1$) | ETS excluded ($\beta_2$) | $R^2_d$ dist. | n of means pairs | n of studies |
| --- | --- | --- | --- | --- | --- | --- |
| Global model | $2.0 \pm 1.2$<br>9.6E-1 . | $0.74 \pm 0.15$<br>1.2E-5 *** | $1.8 \pm 1.2$<br>3.8E-3 ** | 0.47<br>1.8% | 51 | 20 |
| Global model, only pairs excluding ETS | $3.0 \pm 1.8$<br>0.10 | $0.83 \pm 0.21$<br>4.2E-4 *** | | 0.40<br>2.4% | 31 | 14 |
| North America | $4.5 \pm 1.8$<br>2.3E-2 * | $0.33 \pm 0.22$<br>0.17 | $0.2 \pm 1.5$<br>0.87 | 0.09<br>1.1% | 23 | 10 |
| East Asia | $2.1 \pm 1.7$<br>0.26 | $0.74 \pm 0.22$<br>1.3E-6 *** | $-1.0 \pm 1.5$<br>0.53 | 1.02<br>3.4% | 12 | 4 |

**Table 16.** Coefficients mean, standard error, and significance for log-normal weighted regression of mean concentrations of indoor PM1 (and PM0.1-1) with respect to concentrations measured outdoor (on site): Indoor ∼ Intercept + β1 * Weight *Outdoor + β2 * ETS excluded (True or False). Concentrations in μg/m^3^. Weight is total sampling time. Only results from locations with at least 4 different studies are presented. Significance codes: *** <1E-3; **<1E-2; *<5E-2; .<1E-1

| Location | Intercept | Outdoor ($\beta_1$) | ETS excluded ($\beta_2$) | $R^2_d$ dist. | n of means pairs | n of studies |
| --- | --- | --- | --- | --- | --- | --- |
| Global model | $2.3 \pm 0.5$<br>1.1E-5 *** | $0.55 \pm 0.04$<br><2.0E-16 *** | $-2.3 \pm 0.8$<br>3.8E-3 ** | 0.76<br>-13% | 52 | 16 |
| Global model, only pairs excluding ETS | $2.8 \pm 1.6$<br>0.11 | $0.38 \pm 0.08$<br>5.5E-4 *** | | 0.53<br>-22% | 13 | 4 |

**Table 17.** Coefficients mean, standard error, and significance for log-normal weighted regression of mean concentrations of indoor UFP (and PM0.1) with respect to concentrations measured outdoor (on site): Indoor ∼ Intercept + β1 * Weight *Outdoor + β2 * ETS excluded (True or False). Concentrations in cm^-3^. Weight is total sampling time. Only results from locations with at least 4 different studies are presented. Significance codes: *** <1E-3; **<1E-2; *<5E-2; .<1E-1

| Location | Intercept | Outdoor ($\beta_1$) | ETS excluded ($\beta_2$) | $R^2_d$ dist. | n of means pairs | n of studies |
| --- | --- | --- | --- | --- | --- | --- |
| Global model | $1\ 654 \pm 868$<br>7.5E-2 . | $0.79 \pm 0.14$<br>4.8E-5 *** | $-2\ 207 \pm 1\ 479$<br>0.16 | 0.66<br>-5.7% | 20 | 9 |
| Global model, only pairs excluding ETS | $-1\ 366 \pm 1\ 973$<br>0.50 | $0.86 \pm 0.18$<br>6.8E-4 *** | | 0.64<br>-3.3% | 13 | 5 |

Most regression models (and all those with larger data sets) show a significant effect of outdoor PM on indoor concentrations (β_1_ regression coefficient) at all size fractions. The magnitude and uncertainty of these coefficients, approximating the infiltration factor, vary for different size fractions and locations. The global model for PM_2.5,_ which has the most data, has outdoor air contributing 86% ± 5% of its PM_2.5_ concentration to indoor air in homes, while the United States model has 79% ± 13%, and East Asia 48% ± 4%, for example. This factor varies, but not in a consistent pattern, when accounting for uncertainty, for the other size fractions, with 63% ± 4% for PM_10_, 74% ± 15% for PM_2.5 −10_, 55% ± 4% for PM_1_, and 79% ± 14% for UFP in the global models. The intercept terms, approximating indoor source contributions, is less often significant due to greater uncertainty, with similar values of PM_2.5_ (2 to 11 μg/m^3^) in the global models, North America and Western Europe, and higher values in East Asia. There was more similarity in intercept terms for PM_10_ (except for lower values in North America), and PM_2.5-10_. Due to the lower number of measurements, only global regressions were performed for PM_1_ and UFP.

Some coefficients β_1_ for outdoor air are greater than 1.0, which, if interpreted as infiltration factors, do not have a physical meaning in a mechanistic model. In this statistical model, such values indicate that outdoor air pollution has a positive correlation with some indoor sources. To remain within the variables of the model, this is often the case for ETS: spatially, as both ambient air pollution and smoking are higher in some locations and both low in others; and temporally where ambient air pollution declined over the survey period similarly to smoking prevalence^47, 48, 50^. But other correlations may exist, reflecting excess burden of ambient air pollution on poorer communities that can also least afford the energy and material costs of filtration, ventilation, and other measures to reduce exposure to indoor sources^51–54^. This effect (multicollinearity) also makes the regressions underestimate the intercept, interpreted as indoor-generated PM. The issue is especially notable for several models for North America (Tables 12-14), where both the simultaneous decline of outdoor PM and smoking prevalence^47, 48, 50^, and disparities in exposure by socioeconomic status^51^ are well documented, but it is likely affecting other estimates. Estimates using only measurements in ETS-free homes partially correct for this effect and generally show lower coefficients, for the smaller size fractions, but not for PM_10_ and PM_2.5-10_. This effect also explains why our estimates of infiltration factors for PM_2.5_ are in some cases higher than those from measurements and modeling in the literature. For example, Fazli & Stephens^55^ calculated values of 0.4-0.5, and compiled values of 0.45-0.75 from the literature; MESA Air^56^ had values for different communities from 0.47 ± 0.15 to 0.82 ± 0.14; and EXPOLIS^8^ a range of 0.59 ± 0.17 to 0.70 ± 0.12 for different European cities. Estimates of infiltration factors for UFP (0.79-0.86) in the global models are also higher than may be expected from physical considerations (i.e. smaller than for PM_2.5_) and reported values of 0.1 to 0.4^55, 57^. Regression results from East Asia (0.31-0.48), where declines in ambient air pollution and smoking prevalence^50, 58^ have been smaller^59, 60^ and less correlated, are more in line with literature estimates.

The coefficients of the regression models, despite the limitations above, can be used to estimate the fraction of indoor PM that is generated from indoor sources vs. the fraction infiltrated from outdoors. A detailed exploration of these relative contributions is beyond the scope of this summary, but an example can help to better understand the implications of the results in Tables 12 to 17. The breakdown of sources for PM_2.5_ in the United States is particularly interesting as it can be compared to other estimates. From Table 12, the regression coefficients can be used to approximate F_INF_ = 0.788 ± 0.128 and indoor source contribution: 6.99 ± 1.62. At the mean level of outdoor concentration for the data set, 13.14 μg/m^3^, the contribution of outdoor PM_2.5_ is 10.35 ± 1.68 μg/m^3^, so the proportion of indoor generated PM_2.5_ is 6.99/(6.99 +10.35) = 40.3% ± 10.8%. Azimi & Stephens^61^ used data from both RIOPA and MESA Air studies to calculate the fractions of *total* PM_2.5_ exposures taking place in different microenvironments. Exposure in residences was about 70% of total exposure, 42% ± 24 % (of total) from outdoor-generated particles and 28% ± 26 % from those generated indoors, so that the fraction of indoor-generated to total residential particle exposure is 40% ± 42% (using error propagation) for this microenvironment. It is important to note the wide uncertainties around both estimates, despite the coincidental exact match of results and, given the above limitations from multicollinearity, we must consider ours as a lower-bound estimate of the indoor-generated fraction.

Regressions models using outdoor concentrations from off-site ambient monitors (Table 13) show lower coefficients for F_inf_ of PM_2.5_ in the global models, compared to those using outdoor concentrations measured just outside homes (Table 12), and account for a lower fraction of explained variance, as may be expected from the spatial misalignment introduced by these measurements. Results for regional and country models are more uncertain and do not show consistent patterns compared to corresponding models using on-site outdoor concentrations.

In the weighted regression models, infiltration of outdoor air and ETS alone accounted for a fraction ranging from 25% of the observed PM_2.5_ variability for Finland, to 96% for Taiwan (limiting the range to R^2^_d_ with distortions of <5% in Table 12). Infiltration alone explained almost 55% of the variability (with minimal overestimation) in the global model of samples taken in the absence of ETS. As outdoor PM concentrations decrease, infiltration of PM from outdoors accounts for a lesser and lesser share of indoor PM. This pattern can also be seen in Figure 9, where above about 50 μg/m^3^ of outdoor PM_2.5_, indoor/outdoor ratios above 1 are rare, but become frequent at lower outdoor concentrations. For PM_10_ (Table 14), the fraction of variability accounted for by infiltration and ETS is comparable to or higher than for the corresponding PM_2.5_ models, and even higher for ETS-free homes, suggesting a similarly important role for outdoor sources for this size fraction. However, this pattern does not hold for PM_2.5-10,_ with infiltration and smoking explaining 47% of the variability, and infiltration alone 40%. The regional PM_2.5-10_ models for East Asia and North America differ starkly, with the former accounting for almost all the variance and the latter for only 9%. Models of smaller size fractions, PM_1_ mass and UFP number, account for 53% to 76% of the variability, although these measures are substantially negatively distorted and true values are likely higher.

**Figure 7.**
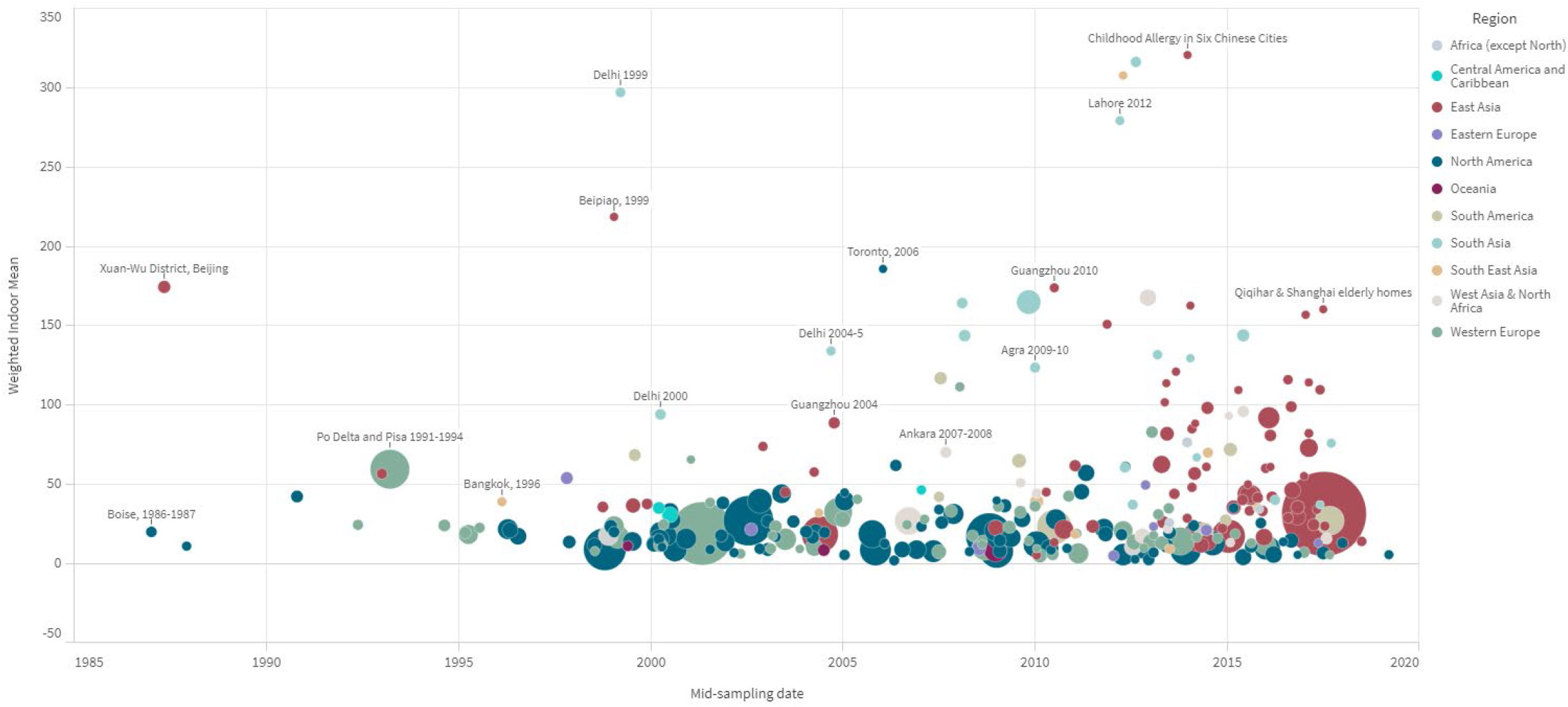
Weighted study means of PM2.5 (μg/m^3^) by mid-point of sampling campaign and region. Marker size is proportional to total study sampling time.

**Figure 8.**
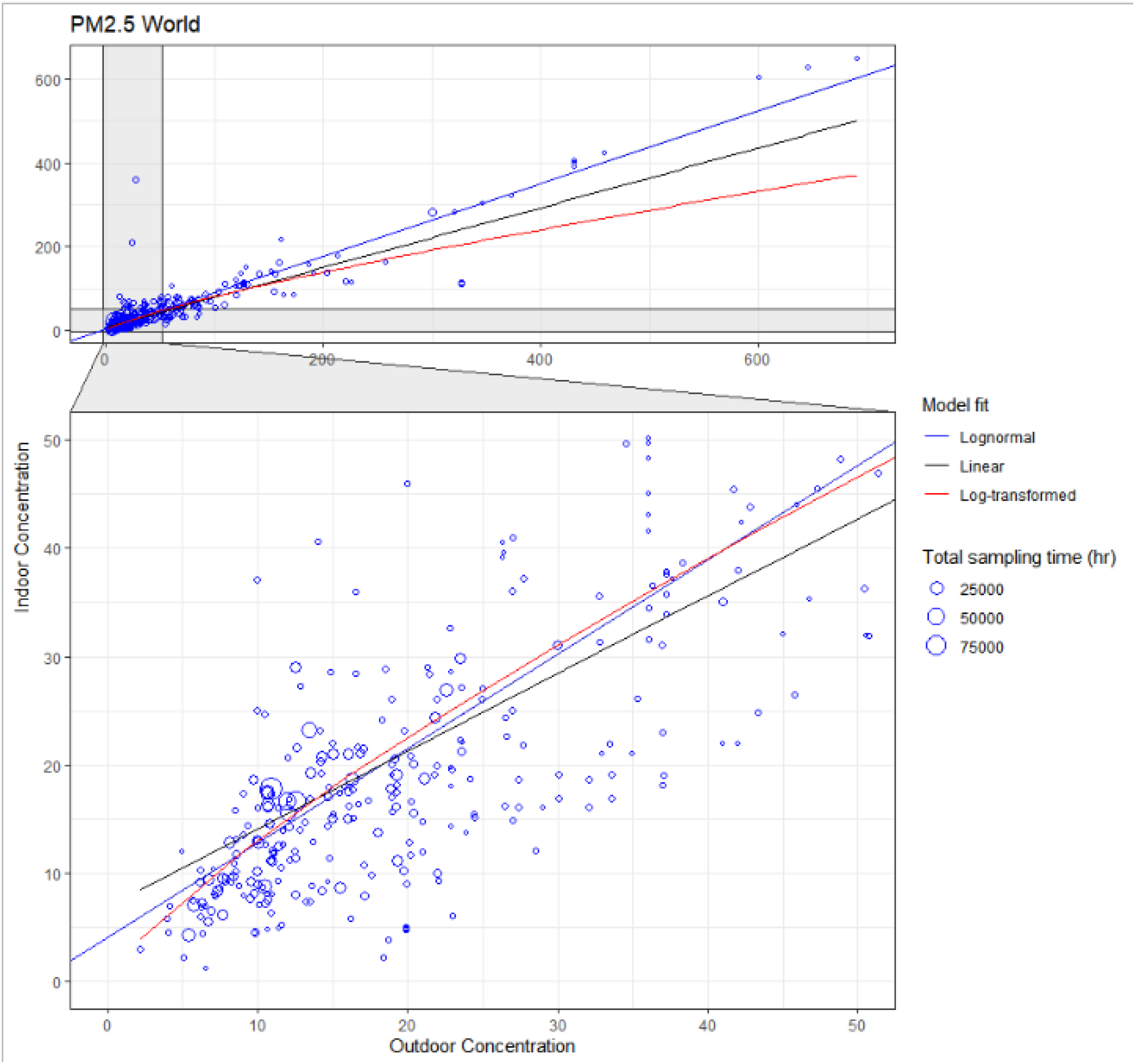
Scatterplot of mean indoor and outdoor (on site) concentrations pairs for PM2.5 (in μg/m^3^). Circle size indicates the total sampling time of the measurements (in hours) across all sampled buildings for a pair of mean concentrations in a study, which were used as weights in the regressions. The Model Fit lines show 3 different fitting approaches: lognormal regression, simple linear regression on unmodified data, and linear regression on log-transformed data. Bottom graph shows magnified region of top graph.

**Figure 9.**
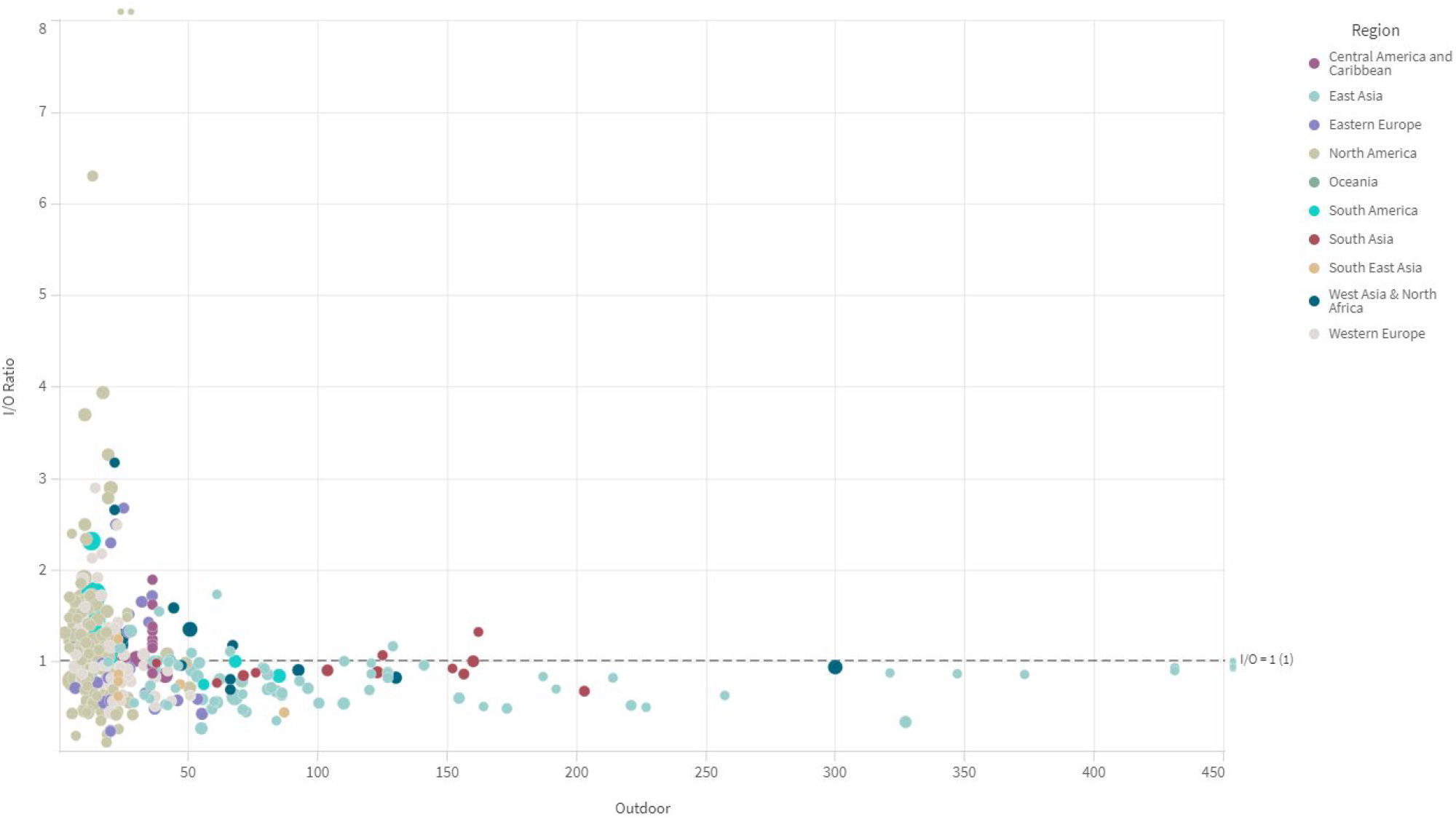
Indoor/outdoor ratios of PM2.5 with respect to outdoor (on site) concentrations (μg/m^3^). Four points outside the grid are shown that extend far beyond the axes ranges shown.

Although outdoor concentrations explain a good fraction of the variability of indoor concentrations between studies, this may not necessarily be the case within individual studies. Within-study variability explained by ambient air (R^2^) spans a wide range in larger multi-site studies, such as 0.06 to 0.44 (0.18 overall) for the RIOPA study^2^, 0.40-0.83 (0.58 overall, calculated) for EXPOLIS^8^, and 0.13 to 0.72 for RUPIOH^57^ (calculated). In general, the variability of indoor concentrations (over time or between homes) is larger than that observed outdoors, on site or at ambient air monitors. Within-study variabilities of mass concentrations, expressed as relative standard deviations, were approximately 50-60% greater indoor than outdoors (Figure 10) for all particle sizes combined and for most size fractions, except for PM_1_, which had similar variability indoor and outdoor; and ambient PM_2.5 -10_ (with greater uncertainty, based only on 6 studies). Indoor variability was even greater for number concentrations, more than twice that reported for outdoors samples. This larger within-study variability indoors reflects the greater complexity of indoor environments, which even when experiencing the same outdoor concentrations differ in air exchange and deposition rates, as well as the variety and intensity of indoor sources.

**Figure 10.**
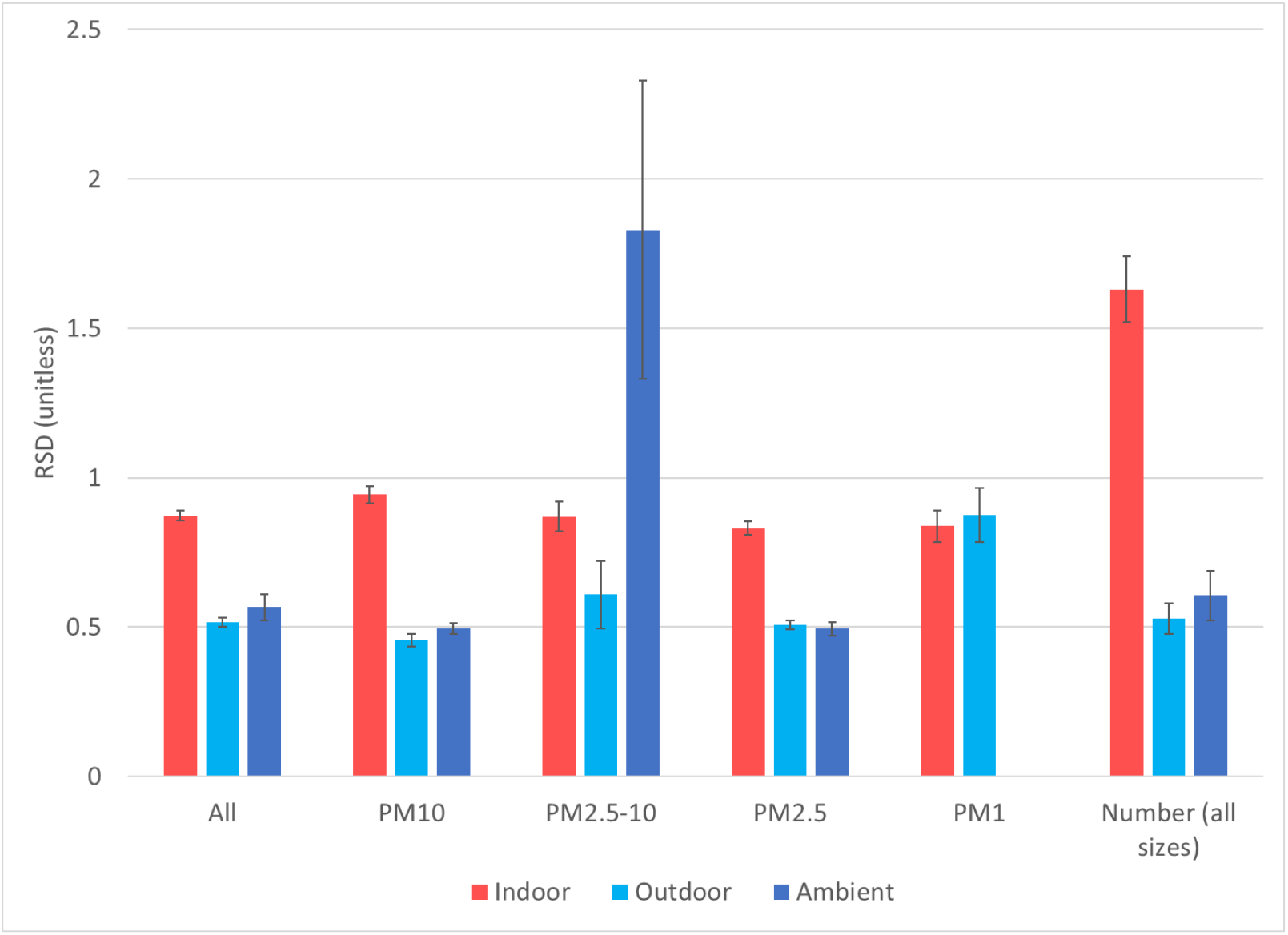
Variability of indoor, outdoor, and ambient monitor measurements expressed as relative standard deviation (SD/Mean). Error bars are standard errors. Only means and SD reported in the studies are included, without calculated ones.

#### 3.4.2. Environmental Tobacco Smoking (ETS)

About 42% of the studies reviewed provided at least some results from measurements in indoor environments free of ETS. These may have been the results of sampling designs that specifically excluded homes with ETS from participation, or in other cases a subset of the measurements limited to ETS-free homes. The remaining studies either did not mention ETS or did not provide separate data. As a result, it is not possible to estimate the full effect of ETS on indoor PM concentrations, but only to distinguish between ETS-free homes and homes with a mix of both smoking and non-smoking conditions, whose proportions vary from case to case. Despite this imperfect discriminator, estimates of PM from ETS-free residences can help us understand indoor air quality in the absence of an important PM source that is within an individual’s ability to control. Tables 2-6 show that the concentrations of PM of almost all sizes under ETS-free conditions was generally very similar to or lower than concentrations in the larger sets of studies. In a few cases, most noticeably for Eastern Europe and South Asia, concentration in ETS-free conditions were higher, but this can be explained by the small number of studies without ETS, mismatched with the larger collection in terms of locations and timing.

Similarly, pooled regression models assessing time trends (Tables 8-9) show significant differences of −3 μg/m^3^ (North America and Western Europe) to −13 μg/m^3^ (East Asia) for PM_2.5_, but no significant differences for PM_10_. Indoor to outdoor regression models (Tables 12-17) also show consistent reductions around −3 μg/m^3^ for North America and Western Europe and −8 μg/m^3^ for East Asia (although no reduction for China alone). PM_10_ indoor-outdoor regressions again do not show significant reductions from ETS-free conditions, except for East Asia (−13 μg/m^3^). Coarse PM and PM_1_ indoor-outdoor regressions have significant increases and decreases respectively for the global models, while the reduction for UFP is not significant. Finally, we examined just the subset of studies that reported results from both ETS-free and mixed ETS conditions (Figure 11 and Table S3). Concentrations are significantly different for PM_2.5_ but not for PM_10_.

**Figure 11.**
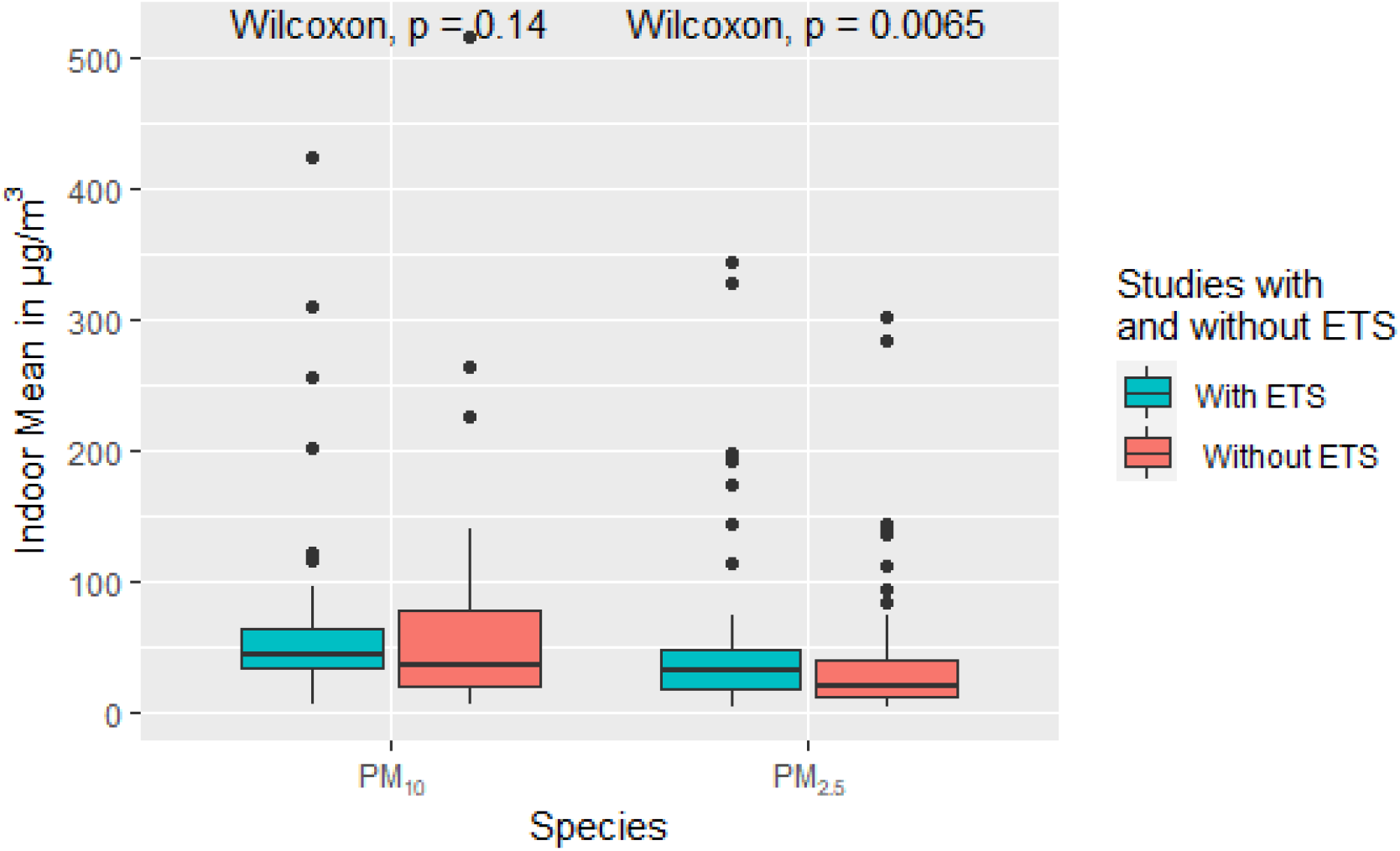
Box plot of indoor mean values for PM10 and PM2.5 for studies with and without ETS. Statistics labeled ‘Without ETS’ are restricted to measurements in homes without environmental tobacco smoke, while statistics labeled ‘With ETS’ contains measurements in homes with and without environmental tobacco smoke. Additional descriptive statistics for these PM species as well as other size fractions can be found in the in the supplementary Information (Table S3)

Overall, these results are consistent with the well-known impact of ETS on finer particles indoors, within the limitations of the crude separation of exposure conditions available.

#### 3.4.3. Air cleaners

Only 31 studies (7%) reported on the use of air cleaners in homes during measurements, typically in intervention studies that provided the devices to study participants and prescribed their hours of operation. With one exception, all air cleaners worked through mechanical filtration. Because of the small number of results, only a few pooled regression models could estimate their effect (Tables 8-9). For PM_2.5_, reductions ranged from −4.5 ± 1.2 μg/m^3^ for North America, to −6.2 ± 2.2 μg/m^3^ for East Asia, and −6.8 ± 2.0 μg/m^3^ for Western Europe. All effects were significant. These values represent relative reductions from (weighted) mean indoor concentrations of −26.8% ± 7.1%, −17.7% ± 6.3%, and −29.4% ± 8.7% respectively. For PM_10_, the pooled model for East Asia estimated a significant reduction of −56.5 ± 24.5 μg/m^3^, or −91.6% ± 39.7% of the weighted mean concentration. Limiting the comparison only to studies that reported measurement with and without air cleaners in use (Table S4), the difference of −11.6 μg/m^3^ is significant only for PM_2.5_ (Figure 12). Overall, these results provide evidence of meaningful reductions in indoor PM_2.5_ concentrations (and less clearly for PM_10_) during home use of air cleaners.

**Figure 12.**
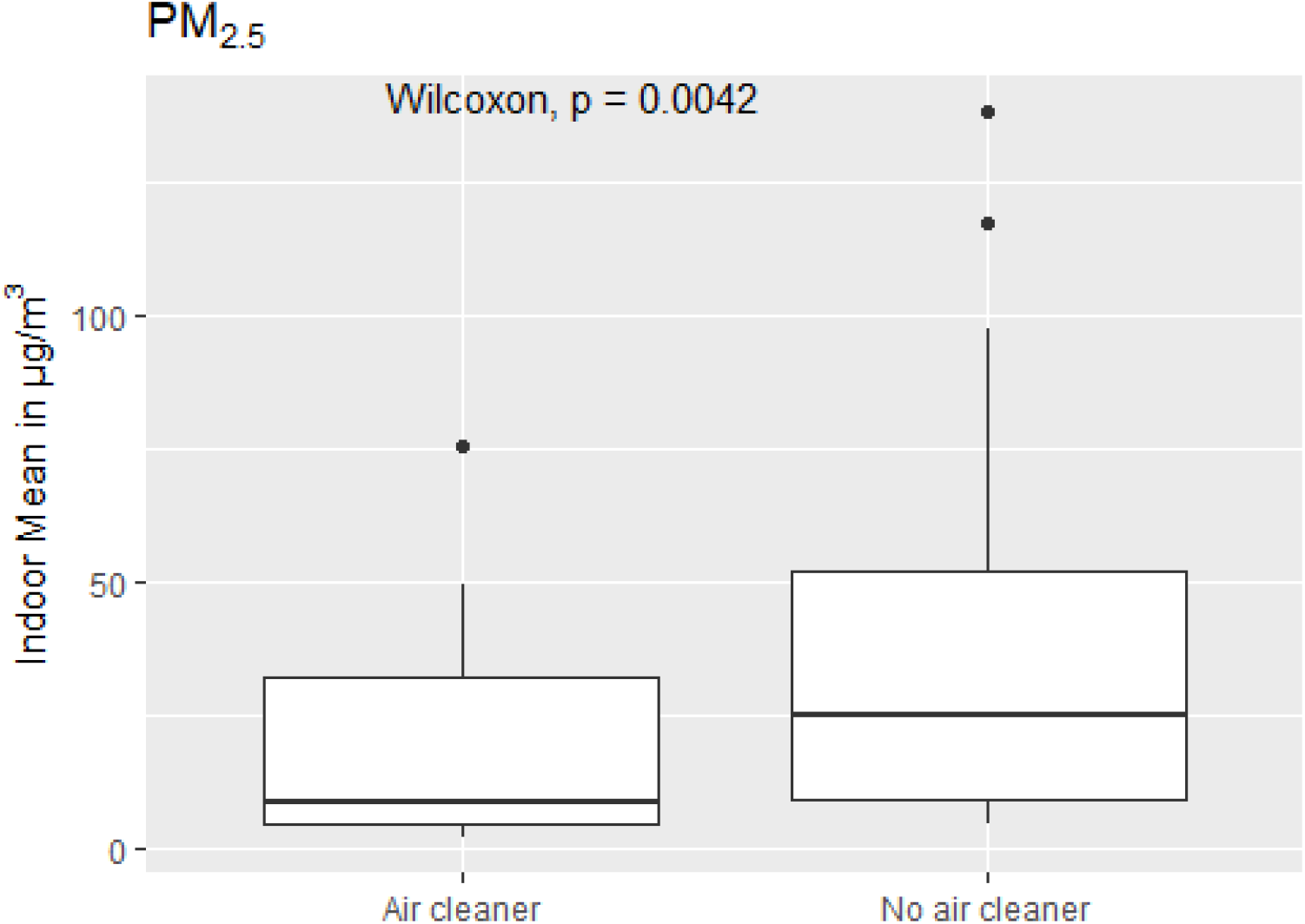
Box plot of indoor mean values for PM2.5 for studies reporting results both with and without air cleaners. Statistics labeled ‘No air cleaner’ are restricted to measurements in homes without air cleaners, yet the corresponding study contains measurements with air cleaners. Statistics labeled ‘Air cleaner’ contains measurements in homes with air cleaners. Additional descriptive statistics for this PM species as well as other size fractions can be found in the in the supplementary Information (Table S4)

## 4. Conclusions

This survey compiled more than 2,000 sets of indoor PM measurements from hundreds of studies worldwide, representing sampling in more than 21,200 homes, for almost 5 million hours across those homes. Three world regions - North America, Western Europe, and East Asia - have been sampled more extensively and the common characteristics of their indoor environments, stemming from their mostly temperate climates, vastly urban and highly economically developed societies, inevitably shape our knowledge of indoor PM concentrations. Even within these more heavily studied countries, many geographic areas remain poorly characterized, especially those in less urban settings, which may potentially differ in terms of indoor sources and building characteristics.

Taken together, however, these measurements of indoor PM concentrations contribute to an improvement in our knowledge of residential PM exposures compared to estimates based primarily on ambient air pollution measurements or models, and behavioral survey data. Our results provide ranges of mean PM concentrations measured in indoor environments (e.g., 7.7- 29.5 μg/m^3^ PM_2.5_ for 10^th^ and 90^th^ percentiles in North America; 20-112 μg/m^3^ PM_10_ for East Asia), as well as their variability. The majority of North American and European indoor environments studied were generally similar with respect to PM concentrations, clustering around the same relatively narrow ranges. Greater variability was observed for all regions of Asia, demonstrating a need for additional measurements, with careful consideration of sampling size in future studies conducted in these regions. In addition to supplying information for exposure and risk assessments, these systematic data could also be of value to product development for a growing list of consumer products related to indoor air quality.

The limited availability of health-based standards specific to indoor PM concentrations makes it difficult to evaluate the public health implications of the concentrations collected in this survey. While ambient standards and guidelines are not always appropriate benchmarks for indoor exposure risk, exposures to levels of concerns for public health are commonly occurring in many indoor environments. Concerns for even greater potential risks are raised when considering that the distributions of means for different studies we report represent a range of exposure concentrations extending, in part, beyond the values tracked. The maxima of the published values provide an indication of just how high indoor PM concentrations can become in real exposure scenarios, exceeding milligrams per cubic meter for PM_2.5_ and PM_10_ even in the absence of solid fuel combustion sources.

The indoor concentrations of PM of different size fractions were mostly stable over the survey period, with the notable exceptions of PM_2.5_ in Europe and most consistently in North America, which declined, at rates of 0.9 and 0.3 μg/m^3^/year respectively. North American concentrations of PM_10_ also had significant declines, of about 1 μg/m^3^/year. The reasons for these declines may perhaps, in part, be attributed to declining tobacco use indoors, and decreasing ambient air concentrations of regulated PM size fractions.

Outdoor air concentrations of PM were consistently a major driver of indoor concentrations in all regions and for all size fractions. The infiltration factors calculated from regressions of paired concentrations were often higher than those published from mechanistic and modeling studies. This may possibly be explained by the correlation between ETS and ambient air pollution over time and by inequalities in the burden of air pollution exposures based on socioeconomic status. Homes free of ETS generally had significantly lower concentrations of PM_2.5,_ as expected. Another factor with large beneficial impacts on indoor PM_2.5_ was the use of filtration-based air cleaners.

Although our study offers the most expansive review of the available literature and data on indoor PM measurements and studies to date, some results are conspicuous for their absence. First, knowledge on indoor PM exposures for some of the world’s largest population centers rests on a handful of studies, population-representative studies are rare everywhere, while little or no data exist for some regions undergoing major economic and social transformations. Second, our understanding of the smaller PM size fractions indoors remains limited, as is the evidence about effective interventions within individual control. Third and perhaps most importantly, even where data do exist, any public health response must confront the lack of systematic and comprehensive records on indoor PM and its sources, as well as a challenging interpretation of measured levels in the near-absence of health-based standards intended for indoor exposures.

## Data Availability

All the data that support the findings of this study are available in peer-reviewed publications. Data sources, by country or territory, that were used to compile the data are listed in the Supplementary Information

## Authors contributions

Based on <u>Contribution Roles Taxonomy</u>

**Vito Ilacqua**: Conceptualization (lead); investigation (equal); data curation (lead); formal analysis (lead); writing – original draft (lead); writing – review and editing (equal); methodology (lead); visualization (equal); project administration(equal). **Nicole Scharko**: investigation (equal); formal analysis (supporting); writing – original draft (supporting); writing – review and editing (equal); validation (equal); visualization (equal). **Jordan Zambrana**: conceptualization (supporting); investigation (supporting); validation (equal); writing – review and editing (equal). **Daniel Malashock:** project administration (equal); validation (equal); writing: review and editing (equal); visualization (supporting)

## Data availability statement

The data that support the findings of this study are available in peer-reviewed publications. Data sources, by country or territory, that were used to compile the data are listed in the Supplementary Information.

## Funding statement

This research received no external funding.

## Conflict of interest disclosure

The authors declare no conflict of interests.

## Online Supplementary Information

Additional tables, figures, and the list of all sources for the data compiled in this survey are in the Supplementary Information.

## Acknowledgements

The authors would like to thank Prof. Vito Muggeo of the University of Palermo for promptly updating the l*ogNormReg* R package when contacted, to enable weighted regressions, and for the fruitful discussions.

## Disclaimer

The views expressed in this document are solely those of the authors and do not necessarily reflect those of the United States Environmental Protection Agency. The US EPA does not endorse any products or commercial services mentioned in this publication.

## Notes

### Competing Interest Statement

The authors have declared no competing interest.

### Funding Statement

This study received no external funding.

