## Supplementary Information for "Survey of residential indoor Particulate Matter measurements 1990-2019"

### Supplemental Material for Survey of residential indoor Particulate Matter measurements 1990-2019

*Table S1. Summary of existing literature reviews of indoor PM.*

Table S1a. Studies that reviewed residential environments and do not provide pooled statistics.

| Study | Timeframe* | Geography | Indoor micro-environment | Particle metrics of interest | Individual study statistics provided | # of Studies Reviewed | Additional notes |
|---|---|---|---|---|---|---|---|
| Vardoulakis et al., 2020 | 2000-2017 | No limitations | Domestic indoor environments | $PM_{2.5}$ and $PM_{10}$ (also include studies with $PM_{2.5-10}$, PNC, UFP etc.) | In the main manuscript they do provide the range for mean concentrations for $PM_{2.5}$ and $PM_{10}$. | 73 studies for $PM_{2.5}$ and 37 studies for $PM_{10}$. | Excluded studies published in other languages and biomass burning homes. |
| Ye et al., 2017 | last 10 years (assuming 2006-2016) | China (separated by rural and urban) | Chinese residential buildings | $PM_1$, $PM_{2.5}$ and $PM_{10}$ | Min, max and mean. | 11 studies for urban and 7 studies for rural (for all PM species) | Only English and Chinese literature reviewed. Studies needed normal ventilation conditions. |
| Li et al., 2017 | 2000-2016 | No limitations | All indoor environments (restaurant, residence, school etc.) | $PM_{2.5}$ | Statistics provided in their review table varied depending on the study results. | 11 studies with residential homes as the micro-environment | Only English and Chinese literature reviewed. |

Table S1b. Studies that reviewed residential environments and report pooled statistics.

| Study | Timeframe* | Geography | Indoor micro-environment | Particle metrics of interest | Summary statistics provided and quantitative approach | # of Studies used in analysis | Additional notes |
|---|---|---|---|---|---|---|---|
| Logue et al., 2011 | 1995–2010 | US and other indus-trialized nations | residences | $PM_{2.5}$ | Mean, 25th, 50th, 75, and 95 percentiles. Studies were weighted by the number of measurements within each study | 13 studies for PM2.5 used to determine weighted mean | One of their study reviews was a new home, and the authors do provide statistics for that home for comparison purpose. |
| Morawska et al., 2013 | January 1989 and October 2012 | No limitation | Residences, personal and schools | $PM_{10}$, $PM_{2.5}$ and PN | Min, 1st quartile, median, 3rd quartile and max. Summary statistics were obtained from average values reported in the studies. | 44 studies for all PM species: $PM_{10}$, $PM_{2.5}$ and PN. (residential environments) | Smoking excluded, biomass burning excluded. |
| Morawska et al., 2017 | 1990–2017 | No limitation | Homes, schools and day cares, offices, and aged care facilities | $PM_{10}$, $PM_{2.5}$ and PN | Mean and standard errors (SE), averaging time 24 h. Summary statistics were obtained from weighing the number of individual locations in each study; assumed to be 1 if not reported | 5 studies for $PM_{10}$, 7 studies for $PM_{2.5}$ and 5 studies for PN. (with residential homes as the microenvironment) | Smoking and unoccupied homes, biomass burning homes or homes where normal activity was not allowed were excluded. studies needed a mean, standard deviation, and averaging period of 24 h or multiple of 24. |

***Timeframe or Paper range used in review for homes**

*Table S2. Performance of calculated means and standard deviations where both actual and calculated value exist.*

| **All values (mean = 385 pairs and SD =283 pairs)** | | | | |
|---|---|---|---|---|
| Statistical measure | $RD_{Mean}$ = (Act Mean- Calc Mean)/Act Mean | $AD_{Mean}$ = abs(Act Mean- Calc Mean)/Act Mean | $RD_{SD}$= (Act SD- Calc SD)/Act SD | $AD_{SD}$= abs(Act SD- Calc SD)/Act SD |
| Min | -3.76 | 0.0004 | -12.3 | 0.0001 |
| 1st Qu. | -0.044 | 0.029 | -0.117 | 0.092 |
| Median | 0.020 | 0.064 | 0.074 | 0.189 |
| Mean | -0.001 | 0.126 | -0.077 | 0.370 |
| 3rd Qu. | 0.084 | 0.133 | 0.230 | 0.355 |
| Max | 0.513 | 3.76 | 0.751 | 12.3 |
| **Indoor values only (mean = 249 pairs and SD = 174 pairs)** | | | | |
| Statistical measure | $RD_{Mean}$ = (Act Mean- Calc Mean)/Act Mean | $AD_{Mean}$ = abs(Act Mean- Calc Mean)/Act Mean | $RD_{SD}$= (Act SD- Calc SD)/Act SD | $AD_{SD}$= abs(Act SD- Calc SD)/Act SD |
| Min | -3.76 | 0.001 | -12.3 | 0.0001 |
| 1st Qu. | -0.051 | 0.033 | -0.114 | 0.092 |
| Median | 0.018 | 0.069 | 0.079 | 0.185 |
| Mean | -0.010 | 0.145 | -0.111 | 0.415 |
| 3rd Qu. | 0.098 | 0.151 | 0.227 | 0.415 |
| Max | 0.513 | 3.76 | 0.741 | 12.3 |
| **Outdoor values only (mean = 110 pairs and SD = 84 pairs)** | | | | |
| Statistical measure | $RD_{Mean}$ = (Act Mean- Calc Mean)/Act Mean | $AD_{Mean}$ = abs(Act Mean- Calc Mean)/Act Mean | $RD_{SD}$= (Act SD- Calc SD)/Act SD | $AD_{SD}$= abs(Act SD- Calc SD)/Act SD |
| Min | -1.84 | 0.001 | -7.13 | 0.001 |
| 1st Qu. | -0.035 | 0.026 | -0.164 | 0.112 |
| Median | 0.023 | 0.055 | 0.025 | 0.219 |
| Mean | 0.014 | 0.102 | -0.070 | 0.340 |
| 3rd Qu. | 0.076 | 0.100 | 0.243 | 0.342 |
| Max | 0.419 | 1.84 | 0.751 | 7.13 |
| **Ambient values only (mean = 26 pairs and SD = 25 pairs)** | | | | |
| Statistical measure | $RD_{Mean}$ = (Act Mean- Calc Mean)/Act Mean | $AD_{Mean}$ = abs(Act Mean- Calc Mean)/Act Mean | $RD_{SD}$= (Act SD- Calc SD)/Act SD | $AD_{SD}$= abs(Act SD- Calc SD)/Act SD |
| Min | -0.073 | 0.000 | -0.218 | 0.026 |
| 1st Qu. | -0.007 | 0.019 | 0.074 | 0.082 |
| Median | 0.024 | 0.041 | 0.110 | 0.173 |
| Mean | 0.025 | 0.047 | 0.135 | 0.161 |
| 3rd Qu. | 0.064 | 0.072 | 0.206 | 0.218 |
| Max | 0.156 | 0.156 | 0.447 | 0.447 |

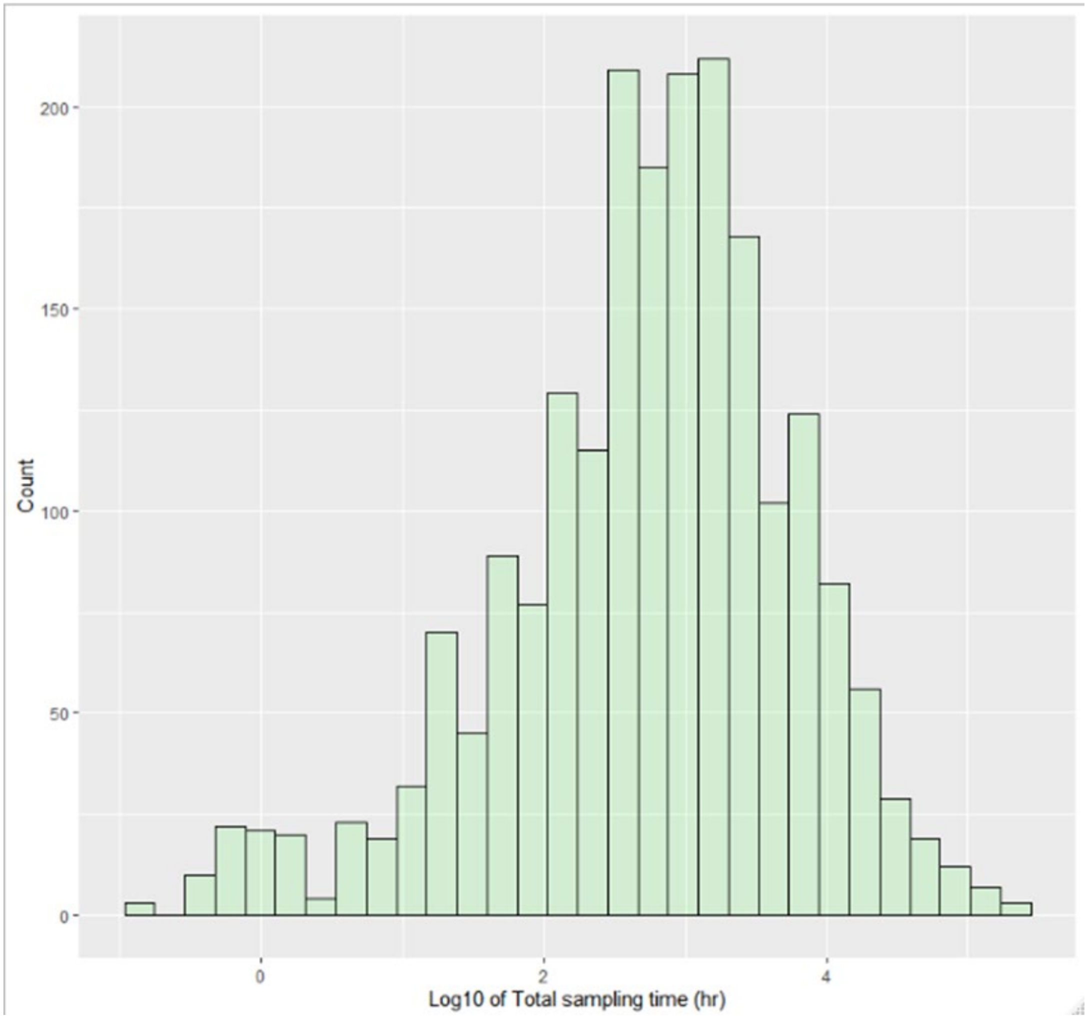

*Figure S1. Distribution of total sampling times (hr) used as weighting factor for the analyses. Values on a log scale.*

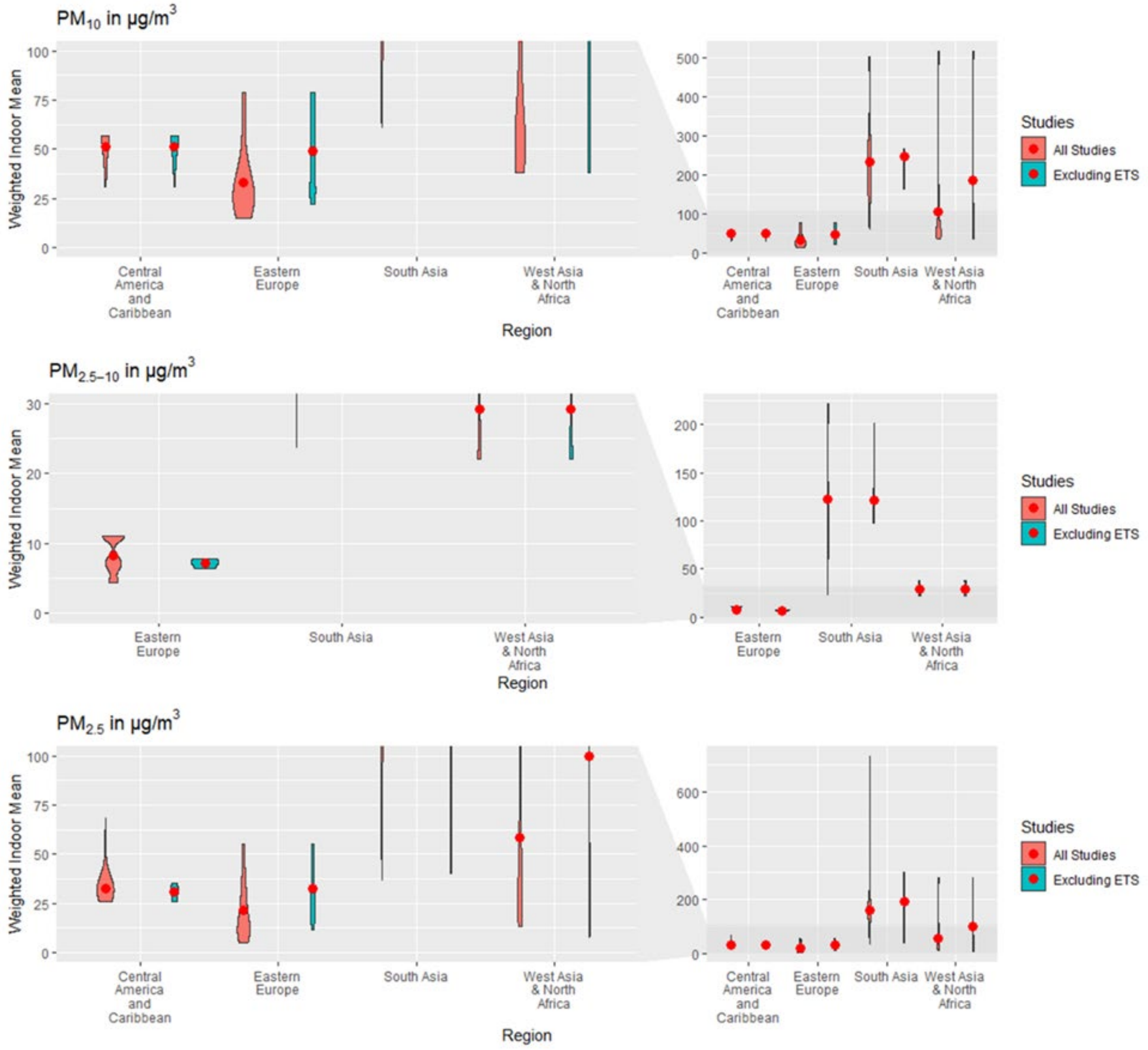

*Figure S2. Violin plots of $PM_{10}$, $PM_{2.5\text{-}10}$ and $PM_{2.5}$ for Central America and Caribbean, Eastern Europe, South Asia and West Asia & North Africa. The red points are displaying the weighted indoor mean, and the density profiles are displaying the weighted distribution of the mean. Studies labeled 'Excluded ETS' are restricted to mean measurements in homes without environmental tobacco smoke.*

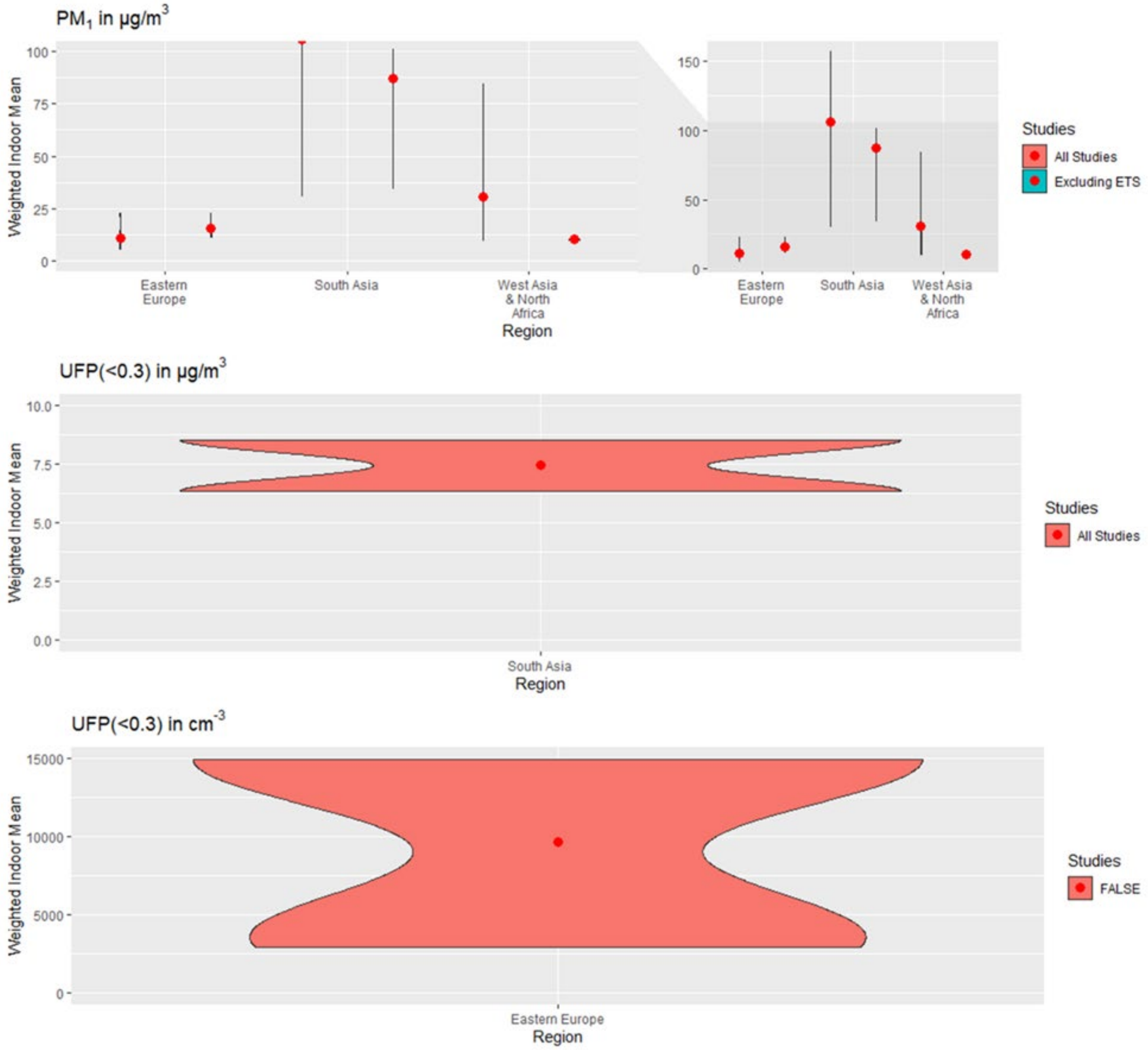

*Figure S3. Violin plots of $PM_1$ and UFP (in $\mu g/m^3$ and $cm^{-3}$) for Eastern Europe, South Asia and West Asia & North Africa. The red points are displaying the weighted indoor mean, and the density profiles are displaying the weighted distribution of the mean. Studies labeled 'Excluded ETS' are restricted to mean measurements in homes without environmental tobacco smoke.*

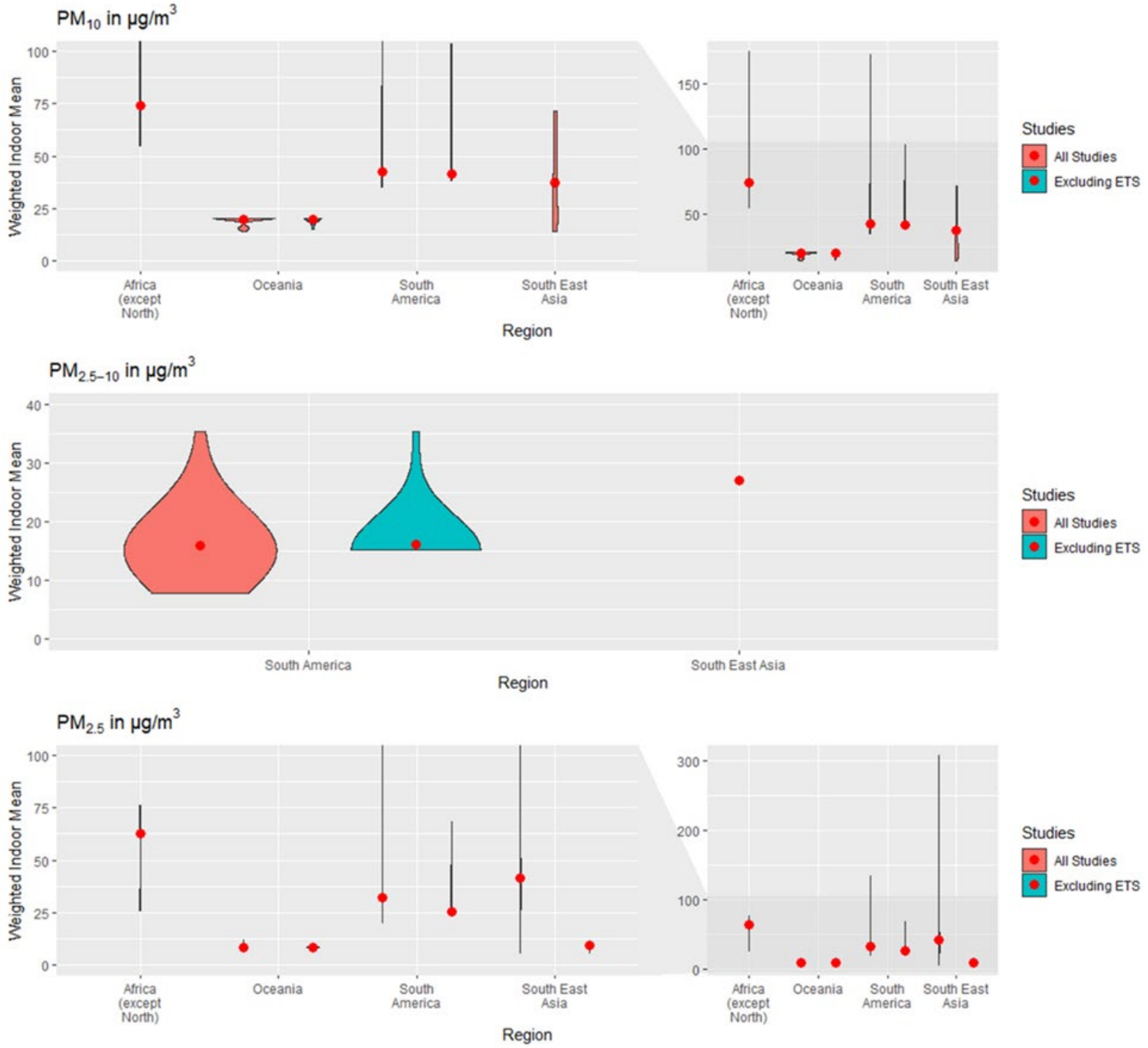

*Figure S4. Violin plots of $PM_{10}$, $PM_{2.5\text{-}10}$ and $PM_{2.5}$ for Africa (except North), Oceania, South America and South East Asia. The red points are displaying the weighted indoor mean and the density profiles, are displaying the weighted distribution of the mean. Studies labeled 'Excluded ETS' are restricted to mean measurements in homes without environmental tobacco smoke.*

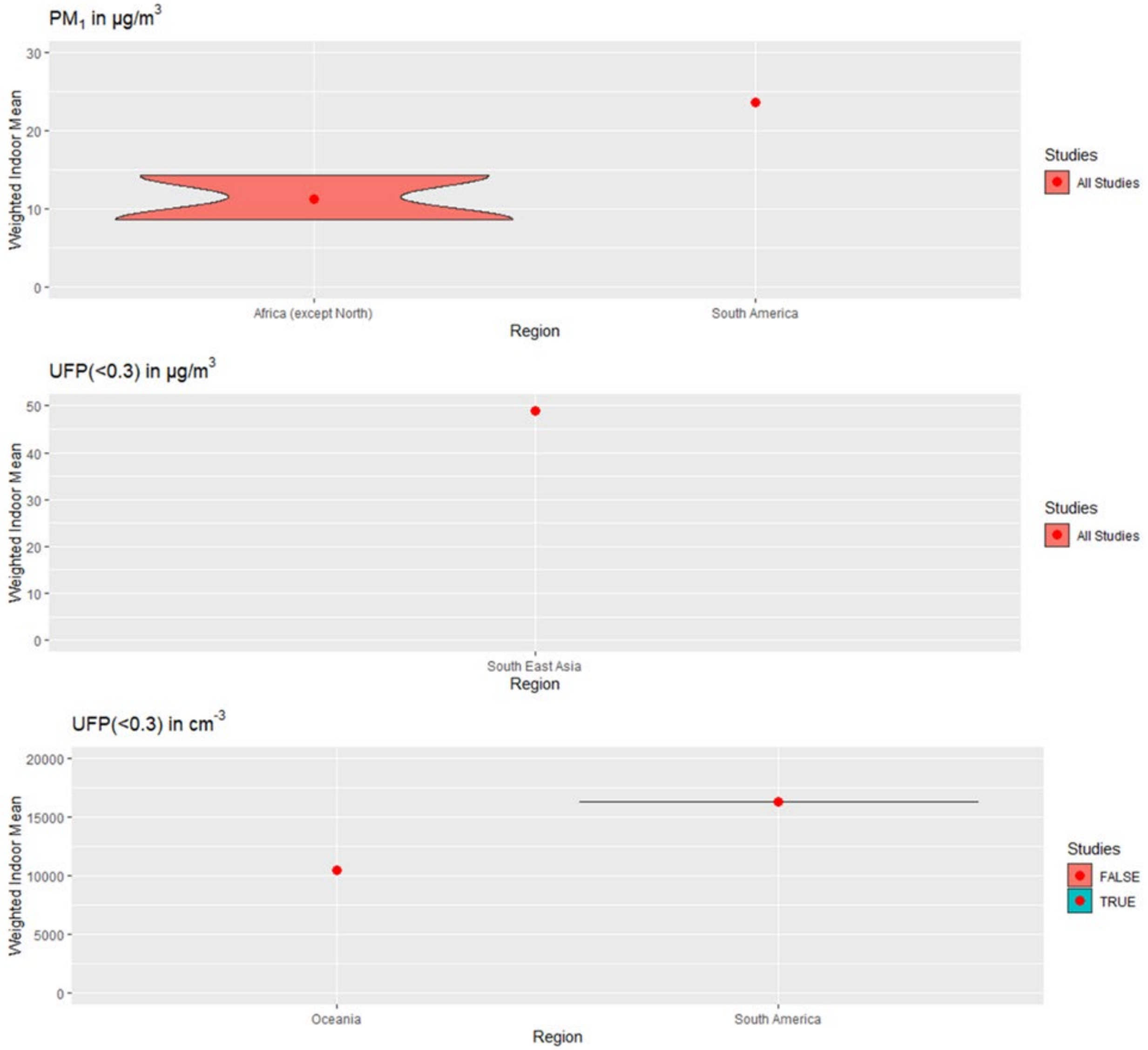

*Figure S5. Violin plots of $PM_1$ and UFP (in µg/m$^3$ and cm$^{-3}$) for Africa (except North), Oceania, South America and South East Asia. The red points are displaying the weighted indoor mean and the density profiles are displaying the weighted distribution of the mean. Studies labeled 'Excluded ETS' are restricted to mean measurements in homes without environmental tobacco smoke.*

*Table S3. Statistics for indoor means from studies that both included and excluded ETS. Statistics labeled 'Without ETS' are restricted to measurements in homes without environmental tobacco smoke, while 'With ETS' contains measurements in homes with and without environmental tobacco smoke. Wilcoxon–Mann–Whitney test was carried out on the original means and the W and p-values are provided in the table. †The exact p-value was not computed; a normal approximation was used.*

| **PM species, unit and with or without ETS** | | **Studies** | **Homes** | **Statistics for Mean Measurements** | | | | | | **Wilcoxon–Mann–Whitney statistics** | |
|---|---|---|---|---|---|---|---|---|---|---|---|
| | | | | n | Mean | SD | Wt. Mean | Min | Max | W | p-value |
| $PM_{2.5}$ ($\mu g/m^3$) | With ETS | 46 | 3302 | 87 | 46.1 | 57.9 | 25.9 | 2.7 | 344 | 4226.5 | 0.0065 |
| | Without ETS | 46 | 1954 | 78 | 36.7 | 52.0 | 21.9 | 2.8 | 302 | | |
| $PM_{2.5\text{-}10}$ ($\mu g/m^3$) | With ETS | 5 | 160 | 20 | 17.3 | 24.8 | 12.3 | 3.4 | 119.2 | 84 | 0.84† |
| | Without ETS | 5 | 53 | 8 | 23.0 | 39.7 | 20.0 | 5.5 | 120.3 | | |
| $PM_{10}$ ($\mu g/m^3$) | With ETS | 28 | 1291 | 49 | 68.3 | 77.8 | 45.1 | 5.5 | 425 | 1158 | 0.14† |
| | Without ETS | 28 | 746 | 40 | 66.3 | 92.6 | 69.9 | 5.6 | 517.74 | | |
| $PM_1$ ($\mu g/m^3$) | With ETS | 6 | 184 | 12 | 38.3 | 42.1 | 27.5 | 5 | 156.3 | 57 | 0.23 |
| | Without ETS | 6 | 142 | 7 | 25.7 | 35.8 | 21.9 | 3 | 101.1 | | |
| Number ($cm^{-3}$) | With ETS | 3 | 203 | 8 | 14223 | 12098 | 10296 | 36.8 | 31816 | 25 | 0.15 |
| | Without ETS | 3 | 84 | 4 | 4964 | 5894 | 1608 | 29.1 | 11815 | | |

*Table S4. Statistics for indoor means from studies with and without air cleaners. Statistics labeled 'No air cleaner' are restricted to measurements in homes without air cleaners, yet the corresponding study contains measurements with air cleaners. Statistics labeled 'Air cleaner' contains measurements in homes with air cleaners. Wilcoxon–Mann–Whitney test was carried out on the original means and the W and p-values are provided in the table. †The exact p-value was not computed; a normal approximation was used.*

| **PM species, unit and with or without air cleaner** | | **Studies** | **Homes** | **Statistics for Mean Measurements** | | | | | | **Wilcoxon–Mann–Whitney statistics** | |
|---|---|---|---|---|---|---|---|---|---|---|---|
| | | | | n | Mean | SD | Wt. Mean | Min | Max | W | p-value |
| $PM_{2.5}$ ($\mu g/m^3$) | No air cleaner | 21 | 591 | 30 | 37.0 | 33.9 | 22.2 | 4.61 | 138.3 | 624 | 0.0042† |
| | Air cleaner | 21 | 520 | 29 | 18.3 | 18.4 | 10.6 | 2.22 | 75.83 | | |
| $PM_{2.5\text{-}10}$ ($\mu g/m^3$) | No air cleaner | 3 | 82 | 4 | 21.2 | 20.6 | 15.9 | 5.7 | 51.2 | 10 | 0.69 |
| | Air cleaner | 3 | 78 | 4 | 14.9 | 13.4 | 12.8 | 3.8 | 34.1 | | |
| $PM_{10}$ ($\mu g/m^3$) | No air cleaner | 3 | 70 | 3 | 45.7 | 24.2 | 27.1 | 18.1 | 63 | 7 | 0.40 |
| | Air cleaner | 3 | 70 | 3 | 26.5 | 18.6 | 7.2 | 5 | 38 | | |
| $PM_1$ ($\mu g/m^3$) | No air cleaner | 2 | 40 | 2 | 47.6 | 0.8 | 47.2 | 47 | 48.2 | 4 | 0.33 |
| | Air cleaner | 2 | 40 | 2 | 28.6 | 3.7 | 27.1 | 25.9 | 31.2 | | |
| Number ($cm^{-3}$) | No air cleaner | 3 | 58 | 3 | 6903 | 11648 | 17787 | 63.4 | 20352 | 6 | 0.70 |
| | Air cleaner | 3 | 71 | 3 | 4090 | 6922 | 10086 | 29.1 | 12083 | | |

Table S5. List of publications with data used in the survey, by country or territory.

| Authors | Title | Journal | Year of publication | Volume | Issue | Pages |
|---|---|---|---|---|---|---|
| **Australia** | | | | | | |
| Cheong CD, HG Neumeister-Kemp, PW Dingle, GS Hardy | Intervention study of airborne fungal spora in homes with portable HEPA filtration units | Journal of Environmental Monitoring | 2004 | 6 | 11 | 866-873 |
| Hargreaves M, S Parappukkaran, L Morawska, J Hitchins, CR He, D Gilbert | A pilot investigation into associations between indoor airborne fungal and non-biological particle concentrations in residential houses in Brisbane, Australia | Science of the Total Environment | 2003 | 312 | NA | 89-101 |
| He C, H Salonen, X Ling, L Crilley, N Jayasundara, HC Cheung, M Hargreaves, F Huygens, LD Knibbs, GA Ayoko, L Morawska | The impact of flood and post-flood cleaning on airborne microbiological and particle contamination in residential houses | Environment International | 2014 | 69 | NA | 43725 |
| Jones J, S Stick, P Dingle, P Franklin | Spatial variability of particulates in homes: Implications for infant exposure | Science of the Total Environment | 2007 | 376 | NA | 317-323 |
| Mazaheri M, S Clifford, R Jayaratne, MA Megat Mokhtar, F Fuoco, G Buonanno, L Morawska | School children's personal exposure to ultrafine particles in the urban environment | Environ Sci Technol | 2014 | 48 | 1 | 113-20 |
| Molloy SB, M Cheng, IE Galbally, MD Keywood, SJ Lawson, JC Powell, R Gillett, E Dunne, PW Selleck | Indoor air quality in typical temperate zone Australian dwellings | Atmospheric Environment | 2012 | 54 | NA | 400-407 |
| Morawska L, CR He, J Hitchins, K Mengersen, D Gilbert | Characteristics of particle number and mass concentrations in residential houses in Brisbane, Australia | Atmospheric Environment | 2003 | 37 | 30 | 4195-4203 |

| **Bangladesh** |
|---|

| | | | | | | |
|---|---|---|---|---|---|---|
| Akther T, Ahmed M, Shohel M, Ferdousi FK, Salam A. | Particulate matters and gaseous pollutants in indoor environment and Association of ultra-fine particulate matters (PM1) with lung function. | NA | 2019 | 26 | 6 | 5475-84. |
| Doshi S, BJ Silk, D Dutt, M Ahmed, AL Cohen, TH Taylor, WA Brooks, D Goswami, SP Luby, AM Fry, PK Ram | Household-level risk factors for influenza among young children in Dhaka, Bangladesh: a case-control study | Tropical Medicine & International Health | 2015 | 20 | 6 | 719-729 |
| Gurley, E.S., Salje, H., Homaira, N., Ram, P.K., Haque, R., Petri Jr, W.A., Bresee, J., Moss, W.J., Luby, S.P., Breysse, P. and Azziz-Baumgartner, E | Seasonal concentrations and determinants of indoor particulate matter in a low-income community in Dhaka, Bangladesh | Environmental research | 2013 | 121 | 0 | ##### |

| **Belgium** | | | | | | |
|---|---|---|---|---|---|---|
| Bentayeb M, D Norback, M Bednarek, A Bernard, GH Cai, S Cerrai, KK Eleftheriou, C Gratziou, GJ Holst, F Lavaud, J Nasilowski, P Sestini, G Sarno, T Sigsgaard, G Wieslander, J Zielinski, G Viegi, I Annesi-Maesano, G Study | Indoor air quality, ventilation and respiratory health in elderly residents Living in nursing homes in Europe | European Respiratory Journal | 2015 | 45 | 5 | 1228-1238 |
| Buczynska AJ, A Krata, R Van Grieken, A Brown, G Polezer, K De Wael, S Potgieter-Vermaak | Composition of PM2.5 and PM1 on high and low pollution event days and its relation to indoor air quality in a home for the elderly | Science of the Total Environment | 2014 | 490 | NA | 134-143 |
| Dons E, LI Panis, M Van Poppel, J Theunis, H Willems, R Torfs, G Wets | Impact of time-activity patterns on personal exposure to black carbon | Atmospheric Environment | 2011 | 45 | 21 | 3594-3602 |

| | | | | | | |
|---|---|---|---|---|---|---|
| Jacobs L, A Buczynska, C Walgraeve, A Delcloo, S Potgieter-Vermaak, R Van Grieken, K Demeestere, J Dewulf, H Van Langenhove, H De Backer, B Nemery, TS Nawrot | Acute changes in pulse pressure in relation to constituents of particulate air pollution in elderly persons | Environmental Research | 2012 | 117 | NA | 60-67 |
| Stranger M, SS Potgieter-Vermaak, R Van Grieken | Particulate matter and gaseous pollutants in residences in Antwerp, Belgium | Science of the Total Environment | 2009 | 407 | 3 | 1182-1192 |

| **Brazil** | | | | | | |
|---|---|---|---|---|---|---|
| Segalin B, P Kumar, K Micadei, A Fornaro, FLT Goncalves | Size-segregated particulate matter inside residences of elderly in the Metropolitan Area of Sao Paulo, Brazil | Atmospheric Environment | 2017 | 148 | NA | 139-151 |

| **Canada** | | | | | | |
|---|---|---|---|---|---|---|
| Akom JB, Sadick AM, Issa MH, Rashwan S, Duhoux M. | The indoor environmental quality performance of green low-income single-family housing. | Journal of Green Building | 2018 | 13 | 2 | 98-120 |
| Allen RW, C Carlsten, B Karlen, S Leckie, S van Eeden, S Vedal, I Wong, M Brauer | An Air Filter Intervention Study of Endothelial Function among Healthy Adults in a Woodsmoke-impacted Community | American Journal of Respiratory and Critical Care Medicine | 2011 | 183 | 9 | 1222-1230 |
| Allen RW, S Leckie, G Millar, M Brauer | The impact of wood stove technology upgrades on indoor residential air quality | Atmospheric Environment | 2009 | 43 | 37 | 5908-5915 |
| Bari MA, M MacNeill, WB Kindzierski, L Wallace, ME Heroux, AJ Wheeler | Predictors of coarse particulate matter and associated endotoxin concentrations in residential environments | Atmospheric Environment | 2014 | 92 | NA | 221-230 |

| | | | | | | |
|---|---|---|---|---|---|---|
| Barn P, T Larson, M Noullett, S Kennedy, R Copes, M Brauer | Infiltration of forest fire and residential wood smoke: an evaluation of air cleaner effectiveness | Journal of Exposure Science and Environmental Epidemiology | 2008 | 18 | 5 | 503-511 |
| Clark NA, RW Allen, P Hystad, L Wallace, SD Dell, R Foty, E Dabek-Zlotorzynska, G Evans, AJ Wheeler | Exploring Variation and Predictors of Residential Fine Particulate Matter Infiltration | International Journal of Environmental Research and Public Health | 2010 | 7 | 8 | 3211-3224 |
| Crump KS | Manganese exposures in Toronto during use of the gasoline additive, methylcyclopentadienyl manganese tricarbonyl | Journal of Exposure Analysis and Environmental Epidemiology | 2000 | 10 | 3 | 227-239 |
| Evans GJ, A Peers, K Sabaliauskas | Particle dose estimation from frying in residential settings | Indoor Air | 2008 | 18 | 6 | 499-510 |
| Georghiou PE, P Blagden, DA Snow, L Winsor, DT Williams | MUTAGENICITY OF INDOOR AIR CONTAINING ENVIRONMENTAL TOBACCO-SMOKE - EVALUATION OF A PORTABLE PM-10 IMPACTOR SAMPLER | Environmental Science & Technology | 1991 | 25 | 8 | 1496-1500 |
| Ghoshdastidar AJ, Z Hu, Y Nazarenko, PA Ariya | Exposure to nanoscale and microscale particulate air pollution prior to mining development near a northern indigenous community in Quebec, Canada | Environ Sci Pollut Res Int | 2018 | 25 | 9 | 8976-8988 |
| Guggisberg M, PA Hessel, D Michaelchuk, M Atiemo | Particulate matter and gaseous contaminants in indoor environments in an isolated northern community | Int J Circumpolar Health | 2003 | 62 | 2 | 120-9 |

| | | | | | | |
|---|---|---|---|---|---|---|
| Heroux ME, N Clark, K Van Ryswyk, R Mallick, NL Gilbert, I Harrison, K Rispler, D Wang, A Anastassopoulos, M Guay, M MacNeill, AJ Wheeler | Predictors of Indoor Air Concentrations in Smoking and Non-Smoking Residences | International Journal of Environmental Research and Public Health | 2010 | 7 | 8 | 3080-3099 |
| Jeong CH, Salehi S, Wu J, North ML, Kim JS, Chow CW, et al. | Indoor measurements of air pollutants in residential houses in urban and suburban areas: Indoor versus ambient concentrations. | Science of The Total Environment | 2019 | 693 | NA | 1E+05 |
| Kajbafzadeh M, M Brauer, B Karlen, C Carlsten, S van Eeden, RW Allen | The impacts of traffic-related and woodsmoke particulate matter on measures of cardiovascular health: a HEPA filter intervention study | Occupational and Environmental Medicine | 2015 | 72 | 6 | 394-400 |
| Kearney J, L Wallace, M MacNeill, ME Heroux, W Kindzierski, A Wheeler | Residential infiltration of fine and ultrafine particles in Edmonton | Atmospheric Environment | 2014 | 94 | NA | 793-805 |
| Kearney J, L Wallace, M MacNeill, X Xu, K VanRyswyk, H You, R Kulka, AJ Wheeler | Residential indoor and outdoor ultrafine particles in Windsor, Ontario | Atmospheric Environment | 2011 | 45 | 40 | 7583-7593 |
| Kovesi T, D Creery, NL Gilbert, R Dales, D Fugler, B Thompson, N Randhawa, JD Miller | Indoor air quality risk factors for severe lower respiratory tract infections in Inuit infants in Baffin Region, Nunavut: a pilot study | Indoor Air | 2006 | 16 | 4 | 266-275 |
| Loo CK, RG Foty, AJ Wheeler, JD Miller, G Evans, DM Stieb, SD Dell | Do questions reflecting indoor air pollutant exposure from a questionnaire predict direct measure of exposure in owner-occupied houses? | Int J Environ Res Public Health | 2010 | 7 | 8 | 3270-97 |

| | | | | | | |
|---|---|---|---|---|---|---|
| MacNeill M, J Kearney, L Wallace, M Gibson, ME Heroux, J Kuchta, JR Guernsey, AJ Wheeler | Quantifying the contribution of ambient and indoor-generated fine particles to indoor air in residential environments | Indoor Air | 2014 | 24 | 4 | 362-375 |
| Miller JD, Dugandzic R, Frescura AM, Salares V | Indoor-and outdoor-derived contaminants in urban and rural homes in Ottawa, Ontario, Canada | Journal of the Air & Waste Management Association | 2007 | 57 | 3 | 297-302 |
| Miller, J.D., Dugandzic, R., Frescura, A.M. and Salares, V. | Indoor- and Outdoor-Derived Contaminants in Urban and Rural Homes in Ottawa, Ontario, Canada | Journal of the Air & Waste Management Association | 2007 | 57 | 7 | 297-302 |
| Pellizzari ED, CA Clayton, CE Rodes, RE Mason, LL Piper, B Fort, G Pfeifer, D Lynam | Particulate matter and manganese exposures in Toronto, Canada | Atmospheric Environment | 1999 | 33 | 5 | 721-734 |
| Rasmussen PE, C Levesque, M Chenier, HD Gardner | Contribution of metals in resuspended dust to indoor and personal inhalation exposures: Relationships between PM10 and settled dust | Building and Environment | 2018 | 143 | NA | 513-522 |
| Smargiassi A, M Baldwin, C Pilger, R Dugandzic, M Brauer | Small-scale spatial variability of particle concentrations and traffic levels in Montreal: a pilot study | Science of the Total Environment | 2005 | 338 | 3 | 243-251 |
| Weichenthal S, A Dufresne, C Infante-Rivard, L Joseph | Indoor ultrafine particle exposures and home heating systems: A cross-sectional survey of Canadian homes during the winter months | Journal of Exposure Science and Environmental Epidemiology | 2007 | 17 | 3 | 288-297 |
| Weichenthal S, G Mallach, R Kulka, A Black, A Wheeler, H You, M St-Jean, R Kwiatkowski, D Sharp | A randomized double-blind crossover study of indoor air filtration and acute changes in cardiorespiratory health in a First Nations community | Indoor Air | 2013 | 23 | 3 | 175-184 |
| Wheeler AJ, LA Wallace, J | Personal, Indoor, and Outdoor | Aerosol Science and Technology | 2011 | 45 | 9 | 1078-1089 |

| | | | | | | |
|---|---|---|---|---|---|---|
| Kearney, K Van Ryswyk, HY You, R Kulka, JR Brook, XH Xu | Concentrations of Fine and Ultrafine Particles Using Continuous Monitors in Multiple Residences | | | | | |
| Wheeler AJ, NA Dobbin, N Lyrette, L Wallace, M Foto, R Mallick, J Kearney, K Van Ryswyk, NL Gilbert, I Harrison, K Rispler, ME Heroux | Residential indoor and outdoor coarse particles and associated endotoxin exposures | Atmospheric Environment | 2011 | 45 | 39 | 7064-7071 |

| **Chile** | | | | | | |
|---|---|---|---|---|---|---|
| Adonis M, L Gil | Indoor air pollution in a zone of extreme poverty of metropolitan Santiago, Chile | Indoor and Built Environment | 2001 | 10 | NA | 138-146 |
| Barraza F, H Jorquera, G Valdivia, LD Montoya | Indoor PM2.5 in Santiago, Chile, spring 2012: Source apportionment and outdoor contributions | Atmospheric Environment | 2014 | 94 | NA | 692-700 |
| Barria RM, M Calvo, P Pino | Indoor air pollution by fine particulate matter in the homes of newborns | Revista Chilena De Pediatria-Chile | 2016 | 87 | 5 | 343-350 |
| Bravo-Linares C, L Ovando-Fuentealba, S Orellana-Donoso, S Gatica, F Klerman, SM Mudge, W Gallardo, JP Pinaud, R Loyola-Sepulveda | Source identification, apportionment and toxicity of indoor and outdoor PM2.5 airborne particulates in a region characterised by wood burning | Environmental Science-Processes & Impacts | 2016 | 18 | 5 | 575-589 |
| Burgos S, P Ruiz, R Koifman | Changes to indoor air quality as a result of relocating families from slums to public housing | Atmospheric Environment | 2013 | 70 | NA | 179-185 |
| Reyes R, Schueftan A, Ruiz C, González AD. | Controlling air pollution in a context of high energy poverty levels in southern Chile: Clean air but colder houses? | Energy Policy | 2019 | 124 | NA | 301-11 |

| | | | | | | |
|---|---|---|---|---|---|---|
| Rojas-Bracho L, HH Suh, P Oyola, P Koutrakis | Measurements of children's exposures to particles and nitrogen dioxide in Santiago, Chile | Science of the Total Environment | 2002 | 287 | 3 | 249-264 |
| Ruiz PA, C Toro, J Caceres, G Lopez, P Oyola, P Koutrakis | Effect of Gas and Kerosene Space Heaters on Indoor Air Quality: A Study in Homes of Santiago, Chile | Journal of the Air & Waste Management Association | 2010 | 60 | 1 | 98-108 |

| **China** | | | | | | |
|---|---|---|---|---|---|---|
| Ai ZT, CM Mak, DJ Cui | On-site measurements of ventilation performance and indoor air quality in naturally ventilated high-rise residential buildings in Hong Kong | Indoor and Built Environment | 2015 | 24 | 2 | 214-224 |
| Brehmer C, Norris C, Barkjohn KK, Bergin MH, Zhang J, Cui X, et al. | The impact of household air cleaners on the oxidative potential of PM2.5 and the role of metals and sources associated with indoor and outdoor exposure. | Environmental Research | 2019 | NA | NA | 1E+05 |
| Cai J, Yu W, Li BZ, Yao RM, Zhang TJW, Guo M, et al. | Particle removal efficiency of a household portable air cleaner in real-world residences: A single-blind cross-over field study. | Energy and Buildings | 2019 | 203 | NA | NA |
| Cao G, Bi J, Ma Z, Shao Z, Wang J. | Seasonal Characteristics of the Chemical Composition of Fine Particles in Residences of Nanjing, China. | International Journal of Environmental Research and Public Health | 2019 | 16 | 6 | E1066 |
| Cao JJ, H Huang, SC Lee, JC Chow, CW Zou, KF Ho, JG Watson | Indoor/Outdoor Relationships for Organic and Elemental Carbon in PM2.5 at Residential Homes in Guangzhou, China | Aerosol and Air Quality Research | 2012 | 12 | 5 | 902-910 |
| Cao JJ, SC Lee, JC Chow, Y Cheng, KF Ho, K Fung, SX Liu, JG Watson | Indoor/outdoor relationships for PM2.5 and associated carbonaceous pollutants at residential | Indoor Air | 2005 | 15 | 3 | 197-204 |

| | | | | | | |
|---|---|---|---|---|---|---|
| | homes in Hong Kong - case study | | | | | |
| Chao CY, EC Cheng | Source apportionment of indoor PM(2.5) and PM(10) in homes | Indoor and Built Environment | 2002 | 11 | 1 | 27-37 |
| Chao CY, KK Wong | Residential indoor PM10 and PM2.5 in Hong kong and the elemental composition | Atmospheric Environment | 2002 | 36 | 2 | 265-277 |
| Chao CY, KK Wong, EC Cheng | Size distribution of indoor particulate matter in 60 homes in Hong Kong | Indoor and Built Environment | 2002 | 11 | 1 | 18-26 |
| Chao CYH, TC Tung | An empirical model for outdoor contaminant transmission into residential buildings and experimental verification | Atmospheric Environment | 2001 | 35 | 9 | 1585-1596 |
| Chao CYH, TCW Tung, J Burnett | Influence of different indoor activities on the indoor particulate levels in residential buildings | Indoor and Built Environment | 1998 | 7 | 2 | 110-121 |
| Chen Y, D Lv, XH Li, TL Zhu | PM2.5-bound phthalates in indoor and outdoor air in Beijing: Seasonal distributions and human exposure via inhalation | Environmental Pollution | 2018 | 241 | NA | 369-377 |
| Chen Y, Zhang HB, Yoshino H, Xie JC, Yanagi U, Hasegawa K, et al. | Winter indoor environment of elderly households: A case of rural regions in northeast and southeast China. | Building and Environment | 2019 | 165 | NA | Epub |
| Chen, Y., Li, X., Zhu, T., Han, Y. and Lv, D., | PM2. 5-bound PAHs in three indoor and one outdoor air in Beijing: Concentration, source and health risk assessment | Science of The Total Environment | 2017 | 586 | NA | 255-264 |
| Cheng YL, M Yan, JL Li, ZR Liu, YH Bai, W Tian, DG Wu, Q Cheng | Variations in indoor PM10 concentrations in sixteen homes in Guiyang City, People's Republic of China | Bulletin of Environmental Contamination and Toxicology | 2006 | 77 | 1 | 112-118 |
| Cheung PK, Jim CY. | Indoor air quality in substandard housing in Hong Kong. | Sustainable Cities and Society | 2019 | 48 | NA | 1E+05 |

| | | | | | | |
|---|---|---|---|---|---|---|
| Chu YY, P Xu, ZW Yang, WL Li | Retrofitting existing buildings to control indoor PM2.5 concentration on smog days: Initial experience of residential buildings in China | Building Services Engineering Research & Technology | 2018 | 39 | 3 | 263-283 |
| Cui X, F Li, J Xiang, L Fang, MK Chung, DB Day, J Mo, CJ Weschler, J Gong, L He, D Zhu, C Lu, H Han, Y Zhang, JJ Zhang | Cardiopulmonary effects of overnight indoor air filtration in healthy non-smoking adults: A double-blind randomized crossover study | Environ Int | 2018 | 114 | NA | 27-36 |
| Dai XL, JJ Liu, XD Li, L Zhao | Long-term monitoring of indoor CO2 and PM2.5 in Chinese homes: Concentrations and their relationships with outdoor environments | Building and Environment | 2018 | 144 | NA | 238-247 |
| Day DB, J Xiang, J Mo, MA Clyde, CJ Weschler, F Li, J Gong, M Chung, Y Zhang, J Zhang | Combined use of an electrostatic precipitator and a high-efficiency particulate air filter in building ventilation systems: Effects on cardiorespiratory health indicators in healthy adults | Indoor Air | 2018 | 177 | 9 | 1344-1353 |
| Deng GF, ZH Li, ZC Wang, JB Gao, ZW Xu, JD Li, ZY Wang | Indoor/outdoor relationship of PM2.5 concentration in typical buildings with and without air cleaning in Beijing | Indoor and Built Environment | 2017 | 26 | 1 | 60-68 |
| Ding A, YJ Yang, ZH Zhao, A Huls, A Vierkotter, ZY Yuan, J Cai, J Zhang, WS Gao, JX Li, MF Zhang, M Matsui, J Krutmann, HD Kan, T Schikowski, L Jin, SJ Wang | Indoor PM2.5 exposure affects skin aging manifestation in a Chinese population | Scientific Reports | 2017 | 7 | NA | NA |

| | | | | | | |
|---|---|---|---|---|---|---|
| Fan GT, JC Xie, HS Yoshino, U Yanagi, K Hasegawa, N Kagi, T Goto, QY Zhang, CY Wang, JP Liu | Indoor environmental conditions in urban and rural homes with older people during heating season: A case in cold region, China | Energy and Buildings | 2018 | 167 | NA | 334-346 |
| Fan GT, JC Xie, JP Liu, H Yoshino | Investigation of indoor environmental quality in urban dwellings with schoolchildren in Beijing, China | Indoor and Built Environment | 2017 | 26 | 5 | 694-716 |
| Han Y, M Qi, Y Chen, H Shen, J Liu, Y Huang, H Chen, W Liu, X Wang, J Liu, B Xing, S Tao | Influences of ambient air PM(2).(5) concentration and meteorological condition on the indoor PM(2).(5) concentrations in a residential apartment in Beijing using a new approach | Environ Pollut | 2015 | 205 | NA | 307-14 |
| Han YJ, XH Li, TL Zhu, D Lv, Y Chen, LA Hou, YP Zhang, MZ Ren | Characteristics and Relationships between Indoor and Outdoor PM2.5 in Beijing: A Residential Apartment Case Study | Aerosol and Air Quality Research | 2016 | 16 | 10 | 2386-2395 |
| Ho KF, JJ Cao, RM Harrison, SC Lee, KK Bau | Indoor/outdoor relationships of organic carbon (OC) and elemental carbon (EC) in PM2.5 in roadside environment of Hong Kong | Atmospheric Environment | 2004 | 38 | 37 | 6327-6335 |
| Hu J, N Li, Y Lv, J Liu, J Xie, H Zhang | Investigation on Indoor Air Pollution and Childhood Allergies in Households in Six Chinese Cities by Subjective Survey and Field Measurements | Int J Environ Res Public Health | 2017 | 14 | 9 | NA |
| Hu JH, NP Li, H Yoshino, U Yanagi, K Hasegawa, N Kagi, YD He, XQ Wei | Field study on indoor health risk factors in households with schoolchildren in south-central China | Building and Environment | 2017 | 117 | NA | 260-273 |

| | | | | | | |
|---|---|---|---|---|---|---|
| Hu YJ, LJ Bao, CL Huang, SM Li, P Liu, EY Zeng | Exposure to air particulate matter with a case study in Guangzhou: Is indoor environment a safe haven in China? | Atmospheric Environment | 2018 | 191 | NA | 351-359 |
| Huang C, X Wang, W Liu, J Cai, L Shen, Z Zou, R Lu, J Chang, X Wei, C Sun, Z Zhao, Y Sun, J Sundell | Household indoor air quality and its associations with childhood asthma in Shanghai, China: On-site inspected methods and preliminary results | Environ Res | 2016 | 151 | NA | 154-167 |
| Huang KL, JS Song, GH Feng, QP Chang, B Jiang, J Wang, W Sun, HX Li, JM Wang, XS Fang | Indoor air quality analysis of residential buildings in northeast China based on field measurements and longtime monitoring | Building and Environment | 2018 | 144 | NA | 171-183 |
| Huang L, Z Pu, M Li, J Sundell | Characterizing the Indoor-Outdoor Relationship of Fine Particulate Matter in Non-Heating Season for Urban Residences in Beijing | PLoS One | 2015 | 10 | 9 | e0138559 |
| Huang MJ, W Wang, HM Leung, CY Chan, WK Liu, MH Wong, KC Cheung | Mercury levels in road dust and household TSP/PM2.5 related to concentrations in hair in Guangzhou, China | Ecotoxicology and Environmental Safety | 2012 | 81 | NA | 27-35 |
| Lai SC, KF Ho, YY Zhang, SC Lee, Y Huang, SC Zou | Characteristics of Residential Indoor Carbonaceous Aerosols: A Case Study in Guangzhou, Pearl River Delta Region | Aerosol and Air Quality Research | 2010 | 10 | 5 | 472-478 |
| Lee SC, M Chang, KY Chan | Indoor and outdoor air quality investigation at six residential buildings in Hong Kong | Environment International | 1999 | 25 | 4 | 489-496 |
| Lee SC, WM Li, CH Ao | Investigation of indoor air quality at residential homes in Hong Kong - case study | Atmospheric Environment | 2002 | 36 | 2 | 225-237 |

| | | | | | | |
|---|---|---|---|---|---|---|
| Li CL, JM Fu, GY Sheng, XH Bi, YM Hao, XM Wang, BX Mai | Vertical distribution of PAHs in the indoor and outdoor PM2.5 in Guangzhou, China | Building and Environment | 2005 | 40 | 3 | 329-341 |
| Li TX, SZ Cao, DL Fan, YQ Zhang, BB Wang, XG Zhao, BP Leaderer, GF Shen, YW Zhang, XL Duan | Household concentrations and personal exposure of PM2.5 among urban residents using different cooking fuels | Science of the Total Environment | 2016 | 548 | NA | 43628 |
| Liu H, ZW Zhang, NJ Wen, C Wang | Determination and risk assessment of airborne endotoxin concentrations in a university campus | Journal of Aerosol Science | 2018 | 115 | NA | 146-157 |
| Liu J, Y Man, Y Liu | Temporal variability of PM10 and PM2.5 inside and outside a residential home during 2014 Chinese Spring Festival in Zhengzhou, China | Natural Hazards | 2014 | 73 | 3 | 2149-2154 |
| Liu JJ, XL Dai, XD Li, SS Jia, JJ Pei, YX Sun, DY Lai, X Shen, HJ Sun, HG Yin, KL Huang, HW Tan, Y Gao, YW Jian | Indoor air quality and occupants' ventilation habits in China: Seasonal measurement and long-term monitoring | Building and Environment | 2018 | 142 | NA | 119-129 |
| Liu S, J Chen, Q Zhao, XM Song, DQ Shao, K Meliefste, YP Du, J Wang, M Wang, T Wang, BH Feng, RS Wu, HB Xu, B He, B Brunekreef, W Huang | Cardiovascular benefits of short-term indoor air filtration intervention in elderly living in Beijing: An extended analysis of BIAPSY study | Environmental Research | 2018 | 167 | NA | 632-638 |
| Liu Z, K Cheng, H Li, G Cao, D Wu, Y Shi | Exploring the potential relationship between indoor air quality and the concentration of airborne culturable fungi: a combined experimental and neural network modeling study | Environ Sci Pollut Res Int | 2018 | 25 | 4 | 3510-3517 |

| | | | | | | |
|---|---|---|---|---|---|---|
| Liu ZJ, AG Li, ZP Hu, HF Sun | Study on the potential relationships between indoor culturable fungi, particle load and children respiratory health in Xi'an, China | Building and Environment | 2014 | 80 | NA | 105-114 |
| Mazaheri M, Lin WW, Clifford S, Yue DL, Zhai YH, Xu MW, et al. | Characteristics of school children's personal exposure to ultrafine particles in Heshan, Pearl River Delta, China - A pilot study. | Environment International | 2019 | 132 | NA | Epub |
| Mullen NA, C Liu, YP Zhang, SX Wang, WW Nazaroff | Ultrafine particle concentrations and exposures in four high-rise Beijing apartments | Atmospheric Environment | 2011 | 45 | 40 | 7574-7582 |
| Niu X, B Guinot, J Cao, H Xu, J Sun | Particle size distribution and air pollution patterns in three urban environments in Xi'an, China | Environ Geochem Health | 2015 | 37 | 5 | 801-12 |
| Niu X, Ho KF, Hu T, Sun J, Duan J, Huang Y, et al. | Characterization of chemical components and cytotoxicity effects of indoor and outdoor fine particulate matter (PM2.5) in Xi'an, China. | Environmental Science and Pollution Research | 2019 | 26 | 31 | 31913-23 |
| Pei JJ, Dong CB, Liu JJ. | Operating behavior and corresponding performance of portable air cleaners in residential buildings, China. | Building and Environment | 2019 | 147 | NA | 473-81 |
| Qi M, X Zhu, W Du, YL Chen, YC Chen, TB Huang, XL Pan, QR Zhong, X Sun, EY Zeng, BS Xing, S Tao | Exposure and health impact evaluation based on simultaneous measurement of indoor and ambient PM2.5 in Haidian, Beijing | Environmental Pollution | 2017 | 220 | NA | 704-712 |
| Shao ZJ, J Bi, ZW Ma, JN Wang | Seasonal trends of indoor fine particulate matter and its determinants in urban residences in Nanjing, China | Building and Environment | 2017 | 125 | NA | 319-325 |

| | | | | | | |
|---|---|---|---|---|---|---|
| Spickett J, K Rumchev, H Jing | The Domestic Environment and Respiratory Health of School Children in Zongshan, China | Asia-Pacific Journal of Public Health | 2014 | 26 | 6 | 596-603 |
| Sun Y, Hou J, Cheng R, Sheng Y, Zhang X, Sundell J. | Indoor air quality, ventilation and their associations with sick building syndrome in Chinese homes. | Energy and Buildings | 2019 | 197 | 15 | 112-9 |
| Tian LW, GQ Zhang, Q Zhang, DJ Moschandreas, JH Hao, JP Lin, YH Liu | The impact of kitchen activities on indoor pollutant concentrations | Indoor and Built Environment | 2008 | 17 | 4 | 377-383 |
| Tong X, Chen XC, Chuang HC, Cao JJ, Ho SSH, Lui KH, et al. | Characteristics and cytotoxicity of indoor fine particulate matter (PM2.5) and PM2.5-bound polycyclic aromatic hydrocarbons (PAHs) in Hong Kong. | Air Quality, Atmosphere & Health | 2019 | 12 | 10 | Epub, Pages 1169-1179 |
| Tong XN, B Wang, WT Dai, JJ Cao, SSH Ho, TCY Kwok, KH Lui, CM Lo, KF Ho | Indoor air pollutant exposure and determinant factors controlling household air quality for elderly people in Hong Kong | Air Quality Atmosphere and Health | 2018 | 11 | 6 | 695-704 |
| Tung TCW, CYH Chao, J Burnett, SW Pang, RYM Lee | A territory wide survey on indoor particulate level in Hong Kong | Building and Environment | 1999 | 34 | 2 | 213-220 |
| Wan MP, CL Wu, GN Szeto, TC Chan, CYH Chao | Ultrafine particles, and PM2.5 generated from cooking in homes | Atmospheric Environment | 2011 | 45 | 34 | 6141-6148 |
| Wang D, Wang P, Wang YW, Zhang WW, Zhu CF, Sun HZ, et al. | Temporal variations of PM2.5-bound organophosphate flame retardants in different microenvironments in Beijing, China, and implications for human exposure. | Science of the Total Environment | 2019 | 666 | NA | 226-34 |
| Wang F, D Meng, X Li, J Tan | Indoor-outdoor relationships of PM2.5 in four residential dwellings in winter in | Environ Pollut | 2016 | 215 | NA | 280-289 |

| | | | | | | |
|---|---|---|---|---|---|---|
| | the Yangtze River Delta, China | | | | | |
| Wang F, YY Zhou, D Meng, MM Han, CQ Jia | Heavy metal characteristics and health risk assessment of PM2.5 in three residential homes during winter in Nanjing, China | Building and Environment | 2018 | 143 | NA | 339-348 |
| Wang JN, Y Zhang | CO and particle pollution of indoor air in Beijing and its elemental analysis | Biomed Environ Sci | 1990 | 3 | 2 | 132-8 |
| Wang ZJ, QW Xue, YC Ji, ZY Yu | Indoor environment quality in a low-energy residential building in winter in Harbin | Building and Environment | 2018 | 135 | NA | 194-201 |
| Wang ZQ, JJ Liu | Spring-time PM2.5 elemental analysis and polycyclic aromatic hydrocarbons measurement in High-rise residential buildings in Chongqing and Xian, China | Energy and Buildings | 2018 | 173 | NA | 623-633 |
| Wu YJ, Li GY, Yang Y, An TC. | Pollution evaluation and health risk assessment of airborne toxic metals in both indoors and outdoors of the Pearl River Delta, China. | Environmental Research | 2019 | 179 | NA | Epub |
| Xiang RB, JB Song, Y Zhu, W Guang | Indoor Air Quality in Kitchens in Rural China | Environmental Engineering Science | 2016 | 33 | 9 | 699-704 |
| Xiao Y, LN Wang, MZ Yu, TT Shui, L Liu, J Liu | Characteristics of indoor/outdoor PM2.5 and related carbonaceous species in a typical severely cold city in China during heating season | Building and Environment | 2018 | 129 | NA | 54-64 |

| | | | | | | |
|---|---|---|---|---|---|---|
| Xie YY, B Zhao | Chemical composition of outdoor and indoor PM2.5 collected during haze events: Transformations and modified source contributions resulting from outdoor-to-indoor transport | Indoor Air | 2018 | 28 | 6 | 828-839 |
| Xu F, W Tang, W Zhang, L Liu, K Lin | Levels, distributions and correlations of polybrominated diphenyl ethers in air and dust of household and workplace in Shanghai, China: implication for daily human exposure | Environ Sci Pollut Res Int | 2016 | 23 | 4 | 3229-38 |
| Yang F, Lau CF, Tong VWT, Zhang KK, Westerdahl D, Ng S, et al. | Assessment of personal integrated exposure to fine particulate matter of urban residents in Hong Kong. | Journal of the Air & Waste Management Association | 2019 | 69 | 1 | 47-57 |
| Yang YB, L Liu, CY Xu, N Li, Z Liu, Q Wang, DQ Xu | Source Apportionment and Influencing Factor Analysis of Residential Indoor PM2.5 in Beijing | International Journal of Environmental Research and Public Health | 2018 | 15 | 4 | NA |
| Yang Z, Shen J, Gao Z. | Ventilation and Air Quality in Student Dormitories in China: A Case Study during Summer in Nanjing. | International journal of environmental research and public health | 2018 | 15 | 7 | Epub |
| Yin H, Liu C, Zhang L, Li A, Ma Z. | Measurement and evaluation of indoor air quality in naturally ventilated residential buildings. | Indoor and Built Environment | 2019 | 28 | 10 | 1307-23 |
| Zhan Y, K Johnson, C Norris, MM Shafer, MH Bergin, Y Zhang, J Zhang, JJ Schauer | The influence of air cleaners on indoor particulate matter components and oxidative potential in residential households in Beijing | Sci Total Environ | 2018 | 626 | NA | 507-518 |

| | | | | | | |
|---|---|---|---|---|---|---|
| Zhang HB, JC Xie, H Yoshino, U Yanagi, K Hasegawa, N Kagi, ZW Lian | Thermal and environmental conditions in Shanghai households: Risk factors for childhood health | Building and Environment | 2016 | 104 | NA | 35-46 |
| Zhang YQ, SZ Cao, XY Xu, J Qiu, MX Chen, D Wang, DH Guan, CY Wang, X Wang, BW Dong, H Huang, N Zhao, L Jin, YN Bai, XL Duan, Q Liu, YW Zhang | Metals compositions of indoor PM2.5, health risk assessment, and birth outcomes in Lanzhou, China | Environmental Monitoring and Assessment | 2016 | 188 | 6 | NA |
| Zhao HY, LY Shao, Q Yao | Microscopic morphology and size distribution of residential indoor PM10 in Beijing City | Indoor and Built Environment | 2005 | 14 | 6 | 513-520 |
| Zhao L, JJ Liu, JL Ren | Impact of various ventilation modes on IAQ and energy consumption in Chinese dwellings: First long-term monitoring study in Tianjin, China | Building and Environment | 2018 | 143 | NA | 99-106 |
| Zhao WC, JP Cheng, ZY Yu, QL Tang, F Cheng, YW Yin, WH Wang | Levels, seasonal variations, and health risks assessment of ambient air pollutants in the residential areas | International Journal of Environmental Science and Technology | 2013 | 10 | 3 | 487-494 |
| Zhou J, B Han, ZP Bai, Y You, JF Zhang, C Niu, YT Liu, N Zhang, F He, X Ding, B Lu, YD Hu | Particle Exposure Assessment for Community Elderly (PEACE) in Tianjin, China: Mass concentration relationships | Atmospheric Environment | 2012 | 49 | NA | 77-84 |
| Zhou R, S Li, Y Zhou, A Haug | Comparison of environmental tobacco smoke concentrations and mutagenicity for several indoor environments | Mutat Res | 2000 | 465 | NA | 191-200 |

| | | | | | | |
|---|---|---|---|---|---|---|
| Zhou, Z., Liu, Y., Yuan, J., Zuo, J., Chen, G., Xu, L. and Rameezdeen, R. | Indoor PM2. 5 concentrations in residential buildings during a severely polluted winter: A case study in Tianjin, China | Renewable and Sustainable Energy Reviews | 2016 | 64 | NA | 372-381 |
| Zhu SW, W Cai, H Yoshino, U Yanagi, K Hasegawa, N Kagi, MQ Chen | Primary pollutants in schoolchildren's homes in Wuhan, China | Building and Environment | 2015 | 93 | NA | 41-53 |

| **Czech Republic** | | | | | | |
|---|---|---|---|---|---|---|
| Branis M, J Kolomaznikova | Monitoring of long-term personal exposure to fine particulate matter (PM2.5) | Air Quality Atmosphere and Health | 2010 | 3 | 4 | 235-243 |
| Branis M, P Rezacova, M Lazaridis | The effect of source type and source strength on inhaled mass of particulate matter during episodic indoor activities | Indoor and Built Environment | 2014 | 23 | 8 | 1106-1116 |
| Braniš, M., Hovorka, J., Rezáccová, P., Domasová, M., & Lazaridis, M. | Effect of Indoor and Outdoor Sources on Particulate Matter Concentration in a Naturally Ventilated Flat (URBAN-AEROSOL Project-Prague) | Indoor Built Environment | 2005 | 14 | 3 | 307-12 |
| Hanninen OO, E Lebret, V Ilacqua, K Katsouyanni, F Kunzli, RJ Sram, M Jantunen | Infiltration of ambient PM2.5 and levels of indoor generated non-ETS PM2.5 in residences of four European cities | Atmospheric Environment | 2004 | 38 | 37 | 6411-6423 |
| Hussein T, T Glytsos, J Ondracek, P Dohanyosova, V Zdimal, K Hameri, M Lazaridis, J Smolik, M Kulmala | Particle size characterization and emission rates during indoor activities in a house | Atmospheric Environment | 2006 | 40 | 23 | 4285-4307 |

| | | | | | | |
|---|---|---|---|---|---|---|
| Kousa A, L Oglesby, K Koistinen, N Kunzli, M Jantunen | Exposure chain of urban air PM2.5 - associations between ambient fixed site, residential outdoor, indoor, workplace and personal exposures in four European cities in the EXPOLIS-study | Atmospheric Environment | 2002 | 36 | 18 | 3031-3039 |
| Lazaridis M, K Eleftheriadis, V Zdimal, J Schwarz, Z Wagner, J Ondracek, Y Drossinos, T Glytsos, S Vratolis, K Torseth, P Moravec, T Hussein, J Smolik | Number Concentrations and Modal Structure of Indoor/Outdoor Fine Particles in Four European Cities | Aerosol and Air Quality Research | 2017 | 17 | 1 | 131-146 |
| Okonski K, C Degrendele, L Melymuk, L Landlova, P Kukucka, S Vojta, J Kohoutek, P Cupr, J Klanova | Particle Size Distribution of Halogenated Flame Retardants and Implications for Atmospheric Deposition and Transport | Environmental Science & Technology | 2014 | 48 | 24 | 14426-14434 |
| Talbot N, L Kubelova, O Makes, J Ondracek, M Cusack, J Schwarz, P Vodicka, N Zikova, V Zdimal | Transformations of Aerosol Particles from an Outdoor to Indoor Environment | Aerosol and Air Quality Research | 2017 | 17 | 3 | 653-665 |
| **Denmark** | | | | | | |
| Beko G, BU Kjeldsen, Y Olsen, J Schipperijn, A Wierzbicka, DG Karottki, J Toftum, S Loft, G Clausen | Contribution of various microenvironments to the daily personal exposure to ultrafine particles: Personal monitoring coupled with GPS tracking | Atmospheric Environment | 2015 | 110 | NA | 122-129 |
| Beko G, CJ Weschler, A Wierzbicka, DG Karottki, J Toftum, S Loft, G Clausen | Ultrafine Particles: Exposure and Source Apportionment in 56 Danish Homes | Environmental Science & Technology | 2013 | 47 | 18 | 10240-10248 |

| | | | | | | |
|---|---|---|---|---|---|---|
| Bentayeb M, D Norback, M Bednarek, A Bernard, GH Cai, S Cerrai, KK Eleftheriou, C Gratziou, GJ Holst, F Lavaud, J Nasilowski, P Sestini, G Sarno, T Sigsgaard, G Wieslander, J Zielinski, G Viegi, I Annesi-Maesano, G Study | Indoor air quality, ventilation and respiratory health in elderly residents Living in nursing homes in Europe | European Respiratory Journal | 2015 | 45 | 5 | 1228-1238 |
| Jantzen K, P Moller, DG Karottki, Y Olsen, G Beko, G Clausen, LG Hersoug, S Loft | Exposure to ultrafine particles, intracellular production of reactive oxygen species in leukocytes and altered levels of endothelial progenitor cells | Toxicology | 2016 | 359 | NA | 43787 |
| Karottki DG, G Beko, G Clausen, AM Madsen, ZJ Andersen, A Massling, M Ketzel, T Ellermann, R Lund, T Sigsgaard, P Moller, S Loft | Cardiovascular and lung function in relation to outdoor and indoor exposure to fine and ultrafine particulate matter in middle-aged subjects | Environment International | 2014 | 73 | NA | 372-381 |
| Karottki DG, M Spilak, M Frederiksen, ZJ Andersen, AM Madsen, M Ketzel, A Massling, L Gunnarsen, P Moller, S Loft | Indoor and Outdoor Exposure to Ultrafine, Fine and Microbiologically Derived Particulate Matter Related to Cardiovascular and Respiratory Effects in a Panel of Elderly Urban Citizens | International Journal of Environmental Research and Public Health | 2015 | 12 | 2 | 1667-1686 |
| Olsen Y, DG Karottki, DM Jensen, G Beko, BU Kjeldsen, G Clausen, LG Hersoug, GJ Holst, A Wierzbicka, T Sigsgaard, A Linneberg, P Moller, S Loft | Vascular and lung function related to ultrafine and fine particles exposure assessed by personal and indoor monitoring: a cross-sectional study | Environmental Health | 2014 | 13 | NA | NA |

| | | | | | | |
|---|---|---|---|---|---|---|
| Raaschou-Nielsen O, M Sorensen, O Hertel, BLK Chawes, N Vissing, K Bonnelykke, H Bisgaard | Predictors of indoor fine particulate matter in infants' bedrooms in Denmark | Environmental Research | 2011 | 111 | 1 | 87-93 |
| Sorensen M, S Loft, HV Andersen, OR Nielsen, LT Skovgaard, LE Knudsen, VNB Ivan, O Hertel | Personal exposure to PM2.5, black smoke and NO2 in Copenhagen: relationship to bedroom and outdoor concentrations covering seasonal variation | Journal of Exposure Analysis and Environmental Epidemiology | 2005 | 15 | 5 | 413-422 |
| Spilak MP, GD Karottki, B Kolarik, M Frederiksen, S Loft, L Gunnarsen | Evaluation of building characteristics in 27 dwellings in Denmark and the effect of using particle filtration units on PM2.5 concentrations | Building and Environment | 2014 | 73 | NA | 55-63 |
| Spilak MP, M Frederiksen, B Kolarik, L Gunnarsen | Exposure to ultrafine particles in relation to indoor events and dwelling characteristics | Building and Environment | 2014 | 74 | NA | 65-74 |

| **Ecuador** | | | | | | |
|---|---|---|---|---|---|---|
| Raysoni AU, RX Armijos, MM Weigel, T Montoya, P Eschanique, M Racines, WW Li | Assessment of indoor and outdoor PM species at schools and residences in a high-altitude Ecuadorian urban center | Environmental Pollution | 2016 | 214 | NA | 668-679 |

| **Egypt** | | | | | | |
|---|---|---|---|---|---|---|
| Abdel-Salam MM | Indoor particulate matter in urban residences of Alexandria, Egypt | J Air Waste Manag Assoc | 2013 | 63 | 8 | 956-62 |
| Abdel-Salam MMM | Indoor Particulate Matter in Different Residential Areas of Alexandria City, Egypt | Indoor and Built Environment | 2012 | 21 | 6 | 857-862 |
| Abdel-Salam MMM | Investigation of PM2.5 and carbon dioxide levels in urban homes | Journal of the Air & Waste Management Association | 2015 | 65 | 8 | 930-936 |

| | | | | | | |
|---|---|---|---|---|---|---|
| Wagdi D, K Tarabieh, MN Abou Zeid | Indoor air quality index for preoccupancy assessment | Air Quality Atmosphere and Health | 2018 | 11 | 4 | 445-458 |

| Ethiopia | | | | | | |
|---|---|---|---|---|---|---|
| Embiale A, Zewge F, Chandravanshi BS, Sahle-Demessie E. | Short-term exposure assessment to particulate matter and total volatile organic compounds in indoor air during cooking Ethiopian sauces (Wot) using electricity, kerosene and charcoal fuels. | Indoor and Built Environment | 2019 | 28 | 8 | 1140-54 |

| Finland | | | | | | |
|---|---|---|---|---|---|---|
| de Hartog JJ, JG Ayres, A Karakatsani, A Analitis, H Brink, K Hameri, RM Harrison, K Katsouyanni, A Kotronarou, I Kavouras, C Meddings, J Pekkanen, G Hoek | Lung function and indicators of exposure to indoor and outdoor particulate matter among asthma and COPD patients | Occupational and Environmental Medicine | 2010 | 67 | 1 | 43506 |
| Du L, T Prasauskas, V Leivo, M Turunen, M Pekkonen, M Kiviste, A Aaltonen, D Martuzevicius, U Haverinen-Shaughnessy | Assessment of indoor environmental quality in existing multi-family buildings in North-East Europe | Environ Int | 2015 | 79 | NA | 74-84 |
| Hanninen OO, E Lebret, V Ilacqua, K Katsouyanni, F Kunzli, RJ Sram, M Jantunen | Infiltration of ambient PM2.5 and levels of indoor generated non-ETS PM2.5 in residences of four European cities | Atmospheric Environment | 2004 | 38 | 37 | 6411-6423 |

| | | | | | | |
|---|---|---|---|---|---|---|
| Hoek G, G Kos, RM Harrison, J de Hartog, K Meliefste, H ten Brink, K Katsouyanni, A Karakatsani, M Lianou, A Kotronarou, I Kavouras, J Pekkanen, M Vallius, M Kulmala, A Puustinen, S Thomas, C Meddings, J Ayres, J van Wijnen, K Hameri | Indoor-outdoor relationships of particle number and mass in four European cities | Atmospheric Environment | 2008 | 42 | 1 | 156-169 |
| Hussein T, K Hameri, MSA Heikkinen, M Kulmala | Indoor and outdoor particle size characterization at a family house in Espoo-Finland | Atmospheric Environment | 2005 | 39 | 20 | 3697-3709 |
| Janssen NAH, T Lanki, G Hoek, M Vallius, JJ de Hartog, R Van Grieken, J Pekkanen, B Brunekreef | Associations between ambient, personal, and indoor exposure to fine particulate matter constituents in Dutch and Finnish panels of cardiovascular patients | Occupational and Environmental Medicine | 2005 | 62 | 12 | 868-877 |
| Janssen, N. A., de Hartog, J. J., Hoek, G., Brunekreef, B., Lanki, T., Timonen, K. L., & Pekkanen, J | Personal Exposure to Fine Particulate Matter in Elderly Subjects: Relation between Personal, Indoor, and Outdoor Concentrations | Journal of the Air & Waste Management Association | 2000 | 50 | 7 | 1133-1143 |
| Kousa A, L Oglesby, K Koistinen, N Kunzli, M Jantunen | Exposure chain of urban air PM2.5 - associations between ambient fixed site, residential outdoor, indoor, workplace and personal exposures in four European cities in the EXPOLIS-study | Atmospheric Environment | 2002 | 36 | 18 | 3031-3039 |

| | | | | | | |
|---|---|---|---|---|---|---|
| Montagne, D., Hoek, G., Nieuwenhuijsen, M., Lanki, T., Siponen, T., Portella, M., Meliefste, K. and Brunekreef, B | Temporal associations of ambient PM2. 5 elemental concentrations with indoor and personal concentrations | Atmospheric Environment | 2014 | 86 | 0 | 203-211 |
| Siponen T, Yli-Tuomi T, Tiittanen P, Taimisto P, Pekkanen J, Salonen RO, et al. | Wood stove use and other determinants of personal and indoor exposures to particulate air pollution and ozone among elderly persons in a Northern Suburb. | Indoor Air: Interational Journal of Indoor Environment and Health | 2019 | 29 | 3 | 413-22 |
| Sippula O, H Rintala, M Happo, P Jalava, K Kuuspalo, A Viren, A Leskinen, A Markkanen, M Komppula, P Markkanen, K Lehtinen, J Jokiniemi, MR Hirvonen | Characterization of Chemical and Microbial Species from Size-Segregated Indoor and Outdoor Particulate Samples | Aerosol and Air Quality Research | 2013 | 13 | 4 | 1212-U343 |
| Toivola M, A Nevalainen, S Alm | Personal exposures to particles and microbes in relation to microenvironmental concentrations | Indoor Air | 2004 | 14 | 5 | 351-359 |
| Wierzbicka A, M Bohgard, JH Pagels, A Dahl, J Londahl, T Hussein, E Swietlicki, A Gudmundsson | Quantification of differences between occupancy and total monitoring periods for better assessment of exposure to particles in indoor environments | Atmospheric Environment | 2015 | 106 | NA | 419-428 |
| Yli-Tuomi T, T Lanki, G Hoek, B Brunekreef, J Pekkanen | Determination of the sources of indoor PM(2.5) in Amsterdam and Helsinki | Environmental Science & Technology | 2008 | 42 | 12 | 4440-4446 |

**France**

| | | | | | | |
|---|---|---|---|---|---|---|
| Audi C, N Baiz, CN Maesano, O Ramousse, D Reboulleau, A Magnan, D Caillaud, I Annesi-Maesano | Serum cytokine levels related to exposure to volatile organic compounds and PM2.5 in dwellings and workplaces in French farmers - a mechanism to explain nonsmoking COPD | International Journal of Chronic Obstructive Pulmonary Disease | 2017 | 12 | NA | 1363-1374 |
| Bentayeb M, D Norback, M Bednarek, A Bernard, GH Cai, S Cerrai, KK Eleftheriou, C Gratziou, GJ Holst, F Lavaud, J Nasilowski, P Sestini, G Sarno, T Sigsgaard, G Wieslander, J Zielinski, G Viegi, I Annesi-Maesano, G Study | Indoor air quality, ventilation and respiratory health in elderly residents Living in nursing homes in Europe | European Respiratory Journal | 2015 | 45 | 5 | 1228-1238 |
| Derbez M, G Wyart, E Le Ponner, O Ramalho, J Riberon, C Mandin | Indoor air quality in energy-efficient dwellings: Levels and sources of pollutants | Indoor Air | 2018 | 28 | 2 | 318-338 |
| Derbez, M, B Bruno, C Valerie, L Murielle, P Cecile, R Jacques, K Severine | Indoor air quality and comfort in seven newly built, energy-efficient houses in France | Building and Environment | 2014 | 72 | NA | 173-187 |
| Gauvin S, P Reungoat, S Cassadou, J Dechenaux, I Momas, J Just, D Zmirou | Contribution of indoor and outdoor environments to PM2.5 personal exposure of children - VESTA study | Science of the Total Environment | 2002 | 297 | NA | 175-181 |
| Hulin M, D Caillaud, I Annesi-Maesano | Indoor air pollution and childhood asthma: variations between urban and rural areas | Indoor Air | 2010 | 20 | 6 | 502-514 |
| Langer S, O Ramalho, M Derbez, J Riberon, S Kirchner, C Mandin | Indoor environmental quality in French dwellings and building characteristics | Atmospheric Environment | 2016 | 128 | NA | 82-91 |

| | | | | | | |
|---|---|---|---|---|---|---|
| Maesano CN, Caillaud D, Youssouf H, Banerjee S, Prud'Homme J, Audi C, et al. | Indoor exposure to particulate matter and volatile organic compounds in dwellings and workplaces and respiratory health in French farmers. | Multidisciplinary Respiratory Medicine | 2019 | 14 | 1 | eCollection 2019 |
| Mosqueron L, I Momas, Y Le Moullec | Personal exposure of Paris office workers to nitrogen dioxide and fine particles | Occupational and Environmental Medicine | 2002 | 59 | 8 | 550-555 |
| Nerriere E, D Zmirou-Navier, O Blanchard, I Momas, J Ladner, Y Le Moullec, MB Personnaz, P Lameloise, W Delmas, A Target, H Desqueyroux | Can we use fixed ambient air monitors to estimate population long-term exposure to air pollutants? The case of spatial variability in the Genotox ER study | Environmental Research | 2005 | 97 | 1 | 32-42 |
| Paunescu AC, M Attoui, S Bouallala, J Sunyer, I Momas | Personal measurement of exposure to black carbon and ultrafine particles in schoolchildren from PARIS cohort (Paris, France) | Indoor Air | 2017 | 27 | 4 | 766-779 |
| Ramalho O, G Wyart, C Mandin, P Blondeau, PA Cabanes, N Leclerc, JU Mullot, G Boulanger, M Redaelli | Association of carbon dioxide with indoor air pollutants and exceedance of health guideline values | Building and Environment | 2015 | 93 | NA | 115-124 |
| **Germany** | | | | | | |
| Berger-Preiss E, K Levsen, G Leng, H Idel, D Sugiri, U Ranft | Indoor pyrethroid exposure in homes with woollen textile floor coverings | Int J Hyg Environ Health | 2002 | 205 | 6 | 459-72 |
| Franck U, O Herbarth, S Roder, U Schlink, M Borte, U Diez, U Kramer, I Lehmann | Respiratory effects of indoor particles in young children are size dependent | Science of the Total Environment | 2011 | 409 | 9 | 1621-1631 |

| | | | | | | |
|---|---|---|---|---|---|---|
| Fromme H, T Lahrz, A Hainsch, A Oddoy, M Piloty, H Ruden | Elemental carbon and respirable particulate matter in the indoor air of apartments and nursery schools and ambient air in Berlin (Germany) | Indoor Air | 2005 | 15 | 5 | 335-341 |
| Moriske HJ, M Drews, G Ebert, G Menk, C Scheller, M Schondube, L Konieczny | Indoor air pollution by different heating systems: Coal burning, open fireplace and central heating | Toxicology Letters | 1996 | 88 | NA | 349-354 |
| Muller J | INDOOR AND OUTDOOR AIR MEASUREMENTS AT A CITY STREET WITH HEAVY TRAFFIC | Staub Reinhaltung Der Luft | 1991 | 51 | 4 | 147-154 |
| Oeder S, S Dietrich, I Weichenmeier, W Schober, G Pusch, RA Jorres, R Schierl, D Nowak, H Fromme, H Behrendt, JTM Buters | Toxicity and elemental composition of particulate matter from outdoor and indoor air of elementary schools in Munich, Germany | Indoor Air | 2012 | 22 | 2 | 148-158 |
| Phillips K, DA Howard, MC Bentley, G Alvan | Measured exposures by personal monitoring for respirable suspended particles and environmental tobacco smoke of housewives and office workers resident in Bremen, Germany | Int Arch Occup Environ Health | 1998 | 71 | 3 | 201-12 |
| Salthammer T, T Schripp, S Wientzek, M Wensing | Impact of operating wood-burning fireplace ovens on indoor air quality | Chemosphere | 2014 | 103 | NA | 205-211 |
| Santen M, M Wesselmann, U Fittschen, R Cremer, P Braun, A Ludecke, HJ Moriske | Measurements of fine and ultrafine particles in indoor environment of living rooms | Gefahrstoffe Reinhaltung Der Luft | 2009 | 69 | 3 | 63-70 |

| | | | | | | |
|---|---|---|---|---|---|---|
| Weschler CJ, T Salthammer, H Fromme | Partitioning of phthalates among the gas phase, airborne particles and settled dust in indoor environments | Atmospheric Environment | 2008 | 42 | 7 | 1449-1460 |

| **Greece** | | | | | | |
|---|---|---|---|---|---|---|
| Assimakopoulos VD, T Bekiari, S Pateraki, T Maggos, P Stamatis, P Nicolopoulou, MN Assimakopoulos | Assessing personal exposure to PM using data from an integrated indoor-outdoor experiment in Athens-Greece | Science of the Total Environment | 2018 | 636 | NA | 1303-1320 |
| Bentayeb M, D Norback, M Bednarek, A Bernard, GH Cai, S Cerrai, KK Eleftheriou, C Gratziou, GJ Holst, F Lavaud, J Nasilowski, P Sestini, G Sarno, T Sigsgaard, G Wieslander, J Zielinski, G Viegi, I Annesi-Maesano, G Study | Indoor air quality, ventilation and respiratory health in elderly residents Living in nursing homes in Europe | European Respiratory Journal | 2015 | 45 | 5 | 1228-1238 |
| de Hartog JJ, JG Ayres, A Karakatsani, A Analitis, H Brink, K Hameri, RM Harrison, K Katsouyanni, A Kotronarou, I Kavouras, C Meddings, J Pekkanen, G Hoek | Lung function and indicators of exposure to indoor and outdoor particulate matter among asthma and COPD patients | Occupational and Environmental Medicine | 2010 | 67 | 1 | 43506 |
| Diapouli E, A Chaloulakou, N Spyrellis | Levels of ultrafine particles in different microenvironments - Implications to children exposure | Science of the Total Environment | 2007 | 388 | NA | 128-136 |
| Diapouli E, A Chaloulakou, N Spyrellis | Indoor and outdoor PM concentrations at a residential environment, in the Athens area | Global Nest Journal | 2008 | 10 | 2 | 201-208 |

| | | | | | | |
|---|---|---|---|---|---|---|
| Diapouli E, A Chaloulakou, N Spyrellis | INDOOR/OUTDOOR PM LEVELS AND EC SURROGATE, AT TYPICAL MICROENVIRONMENTS IN THE ATHENS AREA | Global Nest Journal | 2010 | 12 | 1 | 43818 |
| Diapouli E, K Eleftheriadis, AA Karanasiou, S Vratolis, O Hermansen, I Colbeck, M Lazaridis | Indoor and Outdoor Particle Number and Mass Concentrations in Athens. Sources, Sinks and Variability of Aerosol Parameters | Aerosol and Air Quality Research | 2011 | 11 | 6 | 632-642 |
| Hanninen OO, E Lebret, V Ilacqua, K Katsouyanni, F Kunzli, RJ Sram, M Jantunen | Infiltration of ambient PM2.5 and levels of indoor generated non-ETS PM2.5 in residences of four European cities | Atmospheric Environment | 2004 | 38 | 37 | 6411-6423 |
| Hoek G, G Kos, RM Harrison, J de Hartog, K Meliefste, H ten Brink, K Katsouyanni, A Karakatsani, M Lianou, A Kotronarou, I Kavouras, J Pekkanen, M Vallius, M Kulmala, A Puustinen, S Thomas, C Meddings, J Ayres, J van Wijnen, K Hameri | Indoor-outdoor relationships of particle number and mass in four European cities | Atmospheric Environment | 2008 | 42 | 1 | 156-169 |
| Kousa A, L Oglesby, K Koistinen, N Kunzli, M Jantunen | Exposure chain of urban air PM2.5 - associations between ambient fixed site, residential outdoor, indoor, workplace and personal exposures in four European cities in the EXPOLIS-study | Atmospheric Environment | 2002 | 36 | 18 | 3031-3039 |
| Lazaridis M, K Eleftheriadis, V Zdimal, J Schwarz, Z Wagner, J Ondracek, Y Drossinos, T | Number Concentrations and Modal Structure of Indoor/Outdoor Fine Particles in Four European Cities | Aerosol and Air Quality Research | 2017 | 17 | 1 | 131-146 |

| | | | | | | |
|---|---|---|---|---|---|---|
| Glytsos, S Vratolis, K Torseth, P Moravec, T Hussein, J Smolik | | | | | | |
| Mueller W, Steinle S, Parkka J, Parmes E, Liedes H, Kuijpers E, et al. | Urban greenspace and the indoor environment: Pathways to health via indoor particulate matter, noise, and road noise annoyance. | Environmental Research | 2020 | 180 | NA | 1E+05 |
| Saraga DE, T Maggos, CG Helmis, J Michopoulos, JG Bartzis, C Vasilakos | PM1 and PM2.5 ionic composition and VOCs measurements in two typical apartments in Athens, Greece: investigation of smoking contribution to indoor air concentrations | Environmental Monitoring and Assessment | 2010 | 167 | NA | 321-331 |
| Saraga DE, T Maggos, DA Missia, EI Tolis, C Vasilakos, JG Bartzis | SECONDARY ORGANIC PARTICLES FORMATION FROM OZONE-TERPENES REACTION: A CASE STUDY IN A RESIDENCE OF A MEDITERRANEAN CITY | Fresenius Environmental Bulletin | 2010 | 19 | NA | 2019-2025 |
| Seleventi MK, DE Saraga, CG Helmis, K Bairachtari, C Vasilakos, T Maggos | PM2.5 INDOOR/OUTDOOR RELATIONSHIP AND CHEMICAL COMPOSITION IN IONS AND OC/EC IN AN APARTMENT IN THE CENTER OF ATHENS | Fresenius Environmental Bulletin | 2012 | 21 | 11 | 3177-3183 |
| Stamatelopoulou A, Asimakopoulos DN, Maggos T. | Effects of PM, TVOCs and comfort parameters on indoor air quality of residences with young children. | Building and Environment | 2019 | 150 | NA | 233-44 |

| **Hungary** | | | | | | |
|---|---|---|---|---|---|---|
| Szirtesi K, A Angyal, Z Szoboszlai, E Furu, Z Torok, T Igaz, Z Kertesz | Airborne Particulate Matter: An Investigation of Buildings with Passive House Technology in Hungary | Aerosol and Air Quality Research | 2018 | 18 | 5 | 1282-1293 |

| India | | | | | | |
|---|---|---|---|---|---|---|
| Curto A, Donaire-Gonzalez D, Barrera-Gómez J, Marshall JD, Nieuwenhuijsen MJ, Wellenius GA, et al. | Performance of low-cost monitors to assess household air pollution. | Environmental Research | 2018 | 163 | NA | 53-63 |
| Elf JL, Kinikar A, Khadse S, Mave V, Suryavanshi N, Gupte N, et al. | Sources of household air pollution and their association with fine particulate matter in low-income urban homes in India article. | Journal of Exposure Science and Environmental Epidemiology | 2018 | 28 | 4 | 400-10 |
| Gupta S, A Srivastava, VK Jain | Particle size distribution of aerosols and associated heavy metals in kitchen environments | Environ Monit Assess | 2008 | 142 | NA | 141-8 |
| Khillare PS, R Pandey, S Balachandran | Characterisation of indoor PM10 in residential areas of Delhi | Indoor and Built Environment | 2004 | 13 | 2 | 139-147 |
| Kulshreshtha P, M Khare, P Seetharaman | Indoor air quality assessment in and around urban slums of Delhi city, India | Indoor Air | 2008 | 18 | 6 | 488-98 |
| Kulshrestha A, DD Massey, J Masih, A Taneja | Source Characterization of Trace Elements in Indoor Environments at Urban, Rural and Roadside Sites in a Semi Arid Region of India | Aerosol and Air Quality Research | 2014 | 14 | 6 | 1738-1751 |
| Kulshrestha A, DS Bisht, J Masih, D Massey, S Tiwari, A Taneja | Chemical characterization of water-soluble aerosols in different residential environments of semi aridregion of India | Journal of Atmospheric Chemistry | 2009 | 62 | 2 | 121-138 |
| Kumar P | Characterisation of indoor respirable dust in a locality of Delhi, India | Indoor and Built Environment | 2001 | 10 | 2 | 95-102 |
| Lawrence A, N Fatima | Urban air pollution & its assessment in Lucknow City - The second largest city of North India | Science of the Total Environment | 2014 | 488 | NA | 449-457 |

| | | | | | | |
|---|---|---|---|---|---|---|
| Massey D, A Kulshrestha, J Masih, A Taneja | Seasonal trends of PM10, PM5.0, PM2.5 & PM1.0 in indoor and outdoor environments of residential homes located in North-Central India | Building and Environment | 2012 | 47 | NA | 223-231 |
| Massey D, J Masih, A Kulshrestha, M Habil, A Teneja | Indoor/outdoor relationship of fine particles less than 2.5 mu m (PM2.5) in residential homes locations in central Indian region | Building and Environment | 2009 | 44 | 10 | 2037-2045 |
| Parikh J, K Balakrishnan, V Laxmi, H Biswas | Exposure from cooking with biofuels: pollution monitoring and analysis for rural Tamil Nadu, India | Energy | 2001 | 26 | 10 | 949-962 |
| Priyamvada H, C Priyanka, RK Singh, M Akila, R Ravikrishna, SS Gunthe | Assessment of PM and bioaerosols at diverse indoor environments in a southern tropical Indian region | Building and Environment | 2018 | 137 | NA | 215-225 |
| Roy R, R Jan, S Yadav, MH Vasave, PG Satsangi | Study of metals in radical-mediated toxicity of particulate matter in indoor environments of Pune, India | Air Quality Atmosphere and Health | 2016 | 9 | 6 | 669-680 |
| Saksena S, RK Prasad, VR Shankar | Daily exposure to air pollutants in indoor, outdoor and in-vehicle microenvironments: A pilot study in Delhi | Indoor and Built Environment | 2007 | 16 | 1 | 39-46 |
| Satsangi PG, S Yadav, AS Pipal, N Kumbhar | Characteristics of trace metals in fine (PM2.5) and inhalable (PM10) particles and its health risk assessment along with in-silico approach in indoor environment of India | Atmospheric Environment | 2014 | 92 | NA | 384-393 |
| Singh P, R Saini, A Taneja | Physicochemical characteristics of PM2.5: Low, middle, and high-income group | Atmospheric Pollution Research | 2014 | 5 | 3 | 352-360 |

| | | | | | | |
|---|---|---|---|---|---|---|
| | homes in Agra, India-a case study | | | | | |
| Srivastava A, S Gupta, VK Jain | Winter-time size distribution and source apportionment of total suspended particulate matter and associated metals in Delhi | Atmospheric Research | 2009 | 92 | 1 | 88-99 |

| **Indonesia** | | | | | | |
|---|---|---|---|---|---|---|
| Azhar K, I Dharmayanti, I Mufida | The Indoor Average Level of PM2,5 and ARI Among Children Under Five in Kelurahan Kayuringin Jaya, Kota Bekasi 2014 | Media Penelitian Dan Pengembangan Kesehatan | 2016 | 26 | 1 | 45-52 |
| Pramitha E, Haryanto B. | Effect of Exposure to 2.5 mum Indoor Particulate Matter on Adult Lung Function in Jakarta. | Osong Public Health and Research Perspectives | 2019 | 10 | 2 | 51-5 |

| **Iran** | | | | | | |
|---|---|---|---|---|---|---|
| Hassanvand MS, K Naddafi, H Kashani, S Faridi, N Kunzli, R Nabizadeh, F Momeniha, A Gholampour, M Arhami, A Zare, Z Pourpak, M Hoseini, M Yunesian | Short-term effects of particle size fractions on circulating biomarkers of inflammation in a panel of elderly subjects and healthy young adults | Environmental Pollution | 2017 | 223 | NA | 695-704 |
| Hassanvand MS, K Naddafi, S Faridi, M Arhami, R Nabizadeh, MH Sowlat, Z Pourpak, N Rastkari, F Momeniha, H Kashani, A Gholampour, S Nazmara, M Alimohammadi, G Goudarzi, M Yunesian | Indoor/outdoor relationships of PM10, PM2.5, and PM1 mass concentrations and their water-soluble ions in a retirement home and a school dormitory | Atmospheric Environment | 2014 | 82 | NA | 375-382 |

| | | | | | | |
|---|---|---|---|---|---|---|
| Mirhoseini SH, M Nikaeen, K Satoh, K Makimura | Assessment of Airborne Particles in Indoor Environments: Applicability of Particle Counting for Prediction of Bioaerosol Concentrations | Aerosol and Air Quality Research | 2016 | 16 | 8 | 1903-1910 |
| **Ireland** | | | | | | |
| Broderick A, M Byrne, S Armstrong, J Sheahan, AM Coggins | A pre and post evaluation of indoor air quality, ventilation, and thermal comfort in retrofitted co-operative social housing | Building and Environment | 2017 | 122 | NA | 126-133 |
| Galea KS, JF Hurley, H Cowie, AL Shafrir, AS Jimenez, S Semple, JG Ayres, M Coggins | Using PM2.5 concentrations to estimate the health burden from solid fuel combustion, with application to Irish and Scottish homes | Environmental Health | 2013 | 12 | NA | NA |
| Guo L, JO Lewis, JP McLaughlin | Emissions from Irish domestic fireplaces and their impact on indoor air quality when used as supplementary heating source | Global Nest Journal | 2008 | 10 | 2 | 209-216 |
| Semple, S., Garden, C., Coggins, M., Galea, K.S., Whelan, P., Cowie, H., Sánchez-Jiménez, A., Thorne, P.S., Hurley, J.F. and Ayres, J.G | Contribution of solid fuel, gas combustion, or tobacco smoke to indoor air pollutant concentrations in Irish and Scottish homes | Indoor air | 2012 | 22 | 3 | 212-223 |
| **Israel** | | | | | | |
| Dobson R, Rosen LJ, Semple S. | Monitoring secondhand tobacco smoke remotely in real-time: A simple low-cost approach. | Tobacco Induced Diseases | 2019 | 17 | NA | 18 |

| | | | | | | |
|---|---|---|---|---|---|---|
| Jodeh S, AR Hasan, J Amarah, F Judeh, R Salghi, H Lgaz, W Jodeh | Indoor and outdoor air quality analysis for the city of Nablus in Palestine: seasonal trends of PM10, PM5.0, PM2.5, and PM1.0 of residential homes | Air Quality Atmosphere and Health | 2018 | 11 | 2 | 229-237 |
| Krasnov H, I Katra, MD Friger | Insights into Indoor/Outdoor PM Concentration Ratios due to Dust Storms in an Arid Region | Atmosphere | 2015 | 6 | 7 | 879-890 |
| Krasnov H, I Katra, V Novack, A Vodonos, MD Friger | Increased indoor PM concentrations controlled by atmospheric dust events and urban factors | Building and Environment | 2015 | 87 | NA | 169-176 |
| Rosen, L., Zucker, D., Hovell, M., Brown, N., Ram, A. and Myers, V. | Feasibility of Measuring Tobacco Smoke Air Pollution in Homes: Report from a Pilot Study | International journal of environmental research and public health | 2015 | 12 | 12 | 15129-15142 |

| **Italy** | | | | | | |
|---|---|---|---|---|---|---|
| Bentayeb M, D Norback, M Bednarek, A Bernard, GH Cai, S Cerrai, KK Eleftheriou, C Gratziou, GJ Holst, F Lavaud, J Nasilowski, P Sestini, G Sarno, T Sigsgaard, G Wieslander, J Zielinski, G Viegi, I Annesi-Maesano, G Study | Indoor air quality, ventilation and respiratory health in elderly residents Living in nursing homes in Europe | European Respiratory Journal | 2015 | 45 | 5 | 1228-1238 |
| Buonanno G, S Marini, L Morawska, FC Fuoco | Individual dose and exposure of Italian children to ultrafine particles | Science of the Total Environment | 2012 | 438 | NA | 271-277 |

| | | | | | | |
|---|---|---|---|---|---|---|
| Cattaneo A, C Peruzzo, G Garramone, P Urso, R Ruggeri, P Carrer, DM Cavallo | Airborne particulate matter and gaseous air pollutants in residential structures in Lodi province, Italy | Indoor Air | 2011 | 21 | 6 | 489-500 |
| Drago G, C Perrino, S Canepari, S Ruggieri, L L'Abbate, V Longo, P Colombo, D Frasca, M Balzan, G Cuttitta, G Scaccianoce, G Piva, S Bucchieri, M Melis, G Viegi, F Cibella, RCP Grp | Relationship between domestic smoking and metals and rare earth elements concentration in indoor PM2.5 | Environmental Research | 2018 | 165 | NA | 71-80 |
| Frasca D, Marcoccia M, Tofful L, Simonetti G, Perrino C, Canepari S. | Influence of advanced wood-fired appliances for residential heating on indoor air quality. | Chemosphere | 2018 | 211 | NA | 62-71 |
| Manigrasso M, Protano C, Astolfi ML, Massimi L, Avinod P, Vitali M, et al. | Evidences of copper nanoparticle exposure in indoor environments: Long-term assessment, high-resolution field emission scanning electron microscopy evaluation, in silico respiratory dosimetry study and possible health implications. | Science of the Total Environment | 2019 | 653 | NA | Epub |
| Perrino C, L Tofful, S Canepari | Chemical characterization of indoor and outdoor fine particulate matter in an occupied apartment in Rome, Italy | Indoor Air | 2016 | 26 | 4 | 558-570 |
| Romagnoli P, C Balducci, M Perilli, M Gherardi, A Gordiani, C Gariazzo, MP Gatto, A Cecinato | Indoor PAHs at schools, homes and offices in Rome, Italy | Atmospheric Environment | 2014 | 92 | NA | 51-59 |
| Romagnoli, P.; Balducci, C.; Perilli, M.; Vichi, F.; | Indoor air quality at life and work environments in Rome, Italy | Environmental Science and Pollution Research | 2016 | 23 | NA | 3503-3516 |

| | | | | | | |
|---|---|---|---|---|---|---|
| Imperiali, A.; Cecinato, A | | | | | | |
| Scapellato ML, C Canova, A de Simone, M Carrieri, P Maestrelli, L Simonato, GB Bartolucci | Personal PM10 exposure in asthmatic adults in Padova, Italy: seasonal variability and factors affecting individual concentrations of particulate matter | International Journal of Hygiene and Environmental Health | 2009 | 212 | 6 | 626-636 |
| Simoni M, A Scognamiglio, L Carrozzi, S Baldacci, A Angino, F Pistelli, F Di Pede, G Viegi | Indoor exposures and acute respiratory effects in two general population samples from a rural and an urban area in Italy | Journal of Exposure Analysis and Environmental Epidemiology | 2004 | 14 | NA | S144-S152 |
| Simoni M, L Carrozzi, S Baldacci, A Scognamiglio, F Di Pede, T Sapigni, G Viegi | The Po River Delta (North Italy) indoor epidemiological study: Effects of pollutant exposure on acute respiratory symptoms and respiratory function in adults | Archives of Environmental Health | 2002 | 57 | 2 | 130-136 |
| Zauli-Sajani S, S Rovelli, A Trentini, D Bacco, S Marchesi, F Scotto, C Zigola, P Lauriola, DM Cavallo, V Poluzzi, A Cattaneo, O Hanninen | Higher health effects of ambient particles during the warm season: The role of infiltration factors | Science of the Total Environment | 2018 | 627 | NA | 67-77 |

| **Japan** | | | | | | |
|---|---|---|---|---|---|---|
| Ando M, K Tamura, M Matsumoto | [The suspended particulate matter (SPM) and polycyclic aromatic hydrocarbons (PAHs) in indoor and outdoor air along a main road] | Nihon Eiseigaku Zasshi | 1990 | 45 | 5 | 1007-13 |
| Funasaka K, T Miyazaki, K Tsuruho, K Tamura, T Mizuno, K Kuroda | Relationship between indoor and outdoor carbonaceous particulates in roadside households | Environmental Pollution | 2000 | 110 | 1 | 127-134 |

| | | | | | | |
|---|---|---|---|---|---|---|
| Kanatani K, M Okumura, S Tohno, Y Adachi, K Sato, T Nakayama | Indoor particle counts during Asian dust events under everyday conditions at an apartment in Japan | Environmental Health and Preventive Medicine | 2014 | 19 | 1 | 81-88 |
| Kang Y, K Nagano | Field measurement of indoor air quality and airborne microbes in a near-zero energy house with an earth tube in the cold region of Japan | Science and Technology for the Built Environment | 2016 | 22 | 7 | 1010-1023 |
| Ma CJ, GU Kang, CH Kang | Properties of Indoor Particles Collected in Japanese Homes | Asian Journal of Atmospheric Environment | 2015 | 9 | 1 | 31-38 |
| Michikawa T, S Nakai, H Nitta, K Tamura | Validity of using annual mean particulate matter concentrations as measured at fixed site in assessing personal exposure: An exposure assessment study in Japan | Science of the Total Environment | 2014 | 466 | NA | 673-680 |
| Nakai S, H Nitta, M Ono, K Abe, M Sakaguchi | Measurements of biological contaminants and particulate matter inside a dwelling in Japan | Indoor Air | 1999 | 9 | 1 | 41-46 |
| Ohura T, T Noda, T Amagai, M Fusaya | Prediction of personal exposure to PM2.5 and carcinogenic polycyclic aromatic hydrocarbons by their concentrations in residential microenvironments | Environmental Science & Technology | 2005 | 39 | 15 | 5592-5599 |
| Sugiyama T, T Amagai, H Matsushita, M Soma | Size distribution of indoor airborne particulates collected by a low-flow rate cascade impactor | Indoor and Built Environment | 1999 | 8 | 6 | 361-369 |
| Yoda Y, K Tamura, M Shima | Airborne endotoxin concentrations in indoor and outdoor particulate matter and their predictors in an urban city | Indoor Air | 2017 | 27 | 5 | 955-964 |

| Kuwait | | | | | | |
|---|---|---|---|---|---|---|
| Yassin MF, BEY AlThaqeb, EAE Al-Mutiri | Assessment of indoor PM2.5 in different residential environments | Atmospheric Environment | 2012 | 56 | NA | 65-68 |

| Lebanon | | | | | | |
|---|---|---|---|---|---|---|
| Saliba NA, M Atallah, G Al-Kadamany | Levels and indoor-outdoor relationships of PM10 and soluble inorganic ions in Beirut, Lebanon | Atmospheric Research | 2009 | 92 | 1 | 131-137 |

| Lithuania | | | | | | |
|---|---|---|---|---|---|---|
| Du L, T Prasauskas, V Leivo, M Turunen, M Pekkonen, M Kiviste, A Aaltonen, D Martuzevicius, U Haverinen-Shaughnessy | Assessment of indoor environmental quality in existing multi-family buildings in North-East Europe | Environ Int | 2015 | 79 | NA | 74-84 |
| Prasauskas T, D Martuzevicius, E Krugly, D Ciuzas, I Stasiulaitiene, R Sidaraviciute, V Kauneliene, L Seduikyte, A Jurelionis, U Haverinen-Shaughnessy | Spatial and temporal variations of particulate matter concentrations in multifamily apartment buildings | Building and Environment | 2014 | 76 | NA | 43755 |

| Malaysia | | | | | | |
|---|---|---|---|---|---|---|
| Nazariah, S.S.N., Juliana, J. and Abdah, M.A. | Interleukin-6 via sputum induction as biomarker of inflammation for indoor particulate matter among primary school children in Klang Valley, Malaysia | Global journal of health science | 2013 | 5 | 4 | 93 |

| Mexico |
|---|

| | | | | | | |
|---|---|---|---|---|---|---|
| Cortez-Lugo M, H Moreno-Macias, F Holguin-Molina, JC Chow, JG Watson, V Gutierrez-Avedoy, F Mandujano, M Hernandez-Avila, I Romieu | Relationship between indoor, outdoor, and personal fine particle concentrations for individuals with COPD and predictors of indoor-outdoor ratio in Mexico city | Journal of Exposure Science and Environmental Epidemiology | 2008 | 18 | 1 | 109-115 |
| Cortez-Lugo M, S Rodriguez-Dozal, I Rosas-Perez, U Alamo-Hernandez, H Riojas-Rodriguez | Modeling and estimating manganese concentrations in rural households in the mining district of Molango, Mexico | Environmental Monitoring and Assessment | 2015 | 187 | 12 | NA |
| Holguin F, MM Tellez-Rojo, M Hernandez, M Cortez, JC Chow, JG Watsow, D Mannino, I Romieu | Air pollution and heart rate variability among the elderly in Mexico City | Epidemiology | 2003 | 14 | 5 | 521-527 |
| Vallejo M, C Lerma, O Infante, AG Hermosillo, H Riojas-Rodriguez, M Cardenas | Personal exposure to particulate matter less than 2.5 mu m in Mexico City: a pilot study | Journal of Exposure Analysis and Environmental Epidemiology | 2004 | 14 | 4 | 323-329 |

| **Mongolia** | | | | | | |
|---|---|---|---|---|---|---|
| Barn P, E Gombojav, C Ochir, B Laagan, B Beejin, G Naidan, B Boldbaatar, J Galsuren, T Byambaa, C Janes, PA Janssen, BP Lanphear, T Takaro, SA Venners, GM Webster, W Yuchi, CD Palmer, PJ Parsons, YM Roh, RW Allen | The effect of portable HEPA filter air cleaners on indoor PM2.5 concentrations and second hand tobacco smoke exposure among pregnant women in Ulaanbaatar, Mongolia: The UGAAR randomized controlled trial | Science of the Total Environment | 2018 | 615 | NA | 1379-1389 |

| **Nepal** |
|---|

| | | | | | | |
|---|---|---|---|---|---|---|
| Shakya KM, RE Peltier, H Shrestha, RM Byanju | Measurements of TSP, PM10, PM2.5, BC, and PM chemical composition from an urban residential location in Nepal | Atmospheric Pollution Research | 2017 | 8 | 6 | 1123-1131 |

| **Netherlands** | | | | | | |
|---|---|---|---|---|---|---|
| de Hartog JJ, JG Ayres, A Karakatsani, A Analitis, H Brink, K Hameri, RM Harrison, K Katsouyanni, A Kotronarou, I Kavouras, C Meddings, J Pekkanen, G Hoek | Lung function and indicators of exposure to indoor and outdoor particulate matter among asthma and COPD patients | Occupational and Environmental Medicine | 2010 | 67 | 1 | 43506 |
| de Kluizenaar Y, E Kuijpers, I Eekhout, M Voogt, RCH Vermeulen, G Hoek, RP Sterkenburg, FH Pierik, JH Duyzer, EW Meijer, A Pronk | Personal exposure to UFP in different micro-environments and time of day | Building and Environment | 2017 | 122 | NA | 237-246 |
| Fischer PH, G Hoek, H van Reeuwijk, DJ Briggs, E Lebret, JH van Wijnen, S Kingham, PE Elliott | Traffic-related differences in outdoor and indoor concentrations of particles and volatile organic compounds in Amsterdam | Atmospheric Environment | 2000 | 34 | 22 | 3713-3722 |
| Hoek G, G Kos, RM Harrison, J de Hartog, K Meliefste, H ten Brink, K Katsouyanni, A Karakatsani, M Lianou, A Kotronarou, I Kavouras, J Pekkanen, M Vallius, M Kulmala, A Puustinen, S Thomas, C Meddings, J Ayres, | Indoor-outdoor relationships of particle number and mass in four European cities | Atmospheric Environment | 2008 | 42 | 1 | 156-169 |

| | | | | | | |
|---|---|---|---|---|---|---|
| J van Wijnen, K Hameri | | | | | | |
| Janssen NAH, T Lanki, G Hoek, M Vallius, JJ de Hartog, R Van Grieken, J Pekkanen, B Brunekreef | Associations between ambient, personal, and indoor exposure to fine particulate matter constituents in Dutch and Finnish panels of cardiovascular patients | Occupational and Environmental Medicine | 2005 | 62 | 12 | 868-877 |
| Janssen, N. A., de Hartog, J. J., Hoek, G., Brunekreef, B., Lanki, T., Timonen, K. L., & Pekkanen, J | Personal Exposure to Fine Particulate Matter in Elderly Subjects: Relation between Personal, Indoor, and Outdoor Concentrations | Journal of the Air & Waste Management Association | 2000 | 50 | 7 | 1133-1143 |
| Montagne, D., Hoek, G., Nieuwenhuijsen, M., Lanki, T., Siponen, T., Portella, M., Meliefste, K. and Brunekreef, B | Temporal associations of ambient PM2. 5 elemental concentrations with indoor and personal concentrations | Atmospheric Environment | 2014 | 86 | 0 | 203-211 |
| Mueller W, Steinle S, Parkka J, Parmes E, Liedes H, Kuijpers E, et al. | Urban greenspace and the indoor environment: Pathways to health via indoor particulate matter, noise, and road noise annoyance. | Environmental Research | 2020 | 180 | NA | 1E+05 |
| Yli-Tuomi T, T Lanki, G Hoek, B Brunekreef, J Pekkanen | Determination of the sources of indoor PM(2.5) in Amsterdam and Helsinki | Environmental Science & Technology | 2008 | 42 | 12 | 4440-4446 |

#### Nigeria

| | | | | | | |
|---|---|---|---|---|---|---|
| Ibhafidon LI, DO Obaseki, GE Erhabor, AA Akor, I Irabor, I Obioh | Respiratory symptoms, lung function and particulate matter pollution in residential indoor environment in Ile-Ife, Nigeria | Niger Med J | 2014 | 55 | 1 | 48-53 |

| **North Macedonia** | | | | | | |
|---|---|---|---|---|---|---|
| Vilcekova S, IZ Apostoloski, L Meciarova, EK Burdova, J Kiselak | Investigation of Indoor Air Quality in Houses of Macedonia | Int J Environ Res Public Health | 2017 | 14 | 1 | NA |

| **Norway** | | | | | | |
|---|---|---|---|---|---|---|
| Lazaridis M, V Aleksandropoulou, JE Hanssen, C Dye, K Eleftheriadis, E Katsivela | Inorganic and carbonaceous components in indoor/outdoor particulate matter in two residential houses in Oslo, Norway | Journal of the Air & Waste Management Association | 2008 | 58 | 3 | 346-356 |
| Oie L, P Magnus, BV Johansen | Suspended particulate matter in Norwegian dwellings in relation to fleece and shelf factors, domestic smoking, air exchange rate, and presence of hot wire convection heaters | Environment International | 1997 | 23 | 4 | 465-473 |
| Ormstad H, PI Gaarder, BV Johansen | Quantification and characterisation of suspended particulate matter in indoor air | Science of the Total Environment | 1997 | 193 | 3 | 185-196 |
| Rakkestad KE, CJ Dye, KE Yttri, JA Holme, JK Hongslo, PE Schwarze, R Becher | Phthalate levels in Norwegian indoor air related to particle size fraction | Journal of Environmental Monitoring | 2007 | 9 | 12 | 1419-1425 |
| Wyss AB, AC Jones, AK Bolling, GE Kissling, R Chartier, HJ Dahlman, CE Rodes, J Archer, J Thornburg, PE Schwarze, SJ London | Particulate Matter 2.5 Exposure and Self-Reported Use of Wood Stoves and Other Indoor Combustion Sources in Urban Nonsmoking Homes in Norway | Plos One | 2016 | 11 | 11 | NA |

| | | | | | | |
|---|---|---|---|---|---|---|
| **Oman** | | | | | | |
| Abdul-Wahab SA | Indoor and outdoor relationships of atmospheric particulates in Oman | Indoor and Built Environment | 2006 | 15 | 3 | 247-255 |
| **Pakistan** | | | | | | |
| Abbas M, S Razzaq, R Shahzadi, A Rahman, F Manzoor, Z Ali | DETERMINATION OF INDOOR AND OUTDOOR AIR QUALITY UNDER DIFFERENT VENTILATION CONDITIONS IN A RESIDENTIAL AREA OF LAHORE, PAKISTAN | Journal of Animal and Plant Sciences | 2015 | 25 | 3 | 672-676 |
| Ali Z, F Naz, ZA Nasir, S Sidra, I Colbeck | PARTICULATE POLLUTION IN URBAN RESIDENTIAL BUILT ENVIRONMENTS DURING WINTER AND SUMMER SEASONS IN LAHORE, PAKISTAN | Journal of Animal and Plant Sciences | 2015 | 25 | 3 | 660-665 |
| Colbeck I, Sidra S, Ali Z, Ahmed S, Nasir ZA. | Spatial and temporal variations in indoor air quality in Lahore, Pakistan. | International Journal of Environmental Science and Technology | 2019 | 16 | 6 | 2565-72 |
| Nasir, Z. A., Colbeck, I., Ali, Z., & Ahmad, S | Indoor particulate matter in developing countries: a case study in Pakistan and potential intervention strategies | Environmental Research Letters | 2013 | 8 | 2 | 24002 |
| **Peru** | | | | | | |
| Underhill LJ, S Bose, DL Williams, KM Romero, G Malpartida, PN Breysse, EM Klasen, JM Combe, W Checkley, NN Hansel | Association of Roadway Proximity with Indoor Air Pollution in a Peri-Urban Community in Lima, Peru | International Journal of Environmental Research and Public Health | 2015 | 12 | 10 | 13466-13481 |
| **Poland** | | | | | | |

| | | | | | | |
|---|---|---|---|---|---|---|
| Bentayeb M, D Norback, M Bednarek, A Bernard, GH Cai, S Cerrai, KK Eleftheriou, C Gratziou, GJ Holst, F Lavaud, J Nasilowski, P Sestini, G Sarno, T Sigsgaard, G Wieslander, J Zielinski, G Viegi, I Annesi-Maesano, G Study | Indoor air quality, ventilation and respiratory health in elderly residents Living in nursing homes in Europe | European Respiratory Journal | 2015 | 45 | 5 | 1228-1238 |
| Scibor M, Balcerzak B, Galbarczyk A, Targosz N, Jasienska G. | Are we safe inside? Indoor air quality in relation to outdoor concentration of PM10 and PM2.5 and to characteristics of homes. | Sustainable Cities and Society | 2019 | 48 | NA | 1E+05 |

| **Portugal** | | | | | | |
|---|---|---|---|---|---|---|
| Almeida-Silva M, SM Almeida, JF Gomes, PC Albuquerque, HT Wolterbeek | Determination of airborne nanoparticles in elderly care centers | J Toxicol Environ Health A | 2014 | 77 | NA | 867-78 |
| Almeida-Silva M, Pilou M, Housiadas C, Almeida SM. | Internal dose of particles in the elderly—modeling based on aerosol measurements. | Environmental Science and Pollution Research | 2018 | 25 | 24 | 23645-56 |
| Almeida-Silva M, T Faria, D Saraga, T Maggos, HT Wolterbeek, SM Almeida | Source apportionment of indoor PM10 in Elderly Care Centre | Environmental Science and Pollution Research | 2016 | 23 | 8 | 7814-7827 |
| Almeida-Silva, M., Wolterbeek, H.T. and Almeida, S.M | Elderly exposure to indoor air pollutants | Atmospheric Environment | 2014 | 85 | NA | 54-63 |

| | | | | | | |
|---|---|---|---|---|---|---|
| Belo J, Carreiro-Martins P, Papoila AL, Palmeiro T, Caires I, Alves M, et al. | The impact of indoor air quality on respiratory health of older people living in nursing homes: spirometric and exhaled breath condensate assessments. | Journal of Environmental Science and Health Part a-Toxic/Hazardous Substances & Environmental Engineering | 2019 | 54 | 12 | 1153-8 |
| Castro D, K Slezakova, C Delerue-Matos, MG Alvim-Ferraz, S Morais, MC Pereira | CONTRIBUTION OF TRAFFIC AND TOBACCO SMOKE IN THE DISTRIBUTION OF POLYCYCLIC AROMATIC HYDROCARBONS ON OUTDOOR AND INDOOR PM2.5 | Global Nest Journal | 2010 | 12 | 1 | 43535 |
| Cerqueira M, D Marques, A Caseiro, C Pio | Experimental evidence for a significant contribution of cellulose to indoor aerosol mass concentration | Atmospheric Environment | 2010 | 44 | 6 | 867-871 |
| Custódio, D., Pinho, I., Cerqueira, M., Nunes, T., & Pio, C. | Indoor and outdoor suspended particulate matter and associated carbonaceous species at residential homes in northwestern Portugal | Science of the total environment | 2014 | 473 | NA | 72-76 |
| Gomes JFP, JCM Bordado, PCS Albuquerque | Monitoring exposure to airborne ultrafine particles in Lisbon, Portugal | Inhalation Toxicology | 2012 | 24 | 7 | 425-433 |
| Madureira J, I Paciencia, J Cavaleiro-Rufo, E de Oliveira Fernandes | Indoor pollutant exposure among children with and without asthma in Porto, Portugal, during the cold season | Environ Sci Pollut Res Int | 2016 | 23 | 20 | 20539-20552 |
| Mendes A, AL Papoila, P Carreiro-Martins, S Bonassi, I Caires, T Palmeiro, L Aguiar, C Pereira, P Neves, D Mendes, MAS | The impact of indoor air quality and contaminants on respiratory health of older people living in long-term care residences in Porto | Age and Ageing | 2016 | 45 | 1 | 136-142 |

| | | | | | | |
|---|---|---|---|---|---|---|
| Botelho, N Neuparth, JP Teixeira | | | | | | |
| Miguel AF, AH Reis, M Melgao | Urban indoor-outdoor aerosol measurements in Portugal and the global warming scenario | International Journal of Global Warming | 2009 | 1 | NA | 356-367 |
| Slezakova K, C Texeira, S Morais, MD Pereira | CHILDREN'S INDOOR EXPOSURES TO (ULTRA) FINE PARTICLES IN AN URBAN AREA: COMPARISON BETWEEN SCHOOL AND HOME ENVIRONMENTS | Journal of Toxicology and Environmental Health-Part a-Current Issues | 2015 | 78 | NA | 886-896 |
| Slezakova K, D Castro, C Delerue-Matos, S Morais, MD Pereira | Levels and risks of particulate-bound PAHs in indoor air influenced by tobacco smoke: a field measurement | Environmental Science and Pollution Research | 2014 | 21 | 6 | 4492-4501 |

| **Saudi Arabia** | | | | | | |
|---|---|---|---|---|---|---|
| Ali N. | Polycyclic aromatic hydrocarbons (PAHs) in indoor air and dust samples of different Saudi microenvironments; health and carcinogenic risk assessment for the general population. | NA | 2019 | 696 | NA | 1E+05 |

| **Singapore** | | | | | | |
|---|---|---|---|---|---|---|
| Balasubramanian R, P Nainar, A Rajasekar | Airborne bacteria, fungi, and endotoxin levels in residential microenvironments: a case study | Aerobiologia | 2012 | 28 | 3 | 375-390 |

| | | | | | | |
|---|---|---|---|---|---|---|
| Balasubramanian R, SS Lee | Characteristics of indoor aerosols in residential homes in urban locations: A case study in Singapore | Journal of the Air & Waste Management Association | 2007 | 57 | 8 | 981-990 |
| Lai ACK, YW Ho | Spatial concentration variation of cooking-emitted particles in a residential kitchen | Building and Environment | 2008 | 43 | 5 | 871-876 |
| Sharm R, R Balasubramanian | Size-fractionated Particulate Matter in Indoor and Outdoor Environments during the 2015 Haze in Singapore: Potential Human Health Risk Assessment | Aerosol and Air Quality Research | 2018 | 18 | 4 | 904-917 |
| Zhou J, AL Chen, QL Cao, B Yang, VWC Chang, WW Nazaroff | Particle exposure during the 2013 haze in Singapore: Importance of the built environment | Building and Environment | 2015 | 93 | NA | 14-23 |
| **Slovakia** | | | | | | |
| Brauer M, F Hruba, E Mihalikova, E Fabianova, P Miskovic, A Plzikova, M Lendacka, J Vandenberg, A Cullen | Personal exposure to particles in Banska Bystrica, Slovakia | Journal of Exposure Analysis and Environmental Epidemiology | 2000 | 10 | 5 | 478-487 |
| Meciarova L, S Vilcekova, EK Burdova, J Kiselak | Factors Effecting the Total Volatile Organic Compound (TVOC) Concentrations in Slovak Households | International Journal of Environmental Research and Public Health | 2017 | 14 | 12 | NA |
| **South Africa** | | | | | | |
| Adesina JA, Piketh SJ, Qhekwana M, Burger R, Language B, Mkhatshwa G. | Contrasting indoor and ambient particulate matter concentrations and thermal comfort in coal and non-coal burning households at South Africa Highveld. | Science of the total environment | 2020 | 699 | 10 | 1E+05 |

| | | | | | | |
|---|---|---|---|---|---|---|
| Gumede PR, MJ Savage | Respiratory health effects associated with indoor particulate matter (PM2.5) in children residing near a landfill site in Durban, South Africa | Air Quality Atmosphere and Health | 2017 | 10 | 7 | 853-860 |
| Vanker A, Nduru PM, Barnett W, Dube FS, Sly PD, Gie RP, et al. | Indoor air pollution and tobacco smoke exposure: impact on nasopharyngeal bacterial carriage in mothers and infants in an African birth cohort study. | ERJ Open Research | 2019 | 5 | 1 | 00052-2018 |
| Vanker A, W Barnett, PM Nduru, RP Gie, PD Sly, HJ Zar | Home environment and indoor air pollution exposure in an African birth cohort study | Science of the Total Environment | 2015 | 536 | NA | 362-367 |

| **South Korea** | | | | | | |
|---|---|---|---|---|---|---|
| Baek SO, YS Kim, R Perry | Indoor air quality in homes, offices and restaurants in Korean urban areas - Indoor/outdoor relationships | Atmospheric Environment | 1997 | 31 | 4 | 529-544 |
| Byun H, H Bae, D Kim, H Shin, C Yoon | Effects of socioeconomic factors and human activities on children's PM10 exposure in inner-city households in Korea | International Archives of Occupational and Environmental Health | 2010 | 83 | 8 | 867-878 |
| Choi DH, DH Kang | Infiltration of Ambient PM2.5 through Building Envelope in Apartment Housing Units in Korea | Aerosol and Air Quality Research | 2017 | 17 | 2 | 598-607 |
| Hwang Y, K Lee | Contribution of microenvironments to personal exposures to PM10 and PM2.5 in summer and winter | Atmospheric Environment | 2018 | 175 | NA | 192-198 |

| | | | | | | |
|---|---|---|---|---|---|---|
| Jo WK, JY Lee | Indoor and outdoor levels of respirable particulates (PM10) and Carbon Monoxide (CO) in high-rise apartment buildings | Atmospheric Environment | 2006 | 40 | 32 | 6067-6076 |
| Kang K, Kim H, Kim DD, Lee YG, Kim T. | Characteristics of cooking-generated PM10 and PM2.5 in residential buildings with different cooking and ventilation types. | Science of The Total Environment | 2019 | 668 | NA | 56-66 |
| Lee CH, BK Lee, IB Oh, JH Lee, CS Sim, Y Kim | Indoor Air Quality in Elementary School Children's Homes in Ulsan: Comparison between Groups with and without Allergic Rhinitis | Journal of Korean Society for Atmospheric Environment | 2012 | 28 | 4 | 365-373 |
| Lee JH, HS Lee, MR Park, SW Lee, EH Kim, JB Cho, J Kim, Y Han, K Jung, HK Cheong, SI Lee, K Ahn | Relationship Between Indoor Air Pollutant Levels and Residential Environment in Children With Atopic Dermatitis | Allergy Asthma & Immunology Research | 2014 | 6 | 6 | 517-524 |
| Lee JY, C Kim, J Kim, SH Ryu, GN Bae | Exposure Assessments for Children in Homes and in Daycare Centers to NO2, PMs and Black Carbon | Asian Journal of Atmospheric Environment | 2018 | 12 | 3 | 204-214 |
| Lim S, J Kim, T Kim, K Lee, W Yang, S Jun, S Yu | Personal exposures to PM2.5 and their relationships with microenvironmental concentrations | Atmospheric Environment | 2012 | 47 | NA | 407-412 |
| Oh HJ, Jeong NN, Sohn JR, Kim J. | Personal exposure to indoor aerosols as actual concern: Perceived indoor and outdoor air quality, and health performances. | Building and Environment | 2019 | 165 | NA | 1E+05 |
| Park JS, NY Jee, JW Jeong | Effects of types of ventilation system on indoor particle concentrations in residential buildings | Indoor Air | 2014 | 24 | 6 | 629-638 |

| | | | | | | |
|---|---|---|---|---|---|---|
| Seo SC, IS Kang, SG Lim, JT Choung, Y Yoo | Indoor air pollutants and atopic dermatitis in socioeconomically disadvantaged children | Allergy Asthma & Respiratory Disease | 2015 | 3 | 3 | 206-212 |

| **Spain** | | | | | | |
|---|---|---|---|---|---|---|
| Dadvand P, A de Nazelle, M Triguero-Mas, A Schembari, M Cirach, E Amoly, F Figueras, X Basagana, B Ostro, M Nieuwenhuijsen | Surrounding Greenness and Exposure to Air Pollution During Pregnancy: An Analysis of Personal Monitoring Data | Environmental Health Perspectives | 2012 | 120 | 9 | 1286-1290 |
| Fernandez E, M Ballbe, X Sureda, M Fu, E Salto, JM Martinez-Sanchez | Particulate Matter from Electronic Cigarettes and Conventional Cigarettes: a Systematic Review and Observational Study | Curr Environ Health Rep | 2015 | 2 | 4 | 423-9 |
| Montagne, D., Hoek, G., Nieuwenhuijsen, M., Lanki, T., Siponen, T., Portella, M., Meliefste, K. and Brunekreef, B | Temporal associations of ambient PM2. 5 elemental concentrations with indoor and personal concentrations | Atmospheric Environment | 2014 | 86 | 0 | 203-211 |
| Panella P, M Casas, D Donaire-Gonzalez, R Garcia-Esteban, O Robinson, A Valentin, J Gulliver, I Momas, M Nieuwenhuijsen, M Vrijheid, J Sunyer | Ultrafine particles and black carbon personal exposures in asthmatic and non-asthmatic children at school age | Indoor Air | 2017 | 27 | 5 | 891-899 |
| Schembari A, M Triguero-Mas, A de Nazelle, P Dadvand, M Vrijheid, M Cirach, D Martinez, F Figueras, X Querol, X Basagana, M Eeftens, K Meliefste, MJ Nieuwenhuijsen | Personal, indoor and outdoor air pollution levels among pregnant women | Atmospheric Environment | 2013 | 64 | NA | 287-295 |

| Sri Lanka | | | | | | |
|---|---|---|---|---|---|---|
| Ranasinghe RSA, AGT Sugathapala, SC Lee, WT Hung, KF Ho, CS Chan, Y Huang, Y Cheng | Exploratory study of the indoor and outdoor relationships and chemical compositions of particulate matter in urban households in Colombo | Indoor and Built Environment | 2015 | 24 | 5 | 597-606 |

| Sweden | | | | | | |
|---|---|---|---|---|---|---|
| Bentayeb M, D Norback, M Bednarek, A Bernard, GH Cai, S Cerrai, KK Eleftheriou, C Gratziou, GJ Holst, F Lavaud, J Nasilowski, P Sestini, G Sarno, T Sigsgaard, G Wieslander, J Zielinski, G Viegi, I Annesi-Maesano, G Study | Indoor air quality, ventilation and respiratory health in elderly residents Living in nursing homes in Europe | European Respiratory Journal | 2015 | 45 | 5 | 1228-1238 |
| Isaxon C, A Gudmundsson, EZ Nordin, L Lonnblad, A Dahl, G Wieslander, M Bohgard, A Wierzbicka | Contribution of indoor-generated particles to residential exposure | Atmospheric Environment | 2015 | 106 | NA | 458-466 |
| Johannesson S, P Gustafson, P Molnar, L Barregard, G Sallsten | Exposure to fine particles (PM2.5 and PM1) and black smoke in the general population: personal, indoor, and outdoor levels | Journal of Exposure Science and Environmental Epidemiology | 2007 | 17 | 7 | 613-624 |
| Matson U | Indoor and outdoor concentrations of ultrafine particles in some Scandinavian rural and urban areas | Science of the Total Environment | 2005 | 343 | NA | 169-176 |

| | | | | | | |
|---|---|---|---|---|---|---|
| Molnar P, P Gustafson, S Johannesson, J Boman, L Barregard, G Sallsten | Domestic wood burning and PM2.5 trace elements: Personal exposures, indoor and outdoor levels | Atmospheric Environment | 2005 | 39 | 14 | 2643-2653 |
| Wichmann J, T Lind, MAM Nilsson, T Bellander | PM2.5, soot and NO2 indoor-outdoor relationships at homes, pre-schools and schools in Stockholm, Sweden | Atmospheric Environment | 2010 | 44 | 36 | 4536-4544 |
| Wierzbicka A, M Bohgard, JH Pagels, A Dahl, J Londahl, T Hussein, E Swietlicki, A Gudmundsson | Quantification of differences between occupancy and total monitoring periods for better assessment of exposure to particles in indoor environments | Atmospheric Environment | 2015 | 106 | NA | 419-428 |

| **Switzerland** | | | | | | |
|---|---|---|---|---|---|---|
| Hanninen OO, E Lebret, V Ilacqua, K Katsouyanni, F Kunzli, RJ Sram, M Jantunen | Infiltration of ambient PM2.5 and levels of indoor generated non-ETS PM2.5 in residences of four European cities | Atmospheric Environment | 2004 | 38 | 37 | 6411-6423 |
| Kousa A, L Oglesby, K Koistinen, N Kunzli, M Jantunen | Exposure chain of urban air PM2.5 - associations between ambient fixed site, residential outdoor, indoor, workplace and personal exposures in four European cities in the EXPOLIS-study | Atmospheric Environment | 2002 | 36 | 18 | 3031-3039 |
| Meier R, M Eeftens, HC Phuleria, A Ineichen, E Corradi, M Davey, M Fierz, RE Ducret-Stich, I Aguilera, C Schindler, T Rochat, N Probst-Hensch, MY Tsai, N Kunzli | Differences in indoor versus outdoor concentrations of ultrafine particles, PM2.5, PMabsorbance and NO2 in Swiss homes | J Expo Sci Environ Epidemiol | 2015 | 25 | 5 | 499-505 |

| | | | | | | |
|---|---|---|---|---|---|---|
| Monn C, A Fuchs, D Hogger, M Junker, D Kogelschatz, N Roth, HU Wanner | Particulate matter less than 10 mu m (PM10) and fine particles less than 2.5 mu m (PM2.5): relationships between indoor, outdoor and personal concentrations | Science of the Total Environment | 1997 | 208 | NA | 15-21 |
| Monn C, G Schaeppi | CONCENTRATIONS OF TOTAL SUSPENDED PARTICULATES, FINE PARTICLES AND THEIR ANIONIC COMPOUNDS IN AMBIENT AIR AND INDOOR AIR | Environmental Technology | 1993 | 14 | 9 | 869-875 |
| Phillips K, DA Howard, MC Bentley, G Alvan | Assessment of environmental tobacco smoke and respirable suspended particle exposures for nonsmokers in Basel by personal monitoring | Atmospheric Environment | 1999 | 33 | 12 | 1889-1904 |

| **Taiwan** | | | | | | |
|---|---|---|---|---|---|---|
| Chang LT, GB Hong, SP Weng, HC Chuang, TY Chang, CW Liu, WY Chuang, KJ Chuang | Indoor ozone levels, houseplants and peak expiratory flow rates among healthy adults in Taipei, Taiwan | Environ Int | 2018 | NA | NA | NA |
| Chi, M.C., Guo, S.E., Hwang, S.L., Chou, C.T., Lin, C.M. and Lin, Y.C | Exposure to Indoor Particulate MatterWorsens the Symptoms and Acute Exacerbations in Chronic Obstructive Pulmonary Disease Patients of Southwestern Taiwan: A Pilot Study | International journal of environmental research and public health | 2017 | 14 | 1 | 4 |

| | | | | | | |
|---|---|---|---|---|---|---|
| Chuang HC, KF Ho, LY Lin, TY Chang, GB Hong, CM Ma, IJ Liu, KJ Chuang | Long-term indoor air conditioner filtration and cardiovascular health: A randomized crossover intervention study | Environment International | 2017 | 106 | NA | 91-96 |
| Hsu NY, CC Lee, JY Wang, YC Li, HW Chang, CY Chen, CG Bornehag, PC Wu, J Sundell, HJ Su | Predicted risk of childhood allergy, asthma, and reported symptoms using measured phthalate exposure in dust and urine | Indoor Air | 2012 | 22 | 3 | 186-199 |
| Huang YL, HW Chen, BC Han, CW Liu, HC Chuang, LY Lin, KJ Chuang | Personal Exposure to Household Particulate Matter, Household Activities and Heart Rate Variability among Housewives | Plos One | 2014 | 9 | 3 | NA |
| Li CS | RELATIONSHIPS OF INDOOR OUTDOOR INHALABLE AND RESPIRABLE PARTICLES IN DOMESTIC ENVIRONMENTS | Science of the Total Environment | 1994 | 151 | 3 | 205-211 |
| Li CS | ELEMENTAL COMPOSITION OF RESIDENTIAL INDOOR PM10 IN THE URBAN ATMOSPHERE OF TAIPEI | Atmospheric Environment | 1994 | 28 | 19 | 3139-3144 |
| Li CS, CH Lin | Carbon profile of residential indoor PM1 and PM2.5 in the subtropical region | Atmospheric Environment | 2003 | 37 | 7 | 881-888 |
| Li CS, WH Lin, FT Jenq | SIZE DISTRIBUTIONS OF SUBMICROMETER AEROSOLS FROM COOKING | Environment International | 1993 | 19 | 2 | 147-154 |
| Lin LY, CY Lin, YC Lin, KJ Chuang | The effects of indoor particles on blood pressure and heart rate among young adults in Taipei, Taiwan | Indoor Air | 2009 | 19 | 6 | 482-488 |

| | | | | | | |
|---|---|---|---|---|---|---|
| Lin LY, HC Chuang, IJ Liu, HW Chen, KJ Chuang | Reducing indoor air pollution by air conditioning is associated with improvements in cardiovascular health among the general population | Science of the Total Environment | 2013 | 463 | NA | 176-181 |
| Lin LY, HW Chen, TL Su, GB Hong, LC Huang, KJ Chuang | The effects of indoor particle exposure on blood pressure and heart rate among young adults: An air filtration-based intervention study | Atmospheric Environment | 2011 | 45 | 31 | 5540-5544 |
| Lin LY, IJ Liu, HC Chuang, HY Lin, KJ Chuang | Size and composition effects of household particles on inflammation and endothelial dysfunction of human coronary artery endothelial cells | Atmospheric Environment | 2013 | 77 | NA | 490-495 |
| Lung, S.C.C., Mao, I.F. and Liu, L.J.S. | Residents' particle exposures in six different communities in Taiwan | Science of the total environment | 2007 | 377 | 1 | 81-92 |
| Yen YC, Yang CY, Mena KD, Cheng YT, Chen PS. | Cooking/Window Opening and Associated Increases of Indoor PM2.5 and NO2 Concentrations of Children's Houses in Kaohsiung, Taiwan. | Applied Sciences-Basel | 2019 | 9 | 20 | Epub |
| Yen YC, Yang CY, Mena KD, Cheng YT, Yuan CS, Chen PS. | Jumping on the bed and associated increases of PM10, PM2.5, PM1, airborne endotoxin, bacteria, and fungi concentrations. | Environ Pollut | 2019 | 245 | NA | 799-809 |
| Yu KP, KR Yang, YC Chen, JY Gong, YP Chen, HC Shih, SCC Lung | Indoor air pollution from gas cooking in five Taiwanese families | Building and Environment | 2015 | 93 | NA | 258-266 |

**Thailand**

| | | | | | | |
|---|---|---|---|---|---|---|
| Kammoolkon R, Taneepanichskul N, Siriwong W, Pitaknoppakul N, Lohsoonthorn V. | The relationship between household particulate matter and an increase of carotid intima-media thickness [cimt]: A one-year follow-up study. | Journal of the Medical Association of Thailand | 2018 | 101 | 11 | 1529-36 |
| Lappharat S, Taneepanichskul N, Reutrakul S, Chirakalwasan N. | Effects of bedroom environmental conditions on the severity of obstructive sleep apnea. | Journal of clinical sleep medicine: JCSM: Official Publication of the American Academy of Sleep Medicine | 2018 | 14 | 4 | 565-73 |
| Srithawirat T, MT Latif, FR Sulaiman | Indoor PM10 and its heavy metal composition at a roadside residential environment, Phitsanulok, Thailand | Atmosfera | 2016 | 29 | 4 | 311-322 |
| Tsai FC, KR Smith, N Vichit-Vadakan, BD Ostro, LG Chestnut, N Kungskulniti | Indoor/outdoor PM10 and PM2.5 in Bangkok, Thailand | Journal of Exposure Analysis and Environmental Epidemiology | 2000 | 10 | 1 | 15-26 |

| **Turkey** | | | | | | |
|---|---|---|---|---|---|---|
| Mentese S, AY Rad, M Arisoy, G Gullu | Multiple comparisons of organic, microbial, and fine particulate pollutants in typical indoor environments: Diurnal and seasonal variations | Journal of the Air & Waste Management Association | 2012 | 62 | 12 | 1380-1393 |
| Pekey B, ZB Bozkurt, H Pekey, G Dogan, A Zararsiz, N Efe, G Tuncel | Indoor/outdoor concentrations and elemental composition of PM10/PM2.5 in urban/industrial areas of Kocaeli City, Turkey | Indoor Air | 2010 | 20 | 2 | 112-125 |

| **UK** |
|---|

| | | | | | | |
|---|---|---|---|---|---|---|
| Aizlewood C, C Dimitroulopoulou | The HOPE project: The UK experience | Indoor and Built Environment | 2006 | 15 | 5 | 393-409 |
| BeruBe KA, KJ Sexton, TP Jones, T Moreno, S Anderson, RJ Richards | The spatial and temporal variations in PM10 mass from six UK homes | Science of the Total Environment | 2004 | 324 | NA | 41-53 |
| de Hartog JJ, JG Ayres, A Karakatsani, A Analitis, H Brink, K Hameri, RM Harrison, K Katsouyanni, A Kotronarou, I Kavouras, C Meddings, J Pekkanen, G Hoek | Lung function and indicators of exposure to indoor and outdoor particulate matter among asthma and COPD patients | Occupational and Environmental Medicine | 2010 | 67 | 1 | 43506 |
| Dobson R, Semple S. | How do you know those particles are from cigarettes?: An algorithm to help differentiate second-hand tobacco smoke from background sources of household fine particulate matter. | Environmental Research | 2018 | 166 | NA | 344-7 |
| Galea KS, JF Hurley, H Cowie, AL Shafrir, AS Jimenez, S Semple, JG Ayres, M Coggins | Using PM2.5 concentrations to estimate the health burden from solid fuel combustion, with application to Irish and Scottish homes | Environmental Health | 2013 | 12 | NA | NA |
| Halsall CJ, BA Maher, VV Karloukovski, P Shah, SJ Watkins | A novel approach to investigating indoor/outdoor pollution links: Combined magnetic and PAH measurements | Atmospheric Environment | 2008 | 42 | 39 | 8902-8909 |

| | | | | | | |
|---|---|---|---|---|---|---|
| Hoek G, G Kos, RM Harrison, J de Hartog, K Meliefste, H ten Brink, K Katsouyanni, A Karakatsani, M Lianou, A Kotronarou, I Kavouras, J Pekkanen, M Vallius, M Kulmala, A Puustinen, S Thomas, C Meddings, J Ayres, J van Wijnen, K Hameri | Indoor-outdoor relationships of particle number and mass in four European cities | Atmospheric Environment | 2008 | 42 | 1 | 156-169 |
| Jones NC, CA Thornton, D Mark, RM Harrison | Indoor/outdoor relationships of particulate matter in domestic homes with roadside, urban and rural locations | Atmospheric Environment | 2000 | 34 | 16 | 2603-2612 |
| Kingham S, D Briggs, P Elliott, P Fischer, E Lebret | Spatial variations in the concentrations of traffic-related pollutants in indoor and outdoor air in Huddersfield, England | Atmospheric Environment | 2000 | 34 | 6 | 905-916 |
| Lai HK, M Kendall, H Ferrier, I Lindup, S Alm, O Hanninen, M Jantunen, P Mathys, R Colvile, MR Ashmore, P Cullinan, MJ Nieuwenhuijsen | Personal exposures and microenvironment concentrations of PM2.5, VOC, NO2 and CO in Oxford, UK | Atmospheric Environment | 2004 | 38 | 37 | 6399-6410 |
| Mills LM, SE Semple, IS Wilson, L MacCalman, A Amos, D Ritchie, R O'Donnell, A Shaw, SW Turner | Factors Influencing Exposure to Secondhand Smoke in Preschool Children Living With Smoking Mothers | Nicotine & Tobacco Research | 2012 | 14 | 12 | 1435-1444 |
| Mohammadyan M, M Ashmore | Personal exposure and indoor PM2.5 concentrations in an urban population | Indoor and Built Environment | 2005 | 14 | NA | 313-320 |

| | | | | | | |
|---|---|---|---|---|---|---|
| Moreno-Rangel A, T Sharpe, F Musau, G McGill | Field evaluation of a low-cost indoor air quality monitor to quantify exposure to pollutants in residential environments | Journal of Sensors and Sensor Systems | 2018 | 7 | 1 | 373-388 |
| Mueller W, Steinle S, Parkka J, Parmes E, Liedes H, Kuijpers E, et al. | Urban greenspace and the indoor environment: Pathways to health via indoor particulate matter, noise, and road noise annoyance. | Environmental Research | 2020 | 180 | NA | 1E+05 |
| Nasir ZA, I Colbeck | Particulate pollution in different housing types in a UK suburban location | Science of the Total Environment | 2013 | 445 | NA | 165-176 |
| O'Connell S, HKC Au-Yeung, CJ Gregory, IP Matthews | Outdoor and indoor respirable air particulate concentrations in differing urban traffic microenvironments | Journal of Toxicology and Environmental Health-Part a-Current Issues | 2008 | 71 | 16 | 1069-1072 |
| Osman LM, JG Douglas, C Garden, K Reglitz, J Lyon, S Gordon, JG Ayres | Indoor air quality in homes of patients with chronic obstructive pulmonary disease | American Journal of Respiratory and Critical Care Medicine | 2007 | 176 | 5 | 465-472 |
| Semple, S., Garden, C., Coggins, M., Galea, K.S., Whelan, P., Cowie, H., Sánchez-Jiménez, A., Thorne, P.S., Hurley, J.F. and Ayres, J.G | Contribution of solid fuel, gas combustion, or tobacco smoke to indoor air pollutant concentrations in Irish and Scottish homes | Indoor air | 2012 | 22 | 3 | 212-223 |
| Stewart L, IL Gee, AFR Watson, G Fletcher, R Niven | Indoor air quality and childhood asthma: Method development and preliminary results | Indoor and Built Environment | 2001 | 10 | NA | 271-275 |
| Tan CCL, KN Finney, Q Chen, NV Russell, VN Sharifi, J Swithenbank | Experimental Investigation of Indoor Air Pollutants in Residential Buildings | Indoor and Built Environment | 2013 | 22 | 3 | 471-489 |

| | | | | | | |
|---|---|---|---|---|---|---|
| Wigzell E, M Kendall, MJ Nieuwenhuijsen | The spatial and temporal variation of particulate matter within the home | Journal of Exposure Analysis and Environmental Epidemiology | 2000 | 10 | 3 | 307-314 |
| Wilson I, S Semple, LM Mills, D Ritchie, A Shaw, R O'Donnell, P Bonella, S Turner, A Amos | REFRESHreducing families' exposure to secondhand smoke in the home: a feasibility study | Tobacco Control | 2013 | 22 | 5 | NA |
| Woods KE, A Apsley, S Semple, SW Turner | Domestic airborne fine particulate matter exposure and asthma control among children receiving inhaled steroid treatment | Indoor and Built Environment | 2014 | 23 | 3 | 497-503 |

| **United Arab Emirates** | | | | | | |
|---|---|---|---|---|---|---|
| Weitzman M, AH Yusufali, F Bali, MJR Vilcassim, S Gandhi, R Peltier, A Nadas, S Sherman, L Lee, Z Hong, J Shearston, SH Park, T Gordon | Effects of hookah smoking on indoor air quality in homes | Tobacco Control | 2017 | 26 | 5 | 586-591 |
| Yeatts KB, M El-Sadig, D Leith, W Kalsbeek, F Al-Maskari, D Couper, WE Funk, T Zoubeidi, RL Chan, CB Trent, CA Davidson, MG Boundy, MM Kassab, MY Hasan, I Rusyn, JM Gibson, AF Olshan | Indoor Air Pollutants and Health in the United Arab Emirates | Environmental Health Perspectives | 2012 | 120 | 5 | 687-694 |

| **USA** | | | | | | |
|---|---|---|---|---|---|---|
| Abraham ME | Microanalysis of indoor aerosols and the impact of a Compact High-Efficiency Particulate air (HEPA) filter system | Indoor Air-International Journal of Indoor Air Quality and Climate | 1999 | 9 | 1 | 33-40 |

| | | | | | | |
|---|---|---|---|---|---|---|
| Abt E, HH Suh, G Allen, P Koutrakis | Characterization of indoor particle sources: A study conducted in the metropolitan Boston area | Environmental Health Perspectives | 2000 | 108 | 1 | 35-44 |
| Adgate JL, G Ramachandran, GC Pratt, LA Waller, K Sexton | Spatial and temporal variability in outdoor, indoor, and personal PM2.5 exposure | Atmospheric Environment | 2002 | 36 | 20 | 3255-3265 |
| Adgate JL, Ramachandran G, Pratt GC, Waller LA, Sexton K | Longitudinal variability in outdoor, indoor, and personal PM2.5 exposure in healthy non-smoking adults | Atmospheric Environment | 2003 | 37 | 7 | 993-1002 |
| Allen R, T Larson, L Sheppard, L Wallace, LJ Liu | Use of real-time light scattering data to estimate the contribution of infiltrated and indoor-generated particles to indoor air | Environ Sci Technol | 2003 | 37 | 16 | 3484-92 |
| Anuszewski J, TV Larson, JQ Koenig | Simultaneous indoor and outdoor particle light-scattering measurements at nine homes using a portable nephelometer | Journal of Exposure Analysis and Environmental Epidemiology | 1998 | 8 | 4 | 483-493 |
| Arhami M, A Polidori, RJ Delfino, T Tjoa, C Sioutas | Associations between Personal, Indoor, and Residential Outdoor Pollutant Concentrations: Implications for Exposure Assessment to Size-Fractionated Particulate Matter | Journal of the Air & Waste Management Association | 2009 | 59 | 4 | 392-404 |
| Arku RE, G Adamkiewicz, J Vallarino, JD Spengler, DE Levy | Seasonal variability in environmental tobacco smoke exposure in public housing developments | Indoor Air | 2015 | 25 | 1 | 13-20 |

| | | | | | | |
|---|---|---|---|---|---|---|
| Balmes JR, M Cisternas, PJ Quinlan, L Trupin, FW Lurmann, PP Katz, PD Blanc | Annual average ambient particulate matter exposure estimates, measured home particulate matter, and hair nicotine are associated with respiratory outcomes in adults with asthma | Environmental Research | 2014 | 129 | NA | 43475 |
| Batterman S, C Godwin, CR Jia | Long duration tests of room air filters in cigarette smokers' homes | Environmental Science & Technology | 2005 | 39 | 18 | 7260-7268 |
| Batterman S, L Du, G Mentz, B Mukherjee, E Parker, C Godwin, JY Chin, A O'Toole, T Robins, Z Rowe, T Lewis | Particulate matter concentrations in residences: an intervention study evaluating stand-alone filters and air conditioners | Indoor Air | 2012 | 22 | 3 | 235-252 |
| Baxter LK, JE Clougherty, CJ Paciorek, RJ Wright, JI Levy | Predicting residential indoor concentrations of nitrogen dioxide, fine particulate matter, and elemental carbon using questionnaire and geographic information system based data | Atmospheric Environment | 2007 | 41 | 31 | 6561-6571 |
| Beamer PI, AJ Sugeng, MD Kelly, N Lothrop, W Klimecki, ST Wilkinson, M Loh | Use of dust fall filters as passive samplers for metal concentrations in air for communities near contaminated mine tailings | Environmental Science-Processes & Impacts | 2014 | 16 | 6 | 1275-1281 |
| Belli AJ, S Bose, N Aggarwal, C DaSilva, S Thapa, L Grammer, LM Paulin, NN Hansel | Indoor particulate matter exposure is associated with increased black carbon content in airway macrophages of former smokers with COPD | Environmental Research | 2016 | 150 | NA | 398-402 |
| Berry D, G Mainelis, D Fennell | Effect of an ionic air cleaner on indoor/outdoor particle ratios in a residential environment | Aerosol Science and Technology | 2007 | 41 | 3 | 315-328 |

| | | | | | | |
|---|---|---|---|---|---|---|
| Bhangar S, NA Mullen, SV Hering, NM Kreisberg, WW Nazaroff | Ultrafine particle concentrations and exposures in seven residences in northern California | Indoor Air | 2011 | 21 | 2 | 132-144 |
| Breen MS, TC Long, BD Schultz, RW Williams, J Richmond-Bryant, M Breen, JE Langstaff, RB Devlin, A Schneider, JM Burke, SA Batterman, QY Meng | Air Pollution Exposure Model for Individuals (EMI) in Health Studies: Evaluation for Ambient PM2.5 in Central North Carolina | Environmental Science & Technology | 2015 | 49 | 24 | 14184-14194 |
| Breysse PN, TJ Buckley, D Williams, CM Beck, SJ Jo, B Merriman, S Kanchanaraksa, LJ Swartz, KA Callahan, AM Butz, CS Rand, GB Diette, JA Krishnan, AM Moseley, J Curtin-Brosnan, NB Durkin, PA Eggleston | Indoor exposures to air pollutants and allergens in the homes of asthmatic children in inner-city Baltimore | Environmental Research | 2005 | 98 | 2 | 167-176 |
| Brown KW, JA Sarnat, HH Suh, BA Coull, JD Spengler, P Koutrakis | Ambient site, home outdoor and home indoor particulate concentrations as proxies of personal exposures | Journal of Environmental Monitoring | 2008 | 10 | 9 | 1041-1051 |
| Brown, D.R., Alderman, N., Weinberger, B., Lewis, C., Bradley, J. and Curtis, L. | Outdoor wood furnaces create significant indoor particulate pollution in neighboring homes | Inhalation toxicology | 2014 | 26 | 10 | 628-635 |
| Brugge D, J Vallarino, L Ascolillo, ND Osgood, S Steinbach, J Spengler | Comparison of multiple environmental factors for asthmatic children in public housing | Indoor Air | 2003 | 13 | 1 | 18-27 |

| | | | | | | |
|---|---|---|---|---|---|---|
| Brugge D, MC Simon, N Hudda, M Zellmer, L Corlin, S Cleland, EY Lu, S Rivera, M Byrne, M Chung, JL Durant | Lessons from in-home air filtration intervention trials to reduce urban ultrafine particle number concentrations | Building and Environment | 2017 | 126 | NA | 266-275 |
| Butz AM, EC Matsui, P Breysse, J Curtin-Brosnan, P Eggleston, G Diette, D Williams, J Yuan, JT Bernert, C Rand | A Randomized Trial of Air Cleaners and a Health Coach to Improve Indoor Air Quality for Inner-City Children With Asthma and Secondhand Smoke Exposure | Archives of Pediatrics & Adolescent Medicine | 2011 | 165 | 8 | 741-748 |
| Butz AM, P Breysse, C Rand, J Curtin-Brosnan, P Eggleston, GB Diette, D Williams, JT Bernert, EC Matsui | Household Smoking Behavior: Effects on Indoor Air Quality and Health of Urban Children with Asthma | Maternal and Child Health Journal | 2011 | 15 | 4 | 460-468 |
| Chan WR, JM Logue, X Wu, NE Klepeis, WJ Fisk, F Noris, BC Singer | Quantifying fine particle emission events from time-resolved measurements: Method description and application to 18 California low-income apartments | Indoor Air | 2018 | 28 | 1 | 89-101 |
| Chen Q, LM Hildemann | Size-Resolved Concentrations of Particulate Matter and Bioaerosols Inside versus Outside of Homes | Aerosol Science and Technology | 2009 | 43 | 7 | 699-713 |
| Chen Q, LM Hildemann | The Effects of Human Activities on Exposure to Particulate Matter and Bioaerosols in Residential Homes | Environmental Science & Technology | 2009 | 43 | 13 | 4641-4646 |
| Cheng KC, HK Park, AO Tetteh, D Zheng, NT Ouellette, KC Nadeau, LM Hildemann | Mixing and sink effects of air purifiers on indoor PM2.5 concentrations: A pilot study of eight residential homes in Fresno, California | Aerosol Science and Technology | 2016 | 50 | 8 | 835-845 |

| | | | | | | |
|---|---|---|---|---|---|---|
| Clayton CA, RL Perritt, ED Pellizzari, KW Thomas, RW Whitmore, LA Wallace, H Ozkaynak, JD Spengler | PARTICLE TOTAL EXPOSURE ASSESSMENT METHODOLOGY (PTEAM) STUDY - DISTRIBUTIONS OF AEROSOL AND ELEMENTAL CONCENTRATIONS IN PERSONAL, INDOOR, AND OUTDOOR AIR SAMPLES IN A SOUTHERN CALIFORNIA COMMUNITY | Journal of Exposure Analysis and Environmental Epidemiology | 1993 | 3 | 2 | 227-250 |
| Clougherty JE, EA Houseman, JI Levy | Source apportionment of indoor residential fine particulate matter using land use regression and constrained factor analysis | Indoor Air | 2011 | 21 | 1 | 53-66 |
| Colome SD, NY Kado, P Jaques, M Kleinman | INDOOR OUTDOOR AIR-POLLUTION RELATIONS - PARTICULATE MATTER LESS THAN 10 MU IN AERODYNAMIC DIAMETER (PM-10) IN HOMES OF ASTHMATICS | Atmospheric Environment Part a-General Topics | 1992 | 26 | 12 | 2173-2178 |
| Colton MD, P MacNaughton, J Vallarino, J Kane, M Bennett-Fripp, JD Spengler, G Adamkiewicz | Indoor Air Quality in Green Vs Conventional Multifamily Low-Income Housing | Environmental Science & Technology | 2014 | 48 | 14 | 7833-7841 |
| Coombs KC, GL Chew, C Schaffer, PH Ryan, C Brokamp, SA Grinshpun, G Adamkiewicz, S Chillrud, C Hedman, M Colton, J Ross, T Reponen | Indoor air quality in green-renovated vs. non-green low-income homes of children living in a temperate region of US (Ohio) | Science of the Total Environment | 2016 | 554 | NA | 178-185 |
| Corsi RL, JA Siegel, C Chiang | Particle resuspension during the use of vacuum cleaners on residential carpet | Journal of Occupational and Environmental Hygiene | 2008 | 5 | 4 | 232-238 |

| | | | | | | |
|---|---|---|---|---|---|---|
| Cox J, K Isiugo, P Ryan, SA Grinshpun, M Yermakov, C Desmond, R Jandarov, S Vesper, J Ross, S Chillrud, K Dannemiller, T Reponen | Effectiveness of a portable air cleaner in removing aerosol particles in homes close to highways | Indoor Air | 2018 | 28 | 6 | 818-827 |
| Delfino RJ, N Staimer, T Tjoa | Personal endotoxin exposure in a panel study of school children with asthma | Environmental Health | 2011 | 10 | NA | NA |
| Delfino RJ, N Staimer, T Tjoa, A Polidori, M Arhami, DL Gillen, MT Kleinman, ND Vaziri, J Longhurst, F Zaldivar, C SioutaS | Circulating biomarkers of inflammation, antioxidant activity, and platelet activation are associated with primary combustion aerosols in subjects with coronary artery disease | Environmental Health Perspectives | 2008 | 116 | 7 | 898-906 |
| Delfino RJ, PJ Quintana, J Floro, VM Gastanaga, BS Samimi, MT Kleinman, LJ Liu, C Bufalino, CF Wu, CE McLaren | Association of FEV1 in asthmatic children with personal and microenvironmental exposure to airborne particulate matter | Environ Health Perspect | 2004 | 112 | 8 | 932-41 |
| Diette GB, NN Hansel, TJ Buckley, J Curtin-Brosnan, PA Eggleston, EC Matsui, MC McCormack, DL Williams, PN Breysse | Home indoor pollutant exposures among inner-city children with and without asthma | Environmental Health Perspectives | 2007 | 115 | 11 | 1665-1669 |
| Doll SC, EL Davison, BR Painting | Weatherization impacts and baseline indoor environmental quality in low income single-family homes | Building and Environment | 2016 | 107 | NA | 181-190 |
| Du L, S Batterman, E Parker, C Godwin, JY Chin, A O'Toole, T Robins, W Brakefield-Caldwell, T Lewis | Particle concentrations and effectiveness of free-standing air filters in bedrooms of children with asthma in Detroit, Michigan | Building and Environment | 2011 | 46 | 11 | 2303-2313 |

| | | | | | | |
|---|---|---|---|---|---|---|
| Eggleston PA, A Butz, C Rand, J Curtin-Brosnan, S Kanchanaraksa, L Swartz, P Breysse, T Buckley, G Diette, B Merriman, JA Krishnan | Home environmental intervention in inner-city asthma: a randomized controlled clinical trial | Annals of Allergy Asthma & Immunology | 2005 | 95 | 6 | 518-524 |
| Escobedo LE, WM Champion, N Li, LD Montoya | Indoor air quality in Latino homes in Boulder, Colorado | Atmospheric Environment | 2014 | 92 | NA | 69-75 |
| Evans GF, RV Highsmith, LS Sheldon, JC Suggs, RW Williams, RB Zweidinger, JP Creason, D Walsh, CE Rodes, PA Lawless | The 1999 Fresno particulate matter exposure studies: Comparison of community, outdoor, and residential PM mass measurements | Journal of the Air & Waste Management Association | 2000 | 50 | 11 | 1887-1896 |
| Fleisch AF, Rokoff LB, Garshick E, Grady ST, Chipman JW, Baker ER, et al. | Residential wood stove use and indoor exposure to PM2.5 and its components in Northern New England. | Journal of Exposure Science and Environmental Epidemiology | 2019 | NA | NA | Epub |
| Frey SE, H Destaillats, S Cohn, S Ahrentzen, MP Fraser | The effects of an energy efficiency retrofit on indoor air quality | Indoor Air | 2015 | 25 | 2 | 210-219 |
| Frey SE, H Destaillats, S Cohn, S Ahrentzen, MP Fraser | Characterization of indoor air quality and resident health in an Arizona senior housing apartment building | Journal of the Air & Waste Management Association | 2014 | 64 | 11 | 1251-1259 |
| Fuller CH, D Brugge, PL Williams, MA Mittleman, K Lane, JL Durant, JD Spengler | Indoor and outdoor measurements of particle number concentration in near-highway homes | Journal of Exposure Science and Environmental Epidemiology | 2013 | 23 | 5 | 506-512 |
| Geller MD, MH Chang, C Sioutas, BD Ostro, MJ Lipsett | Indoor/outdoor relationship and chemical composition of fine and coarse particles in the southern California deserts | Atmospheric Environment | 2002 | 36 | 6 | 1099-1110 |

| | | | | | | |
|---|---|---|---|---|---|---|
| Gould CF, SN Chillrud, D Phillips, MS Perzanowski, D Hernandez | Soot and the city: Evaluating the impacts of Clean Heat policies on indoor/outdoor air quality in New York City apartments | Plos One | 2018 | 13 | 6 | NA |
| Habre R, B Coull, E Moshier, J Godbold, A Grunin, A Nath, W Castro, N Schachter, A Rohr, M Kattan, J Spengler, P Koutrakis | Sources of indoor air pollution in New York City residences of asthmatic children | Journal of Exposure Science and Environmental Epidemiology | 2014 | 24 | 3 | 269-278 |
| Hansel NN, MC McCormack, AJ Belli, EC Matsui, RD Peng, C Aloe, L Paulin, DL Williams, GB Diette, PN Breysse | In-Home Air Pollution Is Linked to Respiratory Morbidity in Former Smokers with Chronic Obstructive Pulmonary Disease | American Journal of Respiratory and Critical Care Medicine | 2013 | 187 | 10 | 1085-1090 |
| Hart JE, Grady ST, Laden F, Coull BA, Koutrakis P, Schwartz JD, et al. | Effects of Indoor and Ambient Black Carbon and [Formula: see text] on Pulmonary Function among Individuals with COPD. | Environ Health Perspectives | 2018 | 126 | 12 | 1E+05 |
| Hart JF, TJ Ward, TM Spear, RJ Rossi, NN Holland, BG Loushin | Evaluating the Effectiveness of a Commercial Portable Air Purifier in Homes with Wood Burning Stoves: A Preliminary Study | Journal of Environmental and Public Health | 2011 | NA | NA | NA |
| Henderson DE, JB Milford, SL Miller | Prescribed burns and wildfires in Colorado: Impacts of mitigation measures on indoor air particulate matter | Journal of the Air & Waste Management Association | 2005 | 55 | 10 | 1516-1526 |
| Holm SM, J Balmes, D Gillette, K Hartin, E Seto, D Lindeman, D Polanco, E Fong | Cooking behaviors are related to household particulate matter exposure in children with asthma in the urban East Bay Area of Northern California | Plos One | 2018 | 13 | 6 | NA |

| | | | | | | |
|---|---|---|---|---|---|---|
| Howard-Reed C, AW Rea, MJ Zufall, JM Burke, RW Williams, JC Suggs, LS Sheldon, D Walsh, R Kwok | Use of a continuous nephelometer to measure personal exposure to particles during the US Environmental Protection Agency Baltimore and Fresno panel studies | Journal of the Air & Waste Management Association | 2000 | 50 | 7 | 1125-1132 |
| Hsu SI, K Ito, M Kendall, M Lippmann | Factors affecting personal exposure to thoracic and fine particles and their components | Journal of Exposure Science and Environmental Epidemiology | 2012 | 22 | 5 | 439-447 |
| Huang SD, J Lawrence, CM Kang, J Li, M Martins, P Vokonas, DR Gold, J Schwartz, BA Coull, P Koutrakis | Road proximity influences indoor exposures to ambient fine particle mass and components | Environmental Pollution | 2018 | 243 | NA | 978-987 |
| Hudda N, MC Simon, W Zamore, JL Durant | Aviation-Related Impacts on Ultrafine Particle Number Concentrations Outside and Inside Residences near an Airport | Environmental Science & Technology | 2018 | 52 | 4 | 1765-1772 |
| Hughes SC, Bellettiere J, Nguyen B, Liles S, Klepeis NE, Quintana PJE, et al. | Randomized Trial to Reduce Air Particle Levels in Homes of Smokers and Children. | American Journal of Preventive Medicine | 2018 | 54 | 3 | 359-67 |
| Hunt A, JA Crawford, PF Rosenbaum, JL Abraham | Levels of household particulate matter and environmental tobacco smoke exposure in the first year of life for a cohort at risk for asthma in urban Syracuse, NY | Environment International | 2011 | 37 | 7 | 1196-1205 |
| Isiugo K, Jandarov R, Cox J, Ryan P, Newman N, Grinshpun SA, et al. | Indoor particulate matter and lung function in children. | Science of the Total Environment | 2019 | 663 | NA | 408-17 |

| | | | | | | |
|---|---|---|---|---|---|---|
| Jansen KL, TV Larson, JQ Koenig, TF Mar, C Fields, J Stewart, M Lippmann | Associations between health effects and particulate matter and black carbon in subjects with respiratory disease | Environmental Health Perspectives | 2005 | 113 | 12 | 1741-1746 |
| Johnson R, J Schmid, S Dinakaran, R Seifert | Use of simulink for dynamic air quality modeling in interior Alaska | Journal of Cold Regions Engineering | 2005 | 19 | 1 | 43542 |
| Jung KH, D Torrone, S Lovinsky-Desir, M Perzanowski, J Bautista, JR Jezioro, L Hoepner, J Ross, FP Perera, SN Chillrud, RL Miller | Short-term exposure to PM2.5 and vanadium and changes in asthma gene DNA methylation and lung function decrements among urban children | Respiratory Research | 2017 | 18 | NA | NA |
| Jung KH, K Bernabe, K Moors, B Yan, SN Chillrud, R Whyatt, D Camann, PL Kinney, FP Perera, RL Miller | Effects of Floor Level and Building Type on Residential Levels of Outdoor and Indoor Polycyclic Aromatic Hydrocarbons, Black Carbon, and Particulate Matter in New York City | Atmosphere | 2011 | 2 | 2 | 96-109 |
| Jung KH, MM Patel, K Moors, PL Kinney, SN Chillrud, R Whyatt, L Hoepner, R Garfinkel, BZ Yan, J Ross, D Camann, FP Perera, RL Miller | Effects of heating season on residential indoor and outdoor polycyclic aromatic hydrocarbons, black carbon, and particulate matter in an urban birth cohort | Atmospheric Environment | 2010 | 44 | 36 | 4545-4552 |
| Kamens R, CT Lee, R Wiener, D Leith | A STUDY TO CHARACTERIZE INDOOR PARTICLES IN 3 NONSMOKING HOMES | Atmospheric Environment Part a-General Topics | 1991 | 25 | NA | 939-948 |
| Kang CM, D Gold, P Koutrakis | Downwind O-3 and PM2.5 speciation during the wildfires in 2002 and 2010 | Atmospheric Environment | 2014 | 95 | NA | 511-519 |

| | | | | | | |
|---|---|---|---|---|---|---|
| Keeler GJ, JT Dvonch, FY Yip, EA Parker, BA Israel, FJ Marsik, M Morishita, JA Barres, TG Robins, W Brakefield-Caldwell, M Sam | Assessment of personal and community-level exposures to particulate matter among children with asthma in Detroit, Michigan, as part of Community Action Against Asthma (CAAA) | Environmental Health Perspectives | 2002 | 110 | NA | 173-181 |
| Khurshid SS, JA Siegel, KA Kinney | Particulate reactive oxygen species on total suspended particles - measurements in residences in Austin, Texas | Indoor Air | 2016 | 26 | 6 | 953-963 |
| Khurshid, S.S., Siegel, J.A. and Kinney, KA | Indoor particulate reactive oxygen species concentrations | Environmental research | 2014 | 132 | NA | 46-53 |
| Kilburg-Basnyat B, TM Peters, SS Perry, PS Thorne | Electrostatic dust collectors compared to inhalable samplers for measuring endotoxin concentrations in farm homes | Indoor Air | 2016 | 26 | 5 | 724-733 |
| King, B.A., Travers, M.J., Cummings, K.M., Mahoney, M.C. and Hyland, A.J. | Secondhand smoke transfer in multiunit housing | Nicotine & Tobacco Research | 2010 | 12 | 11 | 1133-1141 |
| Kinney PL, SN Chillrud, S Ramstrom, J Ross, JD Spengler | Exposures to multiple air toxics in New York City | Environmental Health Perspectives | 2002 | 110 | NA | 539-546 |
| Klepeis, N.E., Bellettiere, J., Hughes, S.C., Nguyen, B., Berardi, V., Liles, S., Obayashi, S., Hofstetter, C.R., Blumberg, E. and Hovell, M.F. | Fine particles in homes of predominantly low-income families with children and smokers: Key physical and behavioral determinants to inform indoor-air-quality interventions | PloS one | 2017 | 12 | 5 | e0177718 |
| Koehler K, Good N, Wilson A, Molter A, Moore BF, Carpenter T, et al. | The Fort Collins commuter study: Variability in personal exposure to air pollutants by microenvironment. | Indoor Air: Interational Journal of Indoor Environment and Health | 2019 | 29 | 2 | 231-41 |

| | | | | | | |
|---|---|---|---|---|---|---|
| Kopperud RJ, AR Ferro, LM Hildemann | Outdoor versus indoor contributions to indoor particulate matter (PM) determined by mass balance methods | Journal of the Air & Waste Management Association | 2004 | 54 | 9 | 1188-1196 |
| Lachenmyer C, GM Hidy | Urban measurements of outdoor-indoor PM25 concentrations and personal exposure in the deep south. Part I. Pilot study of mass concentrations for nonsmoking subjects | Aerosol Science and Technology | 2000 | 32 | 1 | 34-51 |
| Leaderer BP, L Naeher, T Jankun, K Balenger, TR Holford, C Toth, J Sullivan, JM Wolfson, P Koutrakis | Indoor, outdoor, and regional summer and winter concentrations of PM10, PM2.5, SO42-, H+, NH4+, NO3-, NH3, and nitrous acid in homes with and without kerosene space heaters | Environmental Health Perspectives | 1999 | 107 | 3 | 223-231 |
| Leaderer BP, P Koutrakis, SLK Briggs, J Rizzuto | THE MASS CONCENTRATION AND ELEMENTAL COMPOSITION OF INDOOR AEROSOLS IN SUFFOLK AND ONONDAGA COUNTIES, NEW-YORK | Indoor Air-International Journal of Indoor Air Quality and Climate | 1994 | 4 | 1 | 23-34 |
| LeBouf R, L Yesse, A Rossner | Seasonal and diurnal variability in airborne mold from an indoor residential environment in northern New York | Journal of the Air & Waste Management Association | 2008 | 58 | 5 | 684-692 |
| Less, B., Mullen, N., Singer, B., & Walker, I | Indoor air quality in 24 California residences designed as high-performance homes | Science and Technology for the Built Environment | 2015 | 21 | 1 | 14-24 |
| Levy JI, T Dumyahn, JD Spengler | Particulate matter and polycyclic aromatic hydrocarbon concentrations in indoor and outdoor microenvironments in Boston, Massachusetts | Journal of Exposure Analysis and Environmental Epidemiology | 2002 | 12 | 2 | 104-114 |

| | | | | | | |
|---|---|---|---|---|---|---|
| Lewis CW | Sources of air pollutants indoors: VOC and fine particulate species | J Expo Anal Environ Epidemiol | 1991 | 1 | 1 | 31-44 |
| Li WW, H Paschold, H Morales, J Chianelli | Correlations between short-term indoor and outdoor PM concentrations at residences with evaporative coolers | Atmospheric Environment | 2003 | 37 | 19 | 2691-2703 |
| Linn WS, H Gong, KW Clark, KR Anderson | Day-to-day particulate exposures and health changes in Los Angeles area residents with severe lung disease | Journal of the Air & Waste Management Association | 1999 | 49 | NA | 108-115 |
| Lioy PJ, NC Freeman, T Wainman, AH Stern, R Boesch, T Howell, SI Shupack | Microenvironmental analysis of residential exposure to chromium-laden wastes in and around New Jersey homes | Risk Anal | 1992 | 12 | 2 | 287-99 |
| Lioy, P. J., Waldman, J. M., Buckley, T., Butler, J., & Pietarinen, C. | The personal, indoor and outdoor concentrations of PM-10 measured in an industrial community during the winter | Atmospheric Environment. Part B. Urban Atmosphere | 1990 | 24 | 1 | 57-66 |
| Liu LJ, M Box, D Kalman, J Kaufman, J Koenig, T Larson, T Lumley, L Sheppard, L Wallace | Exposure assessment of particulate matter for susceptible populations in Seattle | Environ Health Perspect | 2003 | 111 | 7 | 909-18 |
| Long CM, HH Suh, L Kobzik, PJ Catalano, YY Ning, P Koutrakis | A pilot investigation of the relative toxicity of indoor and outdoor fine particles: In vitro effects of endotoxin and other particulate properties | Environmental Health Perspectives | 2001 | 109 | 10 | 1019-1026 |
| Long CM, HH Suh, P Koutrakis | Characterization of indoor particle sources using continuous mass and size monitors | Journal of the Air & Waste Management Association | 2000 | 50 | 7 | 1236-1250 |

| | | | | | | |
|---|---|---|---|---|---|---|
| Long CM, HH Suh, PJ Catalano, P Koutrakis | Using time- and size-resolved particulate data to quantify indoor penetration and deposition behavior | Environmental Science & Technology | 2001 | 35 | 10 | 2089-2099 |
| MacNaughton P, G Adamkiewicz, RE Arku, J Vallarino, DE Levy | The impact of a smoke-free policy on environmental tobacco smoke exposure in public housing developments | Science of the Total Environment | 2016 | 557 | NA | 676-680 |
| Maestas MM, Brook RD, Ziemba RA, Li FY, Crane RC, Klaver ZM, et al. | Reduction of personal PM2.5 exposure via indoor air filtration systems in Detroit: an intervention study. | Journal of Exposure Science and Environmental Epidemiology | 2019 | 29 | 4 | 484-90 |
| Martuzevicius D, SA Grinshpun, T Lee, SH Hu, P Biswas, T Reponen, G LeMasters | Traffic-related PM2.5 aerosol in residential houses located near major highways: Indoor versus outdoor concentrations | Atmospheric Environment | 2008 | 42 | 27 | 6575-6585 |
| McAuley TR, R Fisher, X Zhou, PA Jaques, AR Ferro | Relationships of outdoor and indoor ultrafine particles at residences downwind of a major international border crossing in Buffalo, NY | Indoor Air | 2010 | 20 | 4 | 298-308 |
| McCormack MC, AJ Belli, D Waugh, EC Matsui, RD Peng, DL Williams, L Paulin, A Saha, CM Aloe, GB Diette, PN Breysse, NN Hansel | Respiratory Effects of Indoor Heat and the Interaction with Air Pollution in Chronic Obstructive Pulmonary Disease | Annals of the American Thoracic Society | 2016 | 13 | 12 | 2125-2131 |
| McCormack MC, PN Breysse, EC Matsui, NN Hansel, D Williams, J Curtin-Brosnan, P Eggleston, GB Diette, E Ctr Childhood Asthma Urban | In-Home Particle Concentrations and Childhood Asthma Morbidity | Environmental Health Perspectives | 2009 | 117 | 2 | 294-298 |

| | | | | | | |
|---|---|---|---|---|---|---|
| McCormack MC, PN Breysse, NN Hansel, EC Matsui, ES Tonorezos, J Curtin-Brosnan, DL Williams, TJ Buckley, PA Eggleston, GB Diette | Common household activities are associated with elevated particulate matter concentrations in bedrooms of inner-city Baltimore pre-school children | Environmental Research | 2008 | 106 | 2 | 148-155 |
| Militello-Hourigan RE, SL Miller | The impacts of cooking and an assessment of indoor air quality in Colorado passive and tightly constructed homes | Building and Environment | 2018 | 144 | NA | 573-582 |
| Miller SL, P Scaramella, J Campe, CW Goss, S Diaz-Castillo, E Hendrikson, C DiGuiseppi, J Litt | An assessment of indoor air quality in recent Mexican immigrant housing in Commerce City, Colorado | Atmospheric Environment | 2009 | 43 | 35 | 5661-5667 |
| Min KT, Lundrigan P, Sward K, Collingwood SC, Patwari N. | Smart home air filtering system: A randomized controlled trial for performance evaluation. | Smart Health | 2018 | NA | NA | 62-75 |
| Mukerjee S, WD Ellenson, RG Lewis, RK Stevens, MC Somerville, DS Shadwick, RD Willis | An environmental scoping study in the Lower Rio Grande Valley of Texas .3. Residential microenvironmental monitoring for air, house dust, and soil | Environment International | 1997 | 23 | 5 | 657-673 |
| Na K, DR Cocker | Organic and elemental carbon concentrations in fine particulate matter in residences, schoolrooms, and outdoor air in Mira Loma, California | Atmospheric Environment | 2005 | 39 | 18 | 3325-3333 |
| Noonan CW, TJ Ward, W Navidi, L Sheppard, M Bergauff, C Palmer | Assessing the impact of a wood stove replacement program on air quality and children's health | Res Rep Health Eff Inst | 2011 | NA | 162 | 3-37; discussion 39-47 |

| | | | | | | |
|---|---|---|---|---|---|---|
| Olson DA, J Turlington, RV Duvall, SR Vicdow, CD Stevens, R Williams | Indoor and outdoor concentrations of organic and inorganic molecular markers: Source apportionment of PM2.5 using low-volume samples | Atmospheric Environment | 2008 | 42 | 8 | 1742-1751 |
| Patton AP, L Calderon, YY Xiong, ZC Wang, J Senick, MS Allacci, D Plotnik, R Wener, CJ Andrews, U Krogmann, G Mainelis | Airborne Particulate Matter in Two Multi-Family Green Buildings: Concentrations and Effect of Ventilation and Occupant Behavior | International Journal of Environmental Research and Public Health | 2016 | 13 | 1 | NA |
| Pavilonis BT, TR Anthony, PT O'Shaughnessy, MJ Humann, JA Merchant, G Moore, PS Thorne, CP Weisel, WT Sanderson | Indoor and outdoor particulate matter and endotoxin concentrations in an intensely agricultural county | Journal of Exposure Science and Environmental Epidemiology | 2013 | 23 | 3 | 299-305 |
| Pellizzari ED, CA Clayton, CE Rodes, RE Mason, LL Piper, B Fort, G Pfeifer, D Lynam | Particulate matter and manganese exposures in Indianapolis, Indiana | Journal of Exposure Analysis and Environmental Epidemiology | 2001 | 11 | 6 | 423-440 |
| Pickett AR, ML Bell | Assessment of Indoor Air Pollution in Homes with Infants | International Journal of Environmental Research and Public Health | 2011 | 8 | 12 | 4502-4520 |
| Robin LF, PS Less, M Winget, M Steinhoff, LH Moulton, M Santosham, A Correa | Wood-burning stoves and lower respiratory illnesses in Navajo children | Pediatr Infect Dis J | 1996 | 15 | 10 | 859-65 |
| Rodes CE, PA Lawless, JW Thornburg, RW Williams, CW Croghan | DEARS particulate matter relationships for personal, indoor, outdoor, and central site settings for a general population | Atmospheric Environment | 2010 | 44 | 11 | 1386-1399 |

| | | | | | | |
|---|---|---|---|---|---|---|
| Rojas-Bracho L, HH Suh, P Koutrakis | Relationships among personal, indoor, and outdoor fine and coarse particle concentrations for individuals with COPD | Journal of Exposure Analysis and Environmental Epidemiology | 2000 | 10 | 3 | 294-306 |
| Russo ET, TE Hulse, G Adamkiewicz, DE Levy, L Bethune, J Kane, M Reid, SN Shah | Comparison of Indoor Air Quality in Smoke-Permitted and Smoke-Free Multiunit Housing: Findings From the Boston Housing Authority | Nicotine & Tobacco Research | 2015 | 17 | 3 | 316-322 |
| Sarnat SE, BA Coull, PA Ruiz, P Koutrakis, HH Suh | The influences of ambient particle composition and size on particle infiltration in Los Angeles, CA, residences | Journal of the Air & Waste Management Association | 2006 | 56 | 2 | 186-196 |
| Sawant AA, K Na, XN Zhu, DR Cocker | Chemical characterization of outdoor PM2.5 and gas-phase compounds in Mira Loma, California | Atmospheric Environment | 2004 | 38 | 0 | 5517-5528 |
| Sawant AA, K Na, XN Zhu, K Cocker, S Butt, C Song, DR Cocker | Characterization of PM2.5 and selected gas-phase compounds at multiple indoor and outdoor sites in Mira Loma, California | Atmospheric Environment | 2004 | 38 | 37 | 6269-6278 |
| Shalat SL, AA Stambler, Z Wang, G Mainelis, OH Emoekpere, M Hernandez, PJ Lioy, K Black | Development and in-home testing of the Pretoddler Inhalable Particulate Environmental Robotic (PIPER Mk IV) sampler | Environ Sci Technol | 2011 | 45 | 7 | 2945-50 |
| Shalat SL, PJ Lioy, K Schmeelck, G Mainelis | Improving estimation of indoor exposure to inhalable particles for children in the first year of life | Journal of the Air & Waste Management Association | 2007 | 57 | 8 | 934-939 |
| Shrestha PM, Humphrey JL, Carlton EJ, Adgate JL, Barton KE, Root ED, et al. | Impact of Outdoor Air Pollution on Indoor Air Quality in Low-Income Homes during Wildfire Seasons. | International Journal of Environmental Research and Public Health | 2019 | 16 | 19 | NA |

| | | | | | | |
|---|---|---|---|---|---|---|
| Simons E, J Curtin-Brosnan, T Buckley, P Breysse, PA Eggleston | Indoor environmental differences between inner city and suburban homes of children with asthma | Journal of Urban Health-Bulletin of the New York Academy of Medicine | 2007 | 84 | 4 | 577-590 |
| Singer BC, RZ Pass, WW Delp, DM Lorenzetti, RL Maddalena | Pollutant concentrations and emission rates from natural gas cooking burners without and with range hood exhaust in nine California homes | Building and Environment | 2017 | 122 | NA | 215-229 |
| Thatcher TL, DW Layton | DEPOSITION, RESUSPENSION, AND PENETRATION OF PARTICLES WITHIN A RESIDENCE | Atmospheric Environment | 1995 | 29 | 13 | 1487-1497 |
| Thomas NM, Calderón L, Senick J, Sorensen-Allacci M, Plotnik D, Guo M, et al. | Investigation of indoor air quality determinants in a field study using three different data streams. | Building and Environment | 2019 | 154 | NA | 281-95 |
| Tryner J, Quinn C, Windom BC, Volckens J. | Design and evaluation of a portable PM2.5 monitor featuring a low-cost sensor in line with an active filter sampler. | Environmental Science-Processes & Impacts | 2019 | 21 | 8 | 1403-15 |
| Tunno BJ, S Kyra Naumoff, L Cambal, S Tripathy, F Holguin, P Lioy, JE Clougherty | Indoor air sampling for fine particulate matter and black carbon in industrial communities in Pittsburgh | Sci Total Environ | 2015 | 536 | NA | 108-115 |
| Turpin BJ, CP Weisel, M Morandi, S Colome, T Stock, S Eisenreich, B Buckley | Relationships of Indoor, Outdoor, and Personal Air (RIOPA): part II. Analyses of concentrations of particulate matter species | Res Rep Health Eff Inst | 2007 | NA | NA | 1-77; discussion 79-92 |
| Van Deusen A, A Hyland, MJ Travers, C Wang, C Higbee, BA King, T Alford, KM Cummings | Secondhand smoke and particulate matter exposure in the home | Nicotine & Tobacco Research | 2009 | 11 | 6 | 635-641 |

| | | | | | | |
|---|---|---|---|---|---|---|
| Vette A, J Burke, G Norris, M Landis, S Batterman, M Breen, V Isakov, T Lewis, MI Gilmour, A Kamal, D Hammond, R Vedantham, S Bereznicki, N Tian, C Croghan, S Community Action Against Asthma | The Near-Road Exposures and Effects of Urban Air Pollutants Study (NEXUS): Study design and methods | Science of the Total Environment | 2013 | 448 | NA | 38-47 |
| Wallace L | Indoor sources of ultrafine and accumulation mode particles: Size distributions, size-resolved concentrations, and source strengths | Aerosol Science and Technology | 2006 | 40 | 5 | 348-360 |
| Wallace L | Ultrafine particles from a vented gas clothes dryer | Atmospheric Environment | 2005 | 39 | 32 | 5777-5786 |
| Wallace L, R Williams, A Rea, C Croghan | Continuous weeklong measurements of personal exposures and indoor concentrations of fine particles for 37 health-impaired North Carolina residents for up to four seasons | Atmospheric Environment | 2006 | 40 | 3 | 399-414 |
| Wallace L, W Ott | Personal exposure to ultrafine particles | Journal of Exposure Science and Environmental Epidemiology | 2011 | 21 | 1 | 20-30 |
| Wallace LA, H Mitchell, GT O'Connor, L Neas, M Lippmann, M Kattan, J Koenig, JW Stout, BJ Vaughn, D Wallace, M Walter, K Adams, LJ Liu | Particle concentrations in inner-city homes of children with asthma: the effect of smoking, cooking, and outdoor pollution | Environ Health Perspect | 2003 | 111 | 9 | 1265-72 |

| | | | | | | |
|---|---|---|---|---|---|---|
| Wallace LA, SJ Emmerich, C Howard-Reed | Source strengths of ultrafine and fine particles due to cooking with a gas stove | Environmental Science & Technology | 2004 | 38 | 8 | 2304-2311 |
| Wallace, L; Williams, R; Suggs, J; Sheldon, L; Zweidinger, R; Rea, A; Vette, A; C; Rodes, C; Lawless, P; Thornburg, J; Liu, LJ; Allen, R; Kalman, D; Kaufman, J; Koenig, J; Larson, T; Brown K; Sarnat J; Suh, H; Wheeler, A; Koutrakis, P | Exposure of High Risk Subpopulations to Particles: Final Report (APM 21) | EPA report (peer reviewed) | 2003 | 0 | 0 | NA |
| Ward T, C Noonan | Results of a residential indoor PM(2.5) sampling program before and after a woodstove changeout | Indoor Air | 2008 | 18 | 5 | 408-415 |
| Ward T, J Boulafentis, J Simpson, C Hester, T Moliga, K Warden, C Noonan | Lessons learned from a woodstove changeout on the Nez Perce Reservation | Science of the Total Environment | 2011 | 409 | 4 | 664-670 |
| Ward TJ, EO Semmens, E Weiler, S Harrar, CW Noonan | Efficacy of interventions targeting household air pollution from residential wood stoves | Journal of Exposure Science and Environmental Epidemiology | 2017 | 27 | 1 | 64-71 |
| Waring MS, JA Siegel | The effect of an ion generator on indoor air quality in a residential room | Indoor Air | 2011 | 21 | 4 | 267-276 |
| Williams DL, PN Breysse, MC McCormack, GB Diette, S McKenzie, AS Geyh | Airborne cow allergen, ammonia and particulate matter at homes vary with distance to industrial scale dairy operations: an exposure assessment | Environmental Health | 2011 | 10 | NA | NA |
| Williams R, AG Rappold, M Case, M Schmitt, S | Multi-pollutant exposures in an asthmatic cohort | Atmospheric Environment | 2012 | 61 | NA | 244-252 |

| | | | | | | |
|---|---|---|---|---|---|---|
| Stone, P Jones, J Thornburg, RB Devlin | | | | | | |
| Williams R, J Creason, R Zweidinger, R Watts, L Sheldon, C Shy | Indoor, outdoor, and personal exposure monitoring of particulate air pollution: the Baltimore elderly epidemiology-exposure pilot study | Atmospheric Environment | 2000 | 34 | 24 | 4193-4204 |
| Williams R, J Suggs, A Rea, K Leovic, A Vette, C Croghan, L Sheldon, C Rodes, J Thornburg, A Ejire, M Herbst, W Sanders | The Research Triangle Park particulate matter panel study: PM mass concentration relationships | Atmospheric Environment | 2003 | 37 | 38 | 5349-5363 |
| Williams R, J Suggs, R Zweidinger, G Evans, J Creason, R Kwok, C Rodes, P Lawless, L Sheldon | The 1998 Baltimore particulate matter Epidemiology-Exposure Study: Part 1. Comparison of ambient, residential outdoor, indoor and apartment particulate matter monitoring | Journal of Exposure Analysis and Environmental Epidemiology | 2000 | 10 | 6 | 518-532 |
| Williams, R.O.N., Rea, A., Vette, A., Croghan, C., Whitaker, D., Stevens, C., Mcdow, S., Fortmann, R., Sheldon, L., Wilson, H., Thornburg, J. | The design and field implementation of the Detroit Exposure and Aerosol Research Study | Journal of Exposure Science and Environmental Epidemiology | 2009 | 19 | 7 | 643-659 |
| Wu CF, RJ Delfino, JN Floro, PJE Quintana, BS Samimi, MT Kleinman, RW Allen, LJS Liu | Exposure assessment and modeling of particulate matter for asthmatic children using personal nephelometers | Atmospheric Environment | 2005 | 39 | 19 | 3457-3469 |
| Xiong Y, U Krogmann, G Mainelis, LA Rodenburg, CJ Andrews | Indoor air quality in green buildings: A case-study in a residential high-rise building in the northeastern United States | J Environ Sci Health A Tox Hazard Subst Environ Eng | 2015 | 50 | 3 | 225-42 |

| | | | | | | |
|---|---|---|---|---|---|---|
| Xu Y, S Raja, AR Ferro, PA Jaques, PK Hopke, C Gressani, LE Wetzel | Effectiveness of heating, ventilation and air conditioning system with HEPA filter unit on indoor air quality and asthmatic children's health | Building and Environment | 2010 | 45 | 2 | 330-337 |
| Yip FY, GJ Keeler, JT Dvonch, TG Robins, EA Parker, BA Israel, W Brakefield-Caldwell | Personal exposures to particulate matter among children with asthma in Detroit, Michigan | Atmospheric Environment | 2004 | 38 | 31 | 5227-5236 |
| Zamora ML, JC Pulczinski, N Johnson, R Garcia-Hernandez, A Rule, G Carrillo, J Zietsman, B Sandragorsian, S Vallamsundar, MH Askariyeh, K Koehler | Maternal exposure to PM2.5 in south Texas, a pilot study | Science of the Total Environment | 2018 | NA | NA | 1497-1507 |
| Zanobetti A, PH Stone, FE Speizer, JD Schwartz, BA Coull, HH Suh, BD Nearing, MA Mittleman, RL Verrier, DR Gold | T-Wave Alternans, Air Pollution and Traffic in High-Risk Subjects | American Journal of Cardiology | 2009 | 104 | 5 | 665-670 |
| Zhang T, Chillrud SN, Yang Q, Pitiranggon M, Ross J, Perera F, et al. | Characterizing peak exposure of secondhand smoke using a real-time PM2.5 monitor. | Indoor Air: Interational Journal of Indoor Environment and Health | 2019 | NA | NA | Epub |
| Zhu YF, WC Hinds, M Krudysz, T Kuhn, J Froines, C Sioutas | Penetration of freeway ultrafine particles into indoor environments | Journal of Aerosol Science | 2005 | 36 | 3 | 303-322 |

| **Vietnam** | | | | | | |
|---|---|---|---|---|---|---|
| Quang TN, NT Hue, P Thai, M Mazaheri, L Morawska | Exploratory assessment of indoor and outdoor particle number concentrations in Hanoi households | Science of the Total Environment | 2017 | 599 | NA | 284-290 |
